# Risk assessment of *Escherichia coli* O157:H7 along the farm-to-fork fresh-cut romaine lettuce supply chain

**DOI:** 10.1101/2025.01.26.25321140

**Authors:** Ece Bulut, Sarah I. Murphy, Laura K. Strawn, Michelle D. Danyluk, Martin Wiedmann, Renata Ivanek

**Affiliations:** Department of Population Medicine and Diagnostic Sciences, Cornell University, Ithaca, NY 14853, USA; Department of Food Science and Technology, Virginia Tech, Blacksburg, VA 24061, USA; Food Science and Human Nutrition Department, University of Florida, Lake Alfred, FL 33850, USA; Department of Food Science, Cornell University, Ithaca, NY 14853, USA

**Keywords:** Quantitative microbial risk assessment, Escherichia coli O157:H7, lettuce, farm to fork

## Abstract

Outbreaks of *Escherichia coli* O157:H7 (ECO157) in romaine lettuce remain an ongoing public health concern. Quantitative microbial risk assessment (QMRA) models are key tools for identifying control measures to mitigate foodborne diseases. Here, we introduce a comprehensive QMRA framework along the farm-to-fork fresh-cut romaine lettuce chain, including a novel preharvest difference equation model, to predict annual ECO157 illness cases in the United States and evaluate control strategies. We demonstrated the importance of managing irrigation-related contamination at preharvest to control illness cases. Wildlife intrusions pose lower health risk, followed by runoffs and biological soil amendments of animal origin. When preharvest contamination persists and combines with time-temperature abuses at postharvest, the predicted ECO157 illness cases rise considerably. We showed a broad range of interventions targeting both preharvest and postharvest stages can effectively improve the microbial safety of fresh-cut romaine. The comprehensive practices and interventions explored in this study will aid decision-makers in establishing/enhancing food safety best management practices.

## Main

The number of detected foodborne illness outbreaks linked to leafy greens has increased in the United States (USA) between 1996 and 2016, with *Escherichia coli* O157:H7 (ECO157)-contaminated romaine lettuce emerging as the leading source, followed by other common serotypes such as ECO26 and ECO111 *[1]*. Between 2015 and 2021, ECO157-contaminated romaine has been implicated in 7 outbreaks in the USA, 6 of which were multistate incidents *[2,3]*. These outbreaks resulted in 4,274 laboratory-confirmed illnesses, leading to 766 hospitalizations and 11 deaths.

Contamination of leafy greens, including romaine, can occur at any point during preharvest, harvest and postharvest, involving various risk factors *[4]*. Preharvest risk factors may involve domestic animals in the fields *[5,6]*, wild animals *[4,7-9]*, biological soil amendments of animal origin (BSAAO) amended soil *[10]*, irrigation water *[11]* and runoff from animal operations *[12,13]*. Harvest and postharvest risk factors may include human behavior [14], harvesting/production equipment *[15,16]* and production water *[17]*. In addition, the lack of processing steps that ensure effective removal or inactivation of ECO157 before consumption *[14]*, coupled with the recent increase in romaine consumption *[18]*, further elevates the risk.

In the USA, 90% of the leafy greens are grown by the members of the California or Arizona Leafy Green Products Handler Marketing Agreements (LGMA) *[19,20]*. These agreements have established guidelines to minimize contamination during the production and harvesting of leafy greens. The guidelines cover best practices in areas such as personnel training, agricultural water management, soil amendments, harvest equipment cleaning, packaging, transportation, produce testing, and prevention of animal intrusion. For example, both LGMAs prohibit the use of Type B agricultural waters—potentially contaminated with fecal indicators—for overhead spray irrigation of leafy greens within 21 days of harvest. If Type B water is used, it must be treated to meet Type A standards (agricultural water unlikely to contain fecal contamination) and must pass microbiological testing criteria. Additionally, the use of untreated, un-composted, incompletely composted or non-thermally treated BSAAOs is prohibited. Leafy greens cannot be produced from fields where such BSAAOs have been applied until one year has passed. If wildlife intrusion is detected, affected crops must be isolated within a three-foot radius. Furthermore, the position of animal feeding operations near leafy green farms must be assessed for any issues that might impact safety. The FDA Food Safety Modernization Act (FSMA) Produce Safety Rule (PSR) mandates similar standards for agricultural water, soil amendments, animal management, personnel training and equipment cleaning *[21]*. While FSMA PSR does not require microbiological testing of agricultural water *[22]*, it mandates routine inspections to ensure agricultural water safety and proper maintenance *[22]*. The application of raw BSAAOs is regulated by the ‘90-120-day’ rule, which requires that raw BSAAOs be incorporated into the soil at least 120 days before harvest *[23]*. Stabilized compost must meet a ECO157 count of < 0.3 MPN per gram of BSAAO *[24]*. Produce likely to be contaminated by animal intrusion should not be harvested *[25]*. The impact of nearby animal feeding operations should be evaluated based on their effects on agricultural water, such as drainage or discharge of untreated water *[26]*.

Quantitative microbial risk assessment (QMRA) models are commonly used to quantify the risk of microbial contamination or illness associated with exposure to microbial hazards. The outcome of these models provides valuable information for decision makers for setting regulatory standards, designing interventions to reduce microbial risks, or assessing the effectiveness of control measures *[27]*. Over the years, numerous QMRA models have been developed for leafy greens contaminated with *E. coli* strains, including ECO157 *[28-41]*. These risk assessments have investigated numerous pathways of *E. coli* contamination along the farm-to-fork leafy greens supply chain. However, none had comprehensively evaluated a broader range of factors related to ECO157 exposure and transmission, particularly for the preharvest lettuce supply chain. Furthermore, there is no standard QMRA model currently in use by governmental agencies or industry for ECO157 on lettuce. While some tools, such as FDA-iRISK *[42]* and microbial risk assessments *[43,44]* or guidelines [45] from the USDA (developed over 20 years ago) exist, they do not specifically account for the health risks associated with ECO157 contamination of lettuce or leafy greens. Thus, an updated and comprehensive risk assessment tool for ECO157 on lettuce is needed to thoroughly evaluate the potential health implications of a wider range of exposure and transmission factors for ECO157 on lettuce.

We developed a comprehensive QMRA aiming to model ECO157 contamination along the fresh-cut romaine lettuce production and consumption chain in the USA. Our model incorporates a broad set of factors, utilizes a novel difference equation modeling approach at the preharvest stage, and accounts for ECO157 exposure and spread throughout the farm-to-fork chain, as informed by existing literature and expert elicitation. This QMRA was designed as a decision support tool for managing various risk factors of ECO157 contamination in the USA and is publicly available via a GitHub repository (https://github.com/IvanekLab/Romaine_ECO157_QMRA/).

## Methods

### Overview of the model

In this study, we adapted elements from QMRAs developed for leafy greens in Spain *[27]*, Belgium *[39]* and USA *[32,36,38]*. We integrated these QMRAs with a modeling method for partitioning and mixing processes for microbial organisms in the food supply chain [46], as well as the concept of non-persister and persister cell state dynamics *[47]* for modeling biphasic decay of ECO157 on romaine and in soil. We also used the FSMA PSR, California and Arizona LGMA guidelines, and an expert opinions *[48]* to capture practices specific to the USA. This resulted in a comprehensive risk assessment model that simulates various field and processing/handling conditions. We predicted the number of annual illness cases in the USA attributed to consuming ECO157-contaminated fresh-cut romaine, which is our main model output. We integrated two models into an exposure assessment: the preharvest model (including preharvest and harvest sub-models) and postharvest model (including processing-retail, and consumer sub-models). In the preharvest model, we used a difference equation model to track the population dynamics of ECO157 in soil and romaine prior to harvest. Figure 1 provides an overview of the QMRA model’s structure and Figure 2 describes the parameters used in the model. The unit of analysis in the preharvest model was a romaine ‘batch’, but it becomes a ‘package’ and then a ‘serving’ in the postharvest model. We defined a ‘lot’ as all romaine packages produced in a day between two cleaning and sanitation shifts. We assumed that a lot was harvested from a single 1-acre field and processed in a day. Based on this assumption, each lot contained 12 romaine batches or 26,000 romaine plants. To quantify the variability and uncertainty of the model outcomes, we evaluated the variability and uncertainty separately in a one-dimensional Monte Carlo simulation using Latin Hypercube Sampling with 100,000 iterations. In the variability analysis, variable parameters changed while uncertain parameters remained fixed to their medians—and vice versa for uncertainty analysis. This approach maintained model interpretability while revealing the distinct impacts of variability and uncertainty on predictions. This was executed in Microsoft Excel 2016 (Microsoft Corporation, WA USA), utilizing the @RISK add-on software (version 8.8.1, Lumivero, CO USA). Model validation was carried out by comparing the predicted annual illness cases with the data from existing surveillance studies *[49,50]*. Expert elicitation, along with sensitivity and uncertainty analyses were conducted to guide the selection of promising risk management strategies and inform parameter choices in scenario analyses. Regression trees were used in the preharvest and postharvest models to identify interaction of factors that lead to ECO157 contamination in romaine and illness cases. The following sections detailed the baseline model structure.

**Figure 1.**
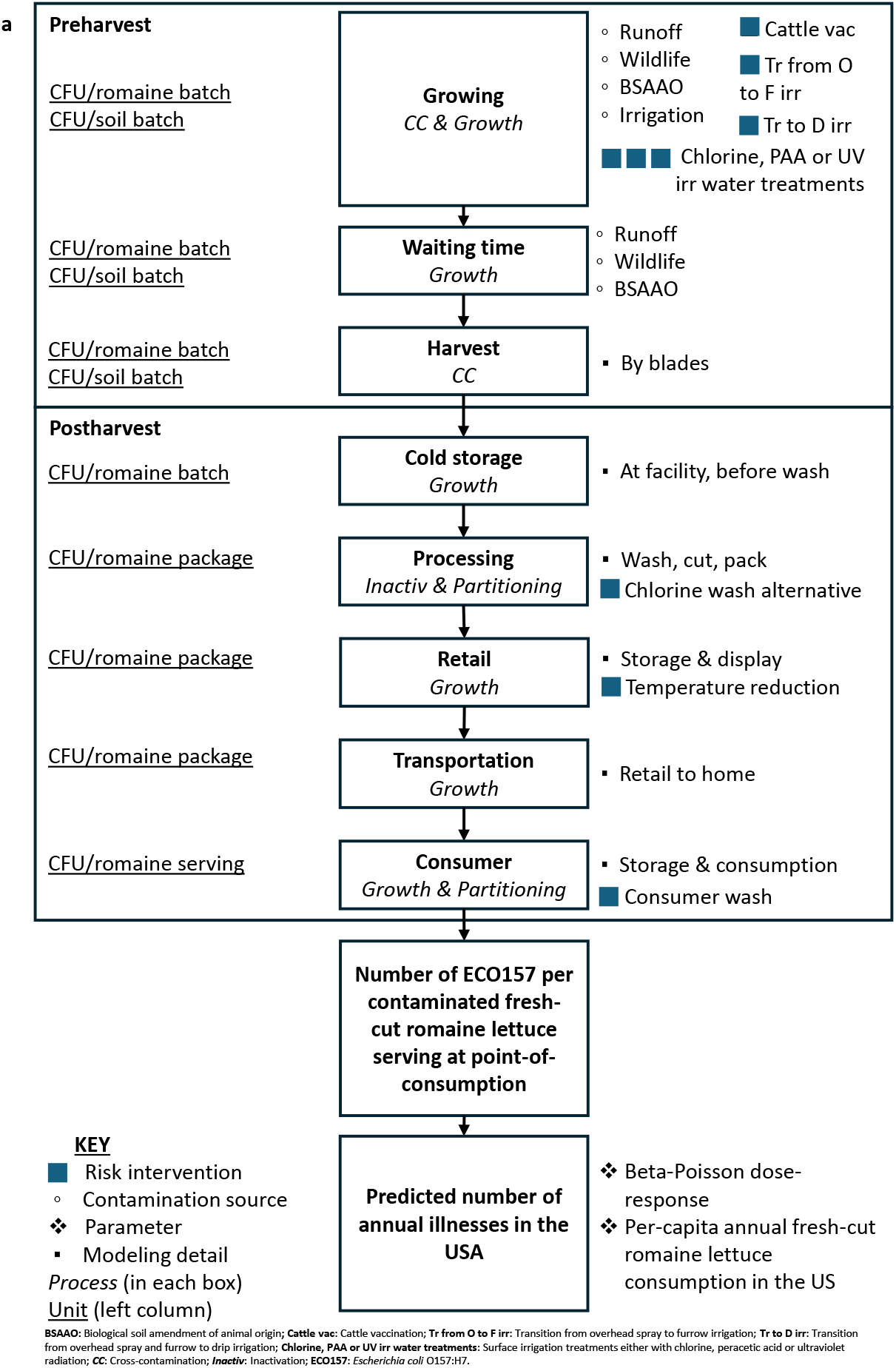
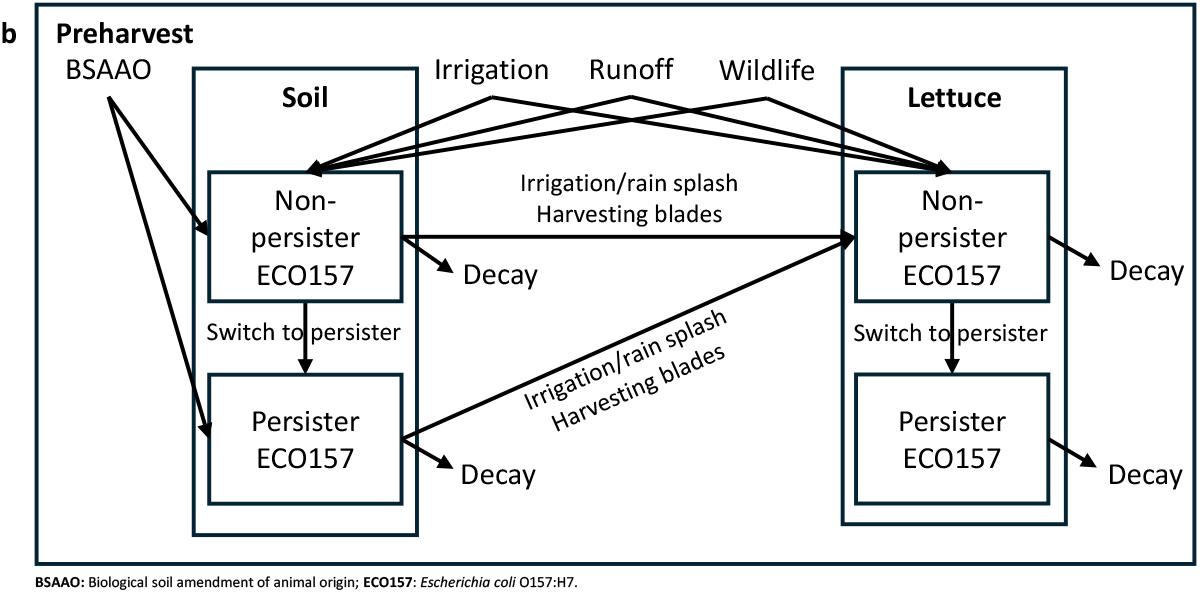
**(a)** Conceptual framework of the quantitative microbial risk assessment (QMRA) model for predicting illnesses due to Escherichia coli O157 (ECO157) contamination in the farm-to-fork fresh-cut romaine supply chain. **(b)** Conceptual framework of the difference equation model for population dynamics on the romaine batch and in soil batch during the 14-day preharvest countdown period.

**Figure 2.**
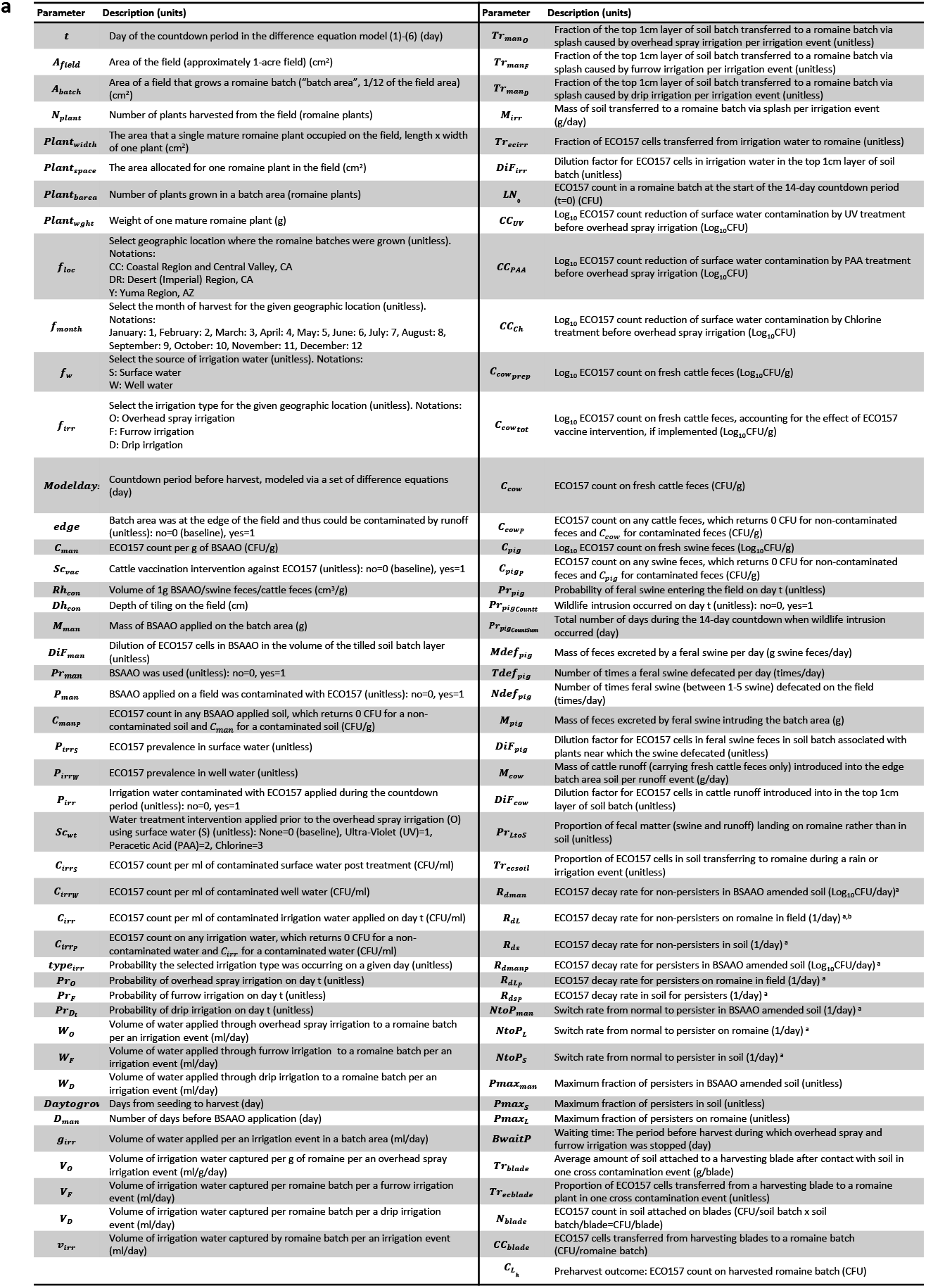
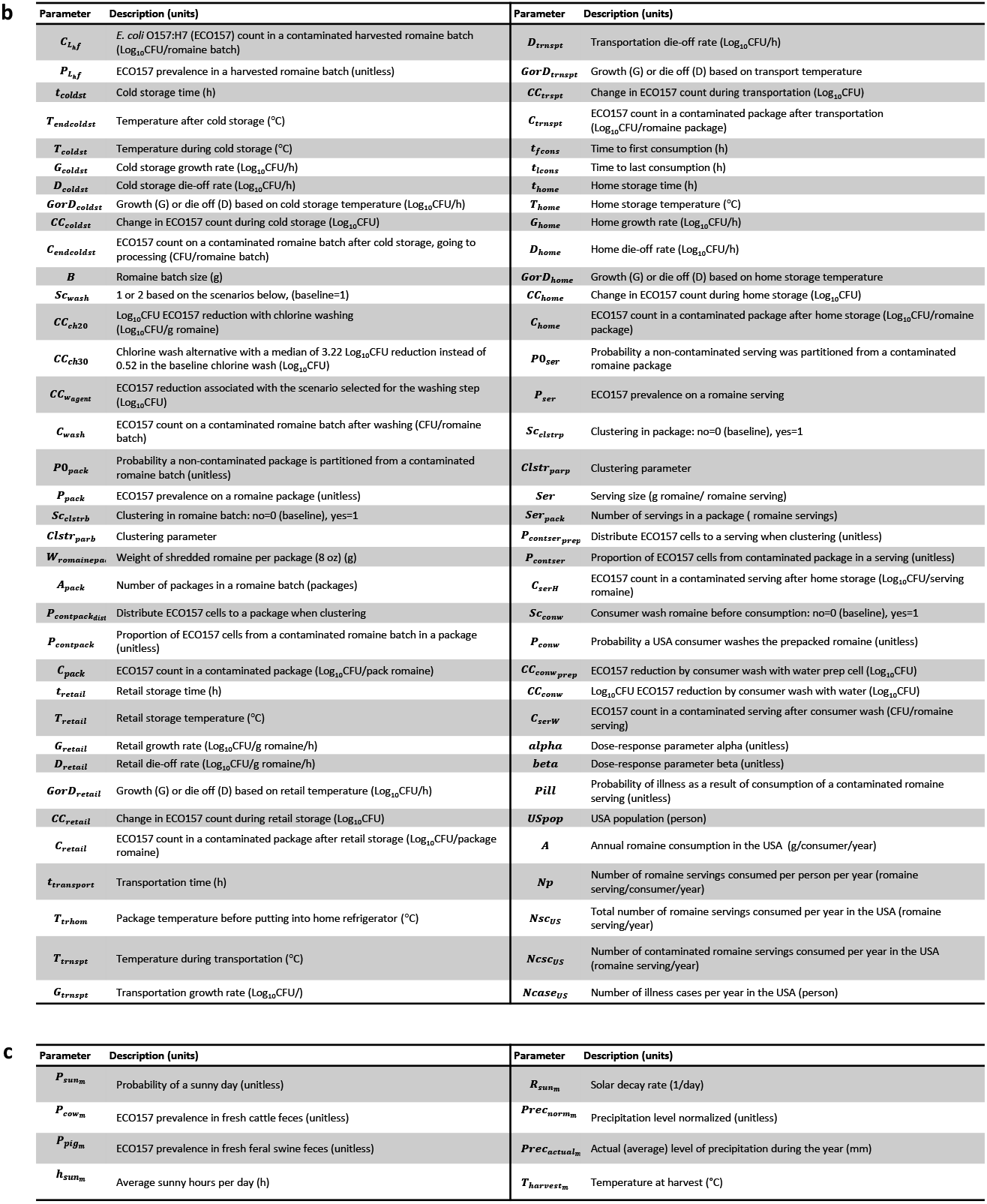
Parameter IDs and definitions used in **(a)** Preharvest model **(b)** Postharvest model; and **(c)** Parameters varying by the harvest month of romaine production

### Preharvest model

The preharvest model included growing and harvesting romaine in an open field. We considered a square field measuring approximately 63 m by 63 m (equivalent to 1-acre) for the cultivation of romaine. A total of 26,000 romaine plants were harvested from the modeled 1-acre field in a day. Each plant was measured 25cm in width, length and height, and weighed 300g *[51]*. The allocated area for each individual plant within the field was 30cm by 50cm *[36]*.

We assumed continuous romaine production throughout the year, with daily harvests, occurring in one of three distinct geographical locations: (i) the Coastal Region and Central Valley in CA (CC), (ii) the Desert Region (Imperial) in CA (DR), and (iii) the Yuma Region in AZ (Y). These locations were selected since they are the top leafy green producers in the USA (CA accounts for 70%, and AZ for 30% of US lettuce production) *[52-54]*. The choice of location affected several model parameters, such as the time from seed planting to harvest, and the type of irrigation systems used in the fields (Figure 2, Supplementary Tables S1, S2 and S3). Additionally, the month of harvest affected the parameters related to weather conditions and survival of ECO157 (Figure 2, Supplementary Table S3).

Contamination of romaine in the field before harvest was attributed to four sources based on the available information in the literature and the expert elicitation approach: (i) runoff from a neighboring dairy farm, (ii) intrusion by feral swine, (iii) irrigation water, and (iv) the application of BSAAO (i.e., treated/composted/processed dairy manure, which may include incompletely treated/composted/processed dairy manure). All calculations were conducted for a randomly selected 1/12 portion of the modeled field (i.e., 1/12 lot), which we refer to as the ‘batch area’, containing 2,125 romaine plants that weigh 637.5kg at harvest. The plants were grown and harvested as a batch unit and population dynamics of ECO157 were tracked for the batch. The size of the batch area enabled the development of a more realistic model by accounting for the risk factors associated with the exposure and spread of ECO157 throughout the farm-to-fork chain. For example, we assumed that only the batch area closest to the edge of the field (and thus to a hypothetical neighboring dairy farm) would be impacted by dairy farm runoff. Due to the rapid decay of ECO157 on field-grown romaine, we adopted a 14-day ‘countdown’ period, representing the final 14 days before harvest, during which we tracked the introduction of ECO157 to the romaine in the field *[39]*. The duration of this period was chosen assuming that furrow irrigation happened every 10 days on average and that an average ‘waiting time’ of 4 days was used before harvest (as explained in the later text, overhead spray and furrow irrigation cease during the waiting time). Throughout this countdown period, we tracked ECO157 counts on day t on both the romaine (*LN*_*t*+1_ ; CFU/romaine batch) and in the top 1cm layer of soil of the batch area (referred to as ‘soil batch’; *SN*_t+1_; CFU/soil batch) using a system of coupled difference equations (1) - (6) coded within the QMRA in @RISK (notations were explained in the following paragraphs and also in Supplementary Tables S4 and S5):

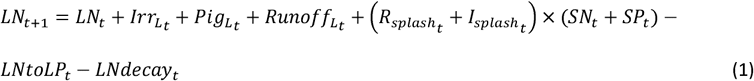

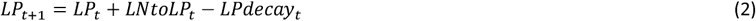

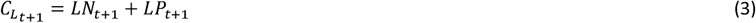

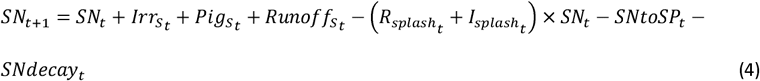

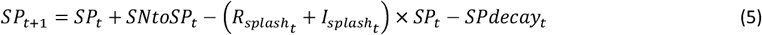

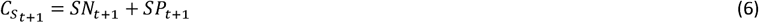

where *t* = 0,1, 2, 3, …, 13, 14 were days of the countdown period (i.e., at t=0 there were 14 days until harvest). If in a time step, the calculated *LN*_*t*+1_, *LP*_*t*+1_, *SN*_*t*+1_ or *SP*_*t*+1_ < 0, we set the corresponding *LN*_*t*+1_, *LP*_*t*+1_, *SN*_*t*+1_ or*SP*_*t*+1_ to 0.

We adopted the concept of non-persister and persister cell state dynamics to provide a more accurate die-off mechanism for ECO157 *[55]*, reflecting the well-known biphasic decay phenomenon (i.e., fast initial decay followed by slower decay) of the ECO157 cells on leafy green and in soil environments *[56-59]*. In Equation (1), *LN*_*t*+1_ represents non-persister ECO157 count in romaine batches on day *t*+1. It was determined by the previous day’s count in romaine batches (*LN*_*t*_); accumulation through irrigation 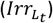, runoff 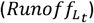, intrusion by feral swine 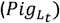, rain splash 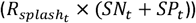 and irrigation splash 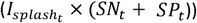 removal of cells that switched from non-persister to persister state (*LNtoLP*_*t*_) and daily reduction on romaine (*LNdecay*_*t*_). Equation (2) describes the persister ECO157 count in romaine batches on day *t*+1 (*LP*_*t+1*_). Total ECO157 count (non-persister and persister cells) in romaine batches on day *t*+1 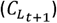 was calculated in Equation (3). Equations (4), (5) and (6) estimated the total ECO157 count (non-persister and persister) in the soil batch on day *t*+1 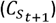 and were similar to the calculations for the corresponding romaine batch. It was assumed that the cross-contamination of ECO157 cells from soil to romaine via splash events triggers an adaptation process, resulting in a transformation of any transferred cells from a persister state to a non-persister state when they land on romaine leaves *[47]*.

The ECO157 counts in the romaine batch 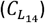 and soil batch 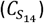 at *t*=14 were used in a harvest sub-model (explained below) to calculate the preharvest outcome: the ECO157 count in the harvested romaine batch 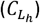. From there, the ECO157 prevalence 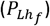 and the ECO157 count in harvested romaine batches 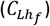 were calculated in one simulation (one simulation consists of 100,000 iterations) and used as inputs in the postharvest model. Specifically, 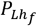 was derived by counting the number of iterations out of 100,000 where the ECO157 count on the harvested romaine batch was at least one (where 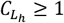CFU/romaine batch). The 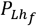 was established as a Triangular distribution, where the mean of the distribution corresponded to the predicted preharvest prevalence in the simulation with 100,000 iterations mentioned above. The distribution’s minimum and maximum were set at half and twice of the predicted prevalence, respectively. The 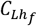 was derived by fitting a distribution around the predicted ECO157 counts in contaminated harvested romaine batches (where 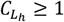 CFU/romaine batch) using @RISK software. The distribution that best fits the data was determined using Akaike Information Criterion, Bayesian Information Criterion and Average Log-Likelihood methods (Supplementary Table S2).

### Accumulation and loss of ECO157 contamination in soil batch (*c*_*s*_)

We assumed that irrigation water, BSAAO, swine, and cattle feces were the only sources of ECO157 contamination for the modeled field, explained in the following sections.

### BSAAO Application (accumulation in soil)

We modeled ECO157 prevalence (*P*_*man*_) and count (*c*_*man*_) in BSAAO by accounting for the reports of ECO157 in stacked/piled manure from dairy farms in the USA [60]. The maximum ECO157 count in BSAAO was the maximum ECO157 count found on raw dairy feces [36]. This represented an unintentional application of an incompletely treated BSAAO, though the application probability of such BSAAO was low. The timing of BSAAO application (*D*_*man*_) was set on the same day of planting romaine seeds. The number of days from planting romaine seeds to harvest (*Daytogrow*) was set between 65 and 130 days depending on the field location (CC, DR or Y regions) [48]. We considered a constant daily decay of ECO157 among both non-persister and persister cells in BSAAO (*R*_*dman*_ and 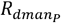), from the day of BSAAO application to the time of harvest [61,62]. A total of 10% of the simulated fields received BSAAO application (*Pr*_*man*_) (BSAAO may be incompletely treated), assuming that those were organic operations that used BSAAO as nitrogen source. The BSAAO application (*M*_*man*_) ranged from 5 to 15 tons per acre *[63]*. When applying BSAAO, we incorporated a dilution factor (*DiF*_*man*_) assuming a uniform distribution of BSAAO across the tilled column of the soil batch. This approach considered that a gram of BSAAO has a volume (*Rh*_*con*)_ of 2.5cm^3^ *[64]* and was assumed to be uniformly distributed across the topsoil (*Dh*_*con*_) up to 15 to 20cm depth *[65]*. In this and other instances of using a dilution factor in the soil column in our model, the assumption of a uniform distribution of ECO157 within the top layer of soil batch was justified since the unit of analysis of romaine contamination was the whole batch. For example, transfer of 100 ECO157 cells from one contaminated spot of soil onto a single romaine plant (which becomes contaminated with 100 ECO157) and transfer of 1 ECO157 cell from each of 100 different contaminated soil spots resulting in 100 plants contaminated with 1 ECO157 each, both resulted in the same contamination at the batch level (i.e., 100 ECO157 in a given romaine batch). The estimation of the initial ECO157 count in soil batch 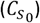 is provided in Supplementary Table S4.

### Runoff from a neighboring dairy farm (accumulation in soil) 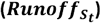

We adapted the runoff contamination component from Mishra et al.’s (2016) QMRA model [36]. Runoff events were considered to occur on rainy days only (at a probability 1-*Bs*_*t*_; a day could either be sunny or rainy). They lead to contamination of batch area at the edge of the field, determined randomly by the ‘*edge*’ parameter. We assumed that 5 to 15kg of dairy cattle feces (*M*_*cow*_) could enter the batch area during a runoff event, where the quantity of fecal material entering the batch area was proportional to the monthly average precipitation of the field location 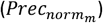. We incorporated ECO157 count (*C*_*cow*_) and month-specific ECO157 prevalence (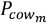, Supplementary Table S3) in dairy cattle feces from Cooley et al. (2013) *[8]*. The spread of runoff across the *edge* batch area affected both romaine batch and soil batch, determined by the ‘*Pr*_*Ltos*_’ parameter. This parameter was a proportion describing the area a romaine plant occupies relative to the soil area allocated for plant growth. The runoff in soil was uniformly distributed and spread into the soil. The ECO157 in runoff was spread to a soil depth of 1cm by a dilution factor (*DiF*_*cow*_). The estimation of the term 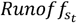 in the model (1) - (6); i.e., the ECO157 count in soil batch added via runoff on day t, is provided in Supplementary Table S5.

### Intrusion by feral swine (accumulation in soil)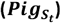

Wildlife intrusion in our model was integrated using the risk model developed by Mishra et al. (2016) [36]. A contamination event was dependent on the probability of feral swine intruding the field 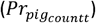, derived from reported feral swine population density in Salinas Valley, CA (∼ 5 swine/km^2^) *[6]*. In the event of an intrusion, we assumed that between 1-5 adult swine would enter the field and defecate, as they could be found individually and in family groups in CA *[66]*. In 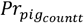, the probability of intrusion by one animal was the highest, and the probability of intrusion by more animals decreased progressively. The fecal quantity found on the field after the defecations (*M*_*pig*_) was estimated by the daily fecal production reported per adult swine (*Mdef* _*pig*_) *[67]*. In addition, we factored in a defecation frequency of 4 times/swine/day (*Tdef* _*pig*_) *[68]* and assumed at least one of those defecation events occurred in the modeled field (*Ndef* _*pig*_). Feral swine could defecate on romaine plants or field soil, determined by the ‘*Pr* _*Ltos*_’ parameter. When feces fall on soil, we assumed the ECO157 in fecal material was spread evenly within the top 1cm of the soil batch, which we modeled using a dilution factor (*DiF* _*pig*_). The ECO157 count (*C*_*pig*_) and the ECO157 prevalence 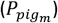 in swine feces were modeled based on the data provided by Cooley et al. (2013) [8]. The term 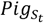 in the model (1) - (6); i.e., ECO157 count in soil batch added via feral swine on day t, was estimated as shown in Supplementary Table S5.

### Irrigation (accumulation in soil)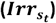

Contamination of soil by irrigation occurred through contaminated water that was delivered to the field and not captured by the romaine plants on the field. The ECO157 prevalence 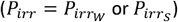 and count (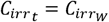 or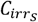) in the irrigation water depended on the water source: surface or well (ground) waters. Water source was selected based on its respective frequency of use in a field location [69]. We assumed the value for the ECO157 prevalence in well water 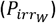 as 0.1%, given the limited literature on well waters, and considering that well waters are generally free of ECO157. Our scenario analysis in Supplementary Table S6 showed that decreasing this prevalence tenfold— to 0.01%—resulted in a 20.7% decrease in predicted illness risk compared to the baseline model. This finding suggests that ECO157 levels in well waters could have a modest yet noteworthy impact on lettuce safety. ECO157 prevalence in surface water 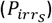 was based on the ECO157 survey on surface waters in Monterey County, CA [8]. The ECO157 counts in surface water included two parts: (i) the generic *E. coli* count and (ii) a factor that converts the generic *E. coli* count to that of ECO157 *[34]*. The ECO157 count in well water 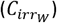 was the LGMA testing limit of generic *E. coli* for Type A agricultural waters, set at 10MPN/100ml *[19,20]*. For consistency across model calculations, we assumed that the most probable number (MPN) was equivalent to colony forming units (CFU) *[70]*. The generic *E. coli* count part of the surface water 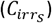 was represented as a PERT distribution, with the minimum and most likely values taken from the standard testing limits set by the LGMAs for Type B agricultural waters used in drip and furrow irrigation systems *[19,20]*. The maximum value of the PERT distribution was based on the highest generic *E. coli* count observed in surface waters from CA region *[71]*. Starting in 2023, the LGMAs implemented more stringent testing limits for overhead spray irrigated surface waters, setting the threshold at 235MPN/100ml generic *E. coli* for any single sample. However, we applied the less stringent testing limit historically used by LGMA (576MPN/100ml) in the baseline model to account for agricultural waters in poorer conditions and to reflect the practices more relevant to the period for which the model validation data were available. However, our scenario analysis in Supplementary Table S6 predicted that adopting the more stringent testing limit led to 31.1% reduction in predicted illness risk compared to our baseline model, suggesting that the 2023 California and Arizona LGMAs may have a positive impact on leafy green safety. Our model included different irrigation systems (*f*_*irr*_ = overhead spray (O), furrow (F), or drip (D)) based on the field location (Figure 2, Supplementary Table S1). Each system delivered varying volumes of irrigation water per irrigation event (*g*_*irr*_), considering that most of the water (i.e., 51% of the total volume required for romaine growth) was applied in the last 30 days before harvest (*w*_*O*_, *w*_*F*_, *w*_*D*_) [48]. The volume of water captured by a romaine batch (*v*_*irr*_) also differed for each system [27,72]. Overhead spray and furrow irrigation only occurred on sunny days (*Bs*_*t*_) [73], and differed in frequency: Overhead spray, applied once every 2-3 days, was reflected in probability *Pr*_*O*_ (*fO*_*t*_ = *Binomial* 1, *Pr*_*O*_)) and furrow was applied about once every 10 days, reflected in probability *Pr*_*F*_ (*fF*_*t*_ *= Binomial* (1, *Pr*_*F*_). Drip irrigation occurred every day 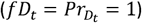 regardless of sunny or rainy weather. We used a dilution factor (*DiF*_*irr*_) to simulate an even distribution of ECO157 introduced by irrigation across the field and spread of cells to a depth of 1cm. To align with typical harvest practices in the USA *[74]*, we introduced a waiting period ranging from 2 to 8 days before harvest (*BwaitP*_*t*_) and terminated overhead spray and furrow irrigation during the selected waiting time. Shallowly buried drip lines were assumed for drip-irrigated fields, making them unaffected by the waiting period. The term 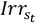 in the model (1) - (6), the ECO157 count in soil batch added via irrigation water on day t, is provided in Supplementary Table S5.

### Soil splash via rain or irrigation (i.e., loss from soil) 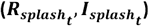

The effect of irrigation and rain splash in the field was based on the QMRA model developed by Allende et al. (2018) *[27]*. Accordingly, splash of soil (and ECO157) to romaine would be facilitated either via irrigation water on a sunny day or rainwater on a rainy day. The maximum quantity of soil splashed by rain was assumed to be equal to that splashed by overhead spray irrigation. However, the quantity of soil splashed by rain was also proportional to the standardized monthly precipitation level 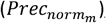, reflecting the intensity of rain. We assumed that the quantity of soil splashed on sunny days varied by different irrigation systems, with overhead spray irrigation had the potential of causing the most amount soil to splash and drip irrigation causing the least (*M*_*irr*_). The amount of soil splashed during irrigation was also affected by the intensity of irrigation systems, where each system assumed to mimic an intermediate rainfall, equal to the median of the standardized precipitation 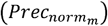. In case of a splash, a fraction of ECO157 in soil would transfer onto the romaine (*LsR* during rain or *LsI* during irrigation, Supplementary Table S4) via transfer of contaminated soil [75]. The terms for modeling rain 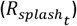 and irrigation splash 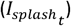 in the model (1) - (6) were derived as shown in Supplementary Table S5.

### Daily decay of ECO157 (loss from soil) (*SNdecay*_*t*_, *SPdecay*_*t*_)

Daily inactivation of persister and non-persister cells in soil was constant. We used four different decay rates to represent non-persister *[36,61]* and persisters’ *[62]* decay in BSAAO-amended soil and soil with no BSAAO application. We used slower inactivation in BSAAO amended soil (*R*_*dman*_ for non-persisters and 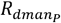 for persisters) and faster inactivation in soil with no BSAAO application (*R*_*ds*_for non-persisters and 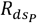for persisters). Persister cells decayed at a rate 10 times slower than that of their non-persister counterparts, based on the decay curve provided in Chen (2020) for ECO157 persisters in soil *[62]*. Decay parameters in soil (*SNdecay*_*t*_, *SNdecay*_*t*_) in the model (1) - (6) are explained in Supplementary Table S5.

### Switch to persister cells (no change in the cell count in soil) (*SNtoSP*_*t*_)

A small fraction of non-persister ECO157 cells were transitioned to persister cells on each day in soil *[47,76,77]*. Persisters coexisted with non-persisters and comprised up to 0.01% of the population in soil environments *[76,77]*. The cells could also switch between the persister and non-persister states *[78]*. Although the characteristics and the formation mechanisms of persisters are complex *[78]*, our model focused only on their enhanced ability to survive, which facilitated modeling biphasic decay. While the direct estimates for the switch rates in BSAAO-amended soil were unavailable, we used the study by Hofstrenge et al. (2013) *[77]*, which indicated a strong one-to-one correlation between switch rates (*NtoP*_*man*_, *NtoP*_*s*_) and the fraction of persisters (*Pmax*_*man*_, *Pmax*_*s*_). We assumed that the persister state was more prevalent in BSAAO-amended soil batches (*NtoP*_*man*_). This assumption was based on the observation that pathogens regrew and recovered to a culturable state months after being introduced into soils through dairy BSAAO application [79]. This delayed recovery might suggest the initial presence of persister cells within the BSAAO that was applied on the field [80]. Details on the switch terms (*SNtoSP*_*t*_) in the model (1) - (6) are provided in Supplementary Table S5.

### Accumulation and loss of ECO157 in romaine batches

Given the rapid decay of ECO157 on field romaine *[27,39,81]*, we assumed that the initial ECO157 count on the field romaine was negligible one day prior to the start of the 14-day countdown period 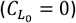. However, the sensitivity of model predictions to this assumption was tested in scenario analysis (see Supplementary Table S7).

### Irrigation (accumulation on romaine)

Contamination of romaine through irrigation followed a similar mechanism to that described for soil contamination. The volume of irrigation water captured by the romaine batches (*v*_*irr*_) transferred a fraction of ECO157 cells from water to the romaine plants (*Tr*_*ecirr*_). The total volume of water delivered to the batch area (*g*_*irr*_) was either captured by romaine or spread into the soil. Thus, we modelled the fraction of irrigation water delivered to the soil 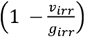 and romaine 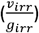. The term 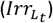 describing the irrigation contamination in the model (1) - (6) is explained in Supplementary Table S5.

### Runoff from a neighboring dairy farm and intrusion by feral swine (accumulation on romaine)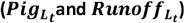

The effect of wildlife and runoff contamination on romaine was modelled mechanistically similar to their effect on soil detailed previously. The difference was the proportion of feces/runoff that was spread in either the soil or on romaine, as determined by the parameter ‘*Pr*_*Ltos*_’ (for soil: 1-*Pr*_*pigLtos*_, and for romaine: *Pr*_*Ltos*_). The feces/runoff was modelled as spreading uniformly through the romaine batches. The terms 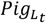 and 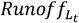 describing the ECO157 count in romaine batches via feral swine intrusion and runoff events in the model (1) - (6) are explained in Supplementary Table S5.

### Soil splash via rain or irrigation (accumulation on romaine)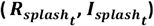

The daily loss of ECO157 from the soil due to rain or irrigation splash was applied to a given romaine batch, resulting in ECO157 accumulation on romaine batches. The transferred ECO157 cells were distributed uniformly in a given romaine batch (Supplementary Table S5).

### Daily decay of ECO157 (loss from romaine) (*LNdecay*_*t*_, *LPdecay*_*t*_)

For non-persister ECO157 decay on romaine (*LNdecay*_*t*_, Supplementary Table S5), the daily decay rate (*R*_*dL*_) followed the month-specific solar decay pattern 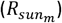 throughout the year *[27]*. Specifically, a slower decay rate, reflecting winter conditions, was applied from December to April (for production in Y and DR regions), while a faster decay rate, reflecting summer conditions, was implemented from May to October (for production in Y region). Additionally, the parameter’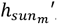, which indicated the average daily sunlight hours in a given month, was determined based on weather data specific to both field location and the time of romaine harvest. The decay for persisters 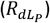 was assumed 10 times slower than the non-persisters (*LPdecay*_*t*_, Supplementary Table S5) *[47,62]*.

### Switch to persister cells (no change in the cell count on romaine) (*LNtoLP*)

The switch rate from non-persister to persister cells on romaine (*LNtoLP*_*t*_, Supplementary Table S5) was constant and established based on the data from the study by Munther et al. (2020) *[47]*.

### Harvest (*cc*_*blade*_)

After modeling the population dynamics of ECO157 in soil and on romaine plants during the 14-day countdown period (equations (1) - (6)), we proceeded with modeling the harvesting process, which accounted for cross-contamination of romaine from soil caused by contaminated coring knife blades [15,38].

### Postharvest model

The bacterial population dynamics outlined by Pang et al. (2017), Danyluk and Schaffner (2011) and Nauta (2005) served as the baseline for the postharvest practices (i.e., cold storage prior to processing, chlorine wash, retail, transportation and consumer stages) *[32,38,46]*.

### Cold storage before chlorine wash (*CC*_*coldst*_)

The harvested romaine was transported to cold storage, where it was kept for a period ranging from 1 to 72 hours (*t*_*coldst*_). In this stage, we used 5°C as the minimum temperature for ECO157 growth. Time spent (*t*_*coldst*_) above 5°C increased ECO157 counts (growth rate, *G*_*coldst*_) and time spent ≤ 5°C decreased ECO157 counts (decay rate, *D*_*coldst*_) [38,82]. Cold storage temperature was assumed to follow a triangular distribution centered around the standard refrigeration temperature, allowing inadequate storage (*T*_*endcoldst*_). We assumed that the temperature of romaine at the time of harvest (*T*_*harvest*_) was equivalent to the air temperature during the harvesting process, under the assumption that harvesting occurs early in the morning *[83]*. Accordingly, we used field location- and month-specific weather data for determining the harvesting temperature. *CC*_*coldst*_ in Supplementary Table S2 represented the change in ECO157 count on a given romaine batch during cold storage.

### Chlorine wash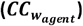

The washing step was assumed to reduce ECO157 count by 0.52 Log_10_CFU/g romaine on average, with a range of reduction between 0.01 Log_10_CFU/g romaine and 1 Log_10_CFU/g romaine (Supplementary Table S2) *[84]*.

### Partitioning of the romaine batches into packages (*Sc*_*clstrb*_, *Cluster*_*parb*_)

The partitioning of a chlorine-washed romaine batches (*C*_*wash*_) into individual packages included potential clustering of ECO157 cells (or aggregation of cells) *[46,85]*. To estimate the ECO157 count within a contaminated romaine package (*C*_*pack*_), we used the following equations:

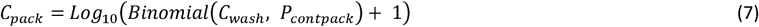

where

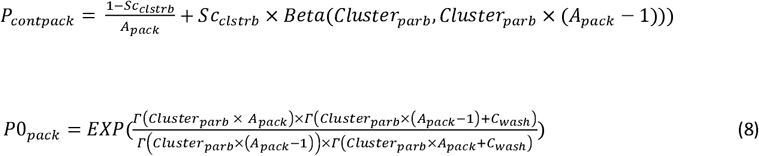

Here, *P*_*contpack*_ is an intermediary parameter used to determine the ECO157 count within a given package, accounting for the presence of clustering (*Sc*_*clstrb*_; set to 0 when clustering was absent and 1 when present) and the degree of clustering when it occurred (with maximum clustering when *cluster*_*parp*_ = 0, decreasing as *cluster*_*parp*_ = 0 approaches positive infinity). In the baseline model, we assumed no clustering (*Sc*_*clstrb*_ = 0), and as a result, the ECO157 cells from the romaine batches were evenly distributed to each package 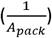. The ‘*P0*_*pack*_*’* parameter in Supplementary Table S2 (equation (8)) determined the probability of an uncontaminated package *[46]*. We derived the probability of a package being contaminated as (1-P0_pack_).

### Retail storage, transport from retail to home, and consumer (*CC*_*retail*_, *CC*_*trspt*_, *CC*_*home*_)

We used the same growth and decay approach implemented in cold storage process to model retail storage, transport from retail to home, and consumer steps (i.e., home storage) albeit with stage specific time and temperature parameters *[38,81]*. The values of the time and temperature parameters were previously parameterized by Pang et al. (2017) *[38]*.

### Partitioning of the romaine package into servings (*Sc*_*clstrp*_, *cluster*_*parp*_)

We adopted a serving size of 85g for romaine (*ser*) based on reference serving size provided by USDA (2015) *[86]*. To distribute ECO157 cells from romaine packages to servings, we followed the same approach used for partitioning romaine batches to packages (no partitioning at the baseline model, Equations (7) - (8)) *[46]*.

### Dose-response

Following the partitioning process, the ECO157 count (*C*_*serW*_) and prevalence (*P*_*ser*_) in romaine servings were estimated. The *c*_*serW*_ was used in a Beta-Poisson dose-response model (*Pill*) (Supplementary Table S2) *[32,87]*. We estimated our main outcome, the predicted number of illness cases per year in the USA from consuming ECO157-contaminated romaine servings (*Ncase* _*US*_), by using the dose-response model along with the number of contaminated servings consumed annually in the USA (*Ncsc*_*US*_).

### Expert elicitation

Several parameters across the farm-to-fork chain, as well as specific interventions for the scenario analysis, were determined by using the input from study authors and industry experts who served on an advisory council for the USDA-NIFA-SCRI grant #2019-51181-30016. Three informal interviews were conducted with the advisory council members, who were from produce production and processing, distribution and retail, and sanitation industries. Additionally, an online survey was created in Qualtrics (Provo, UT, USA) and administered to the advisory council (12 respondents) to identify the intervention strategies of greatest interest to industry. Based on the feedback, (i) cattle vaccination, (ii) transition from overhead spray and furrow to drip irrigation, and (iii) transition from overhead spray to furrow irrigation, were the most requested intervention strategies and were included in the scenario analysis. Further, in another online survey with the council conducted via Qualtrics, we presented the scope of the risk assessment model, including contamination sources and interventions.

### Model validation

The main model outcome was validated with the information from the CDC for the year 2011 (most recent data). Since the most recent data was from 2011, we used the 2011 USA population data (*USpop*) *[88]* along with 2011 USA fresh romaine and leaf lettuce consumption data (consumer weight without accounting for the food loss at consumer level) *[18]*. The validation estimated the USA foodborne diseases associated with leafy greens contaminated with ECO157 *[49]*. Accordingly, the estimated number of foodborne diseases due to ECO157 contamination in the USA averages 63,153 cases annually, with 90% credible interval from 17,587 to 149,631. Of these, 19.3% - 31.5% are attributed to leafy greens *[50]*, resulting in an average of 12,189 – 19,893 cases each year in the USA linked to consuming contaminated leafy greens. Considering the 90% credible interval, the true range of annual illness cases attributable to consumption of romaine could be between 3,394 and 47,134.

### Sensitivity and uncertainty analysis

We evaluated the effect of variable and uncertain parameters in the baseline model to determine the key factors influencing the main outcome (the predicted number of illnesses in the USA). We used @RISK and R (*dplyr* package) for the sensitivity and uncertainty analyses (for preharvest and postharvest models) and generated tornado plots of Spearman rank-order correlation coefficients (SRCCs) *[89]*. The SRCC analysis for the baseline preharvest model was based on a set of core and derived parameters, where derived parameters were calculated from core parameters. However, no parameter was included in the analysis more than once. This approach was taken to ensure that the results were more interpretable and informative. More information about the selection of parameters is provided in Supplementary Table S8. Parameters with absolute value of the SRCC |ρ| < 0.05 were considered negligible and were excluded from the tornado plots. Likewise, only parameters with statistically significant SRCC were presented in graphs, adhering to a Bonferroni-corrected alpha threshold of 0.05 divided by the number of evaluated parameters (Sensitivity analysis: 21 for preharvest, 18 for postharvest; uncertainty analysis: 11 for preharvest). The absolute value of the SRCC, or |ρ|, was interpreted according to its common usage in the literature (ρ=0 to <0.1, none; ρ=0.1 to <0.3, poor; ρ=0.3 to <0.6, fair; ρ=0.6 to <0.8, moderate; ρ=0.8 to 1.0 very strong correlation) *[90,91]*. In the sensitivity analysis of variable parameters, all uncertain parameters were set to their fixed deterministic values. A parameter was classified as an uncertainty parameter when no information was available in the literature, requiring assumptions. In the uncertainty analysis of the preharvest model, we fixed the variable parameters to the median values of their respective distributions and created a distribution for each of the uncertain parameters by establishing a triangular distribution. The minimum of each triangular distribution was set as a 50% reduction from the baseline, and the maximum was set as a 50% increase from the baseline for all uncertain parameters, with the original baseline value as the mean/mode. In the preharvest model, this definition applied to the following parameters: (i) volume of irrigation water captured by romaine through furrow or drip irrigation per irrigation event (*V*_*F*_, *V*_*D*_); (ii) fraction of soil batch transferred to romaine batches via splash caused by furrow or drip irrigation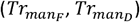); (iii) dilution factors used for the distribution of irrigation water, feces and runoff in soil batch (*DiF*_*irr*_, *DiF*_*pig*_, *DiF*_*cow*_); (iv) switch rate from non-persister to persister cells in BSAAO-amended soil (*NtoP*_*man*_); (v) ECO157 prevalence 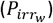 in well water; and (vi) transfer of ECO157 cells from irrigation water to the romaine batch in an irrigation event (*Tr*_*ecirr*_).

### Regression trees

Decision trees are developed for both preharvest and postharvest models using the rpart package in R *[92]*. All analyses were implemented at default settings in rpart.control function. To prevent tree overfitting, we used 10-fold cross-validation *[81]*. Similar to the SRCC analysis, the regression tree for the preharvest model was based on the set of the core and derived parameters previously determined (Supplementary Table S8).

### Scenario analysis of risk management strategies

Based on the sensitivity and uncertainty analyses, as well as the results of expert elicitation, we identified nine risk management strategies for evaluation in the scenario analysis (Table 1, Supplementary Table S9). These scenarios are (i) cattle vaccination, (ii - iv) treatment of surface water, used for overhead spray irrigation, by either chlorine, peracetic acid or ultraviolet radiation, (v) transition from overhead spray to furrow irrigation system, (vi) transition from overhead spray and furrow to drip irrigation, (vii) chlorine wash alternatives during postharvest processing, (viii) temperature reduction during retail, and (ix) consumer wash (the modifications to the parameters to apply to these scenarios were provided in Table 1). In addition, we evaluated the effect of parameters related to clustering of ECO157 cells during the partitioning process (i.e., romaine batch to package and package to serving) as an additional scenario (Table 1). The effect of each risk management strategy or clustering was assessed based on their impact on the main outcome.

**Table 1.** Description of the scenarios and changes in the illness outcome for potential risk management strategies.

| Scenario | Definition (Reference) | ID of parameters affected (unit) | Parameter value in the scenario | Predicted illness cases resulted from the tested scenario: 5<sup>th</sup>; 50<sup>th</sup>; 95<sup>th</sup> percentile |
| --- | --- | --- | --- | --- |
| Baseline | No changes made in the model. | None | All parameters were at their baseline conditions. | 42; 19,040; $3.6 \times 10^6$ |
| Cattle vaccination | ECO157 counts were reduced by $1 - \log_{10}$ CFU/g in BSAAO and cattle feces [93]. | $C_{man}$ ($\log_{10}$ CFU/g BSAAO) and $C_{cow}$ ($\log_{10}$ CFU/g feces) | $C_{man} = \text{Uniform}(3.1, 8.4) - 1$<br>$C_{cow} = \text{Pert}(0.89, 3.08, 8.4) - 1$ | 43; 14,113; $2.6 \times 10^6$<br>Median number of cases reduced by 25.9%. |
| Surface water treatment for overhead spray irrigation by chlorine | ECO157 counts in surface water were reduced when chlorine was applied [94]. | $C_{irr_s}$ (CFU/ml) | $\frac{C_{irr_s}}{10^{\text{Uniform}(1.00, 2.78)}}$ | 27; 1,803; $4.7 \times 10^5$<br>Median number of cases reduced by 90.5%. |
| Surface water treatment for overhead spray irrigation by peracetic acid (PAA) | ECO157 counts in surface water were reduced when PAA was applied [94,95]. | $C_{irr_s}$ (CFU/ml) | $\frac{C_{irr_s}}{10^{\text{Uniform}(2.48, 3.09)}}$ | 20; 608; $2.7 \times 10^5$<br>Median number of cases reduced by 96.8%. |
| Surface water treatment for overhead spray irrigation by Ultraviolet radiation (UV) | ECO157 counts in surface water were reduced when UV was applied [94,96]. | $C_{irr_s}$ (CFU/ml) | $\frac{C_{irr_s}}{10^{\text{Uniform}(1.30, 2.91)}}$ | 27; 1,705; $4.4 \times 10^5$<br>Median number of cases reduced by 91.1%. |
| Transition from overhead spray to furrow irrigation (i.e., all irrigation was through either furrow or drip) | All overhead spray irrigated fields were transitioned to furrow irrigated fields. | $Prp_O$ and $Prp_F$ (unitless) | $Prp_O = CC: 0.0; DR: 0.0; Y: 0.0$<br>$Prp_F = CC: 0.4; DR: 0.7; Y: 1.0$ | 12; 715; $6.3 \times 10^5$<br>Median number of cases reduced by 96.3%. |
| Transition from overhead spray and furrow to drip irrigation (all irrigation was through drip) | All overhead spray and furrow irrigated fields were transitioned to drip irrigated fields. | $Prp_O$, $Prp_F$ and $Prp_D$ (unitless) | $Prp_O$ and $Prp_F = CC: 0.0; DR: 0.0; Y: 0.0$<br>$Prp_D = CC: 1.0; DR: 1.0; Y: 1.0$ | 14; 706; $5.3 \times 10^5$<br>Median number of cases reduced by 96.3%. |
| Produce wash alternatives with varying $\log_{10}$CFU reductions | Alternative produce washes at postharvest with varying reductions were used instead of the baseline chlorine wash (all scenarios, except $CC_{ch5}$ [97] are based on assumptions). | $CC_{ch20}$ ($\log_{10}$CFU) | $CC_{ch20} = Pert(0.01, 0.52, 1.03)$ was changed to $CC_{ch1} = Pert(0.5, 1.0, 1.5)$ | 31; 7,251; $1.2 \times 10^6$<br>Median number of cases reduced by 61.9%. |
| | | | $CC_{ch2} = Pert(1.0, 1.5, 2.0)$ | 25; 2,426; $4.1 \times 10^5$<br>Median number of cases reduced by 87.3%. |
| | | | $CC_{ch3} = Pert(1.5, 2.0, 2.5)$ | 22; 829; $1.4 \times 10^5$<br>Median number of cases reduced by 95.7%. |
| | | | $CC_{ch4} = Pert(2.0, 2.5, 3.0)$ | 21; 307; $5.3 \times 10^4$<br>Median number of cases reduced by 98.4%. |
| | | | $CC_{ch5} = Uniform(3.0, 5.0)$ | 18; 56; $5.3 \times 10^3$<br>Median number of cases reduced by 99.7%. |
| Temperature reduction during retail | Temperature distribution was changed [98,99]. | $T_{retail}$ ($^{\circ}\text{C}$) | $T_{retail} = Normal(4.4441, 2.9642, Truncate(0, 20.56))$ was changed to $Uniform(0, 4.4)$ | 38; 15,792; $5.5 \times 10^5$<br>Median number of cases reduced by 17.1%. |
| Consumer wash | 62% of consumers washed the product (yielding a median of 0.99 $\log_{10}$ CFU ECO157 reduction before consumption) [100]. | $CC_{conw}$ (unitless) | $CC_{conw} = 0$ was changed to $Triang(0.65, 0.99, 0.99) \times 0.62$ | 32; 13,717; $1.2 \times 10^6$<br>Median number of cases reduced by 28.0%. |
| Clustering effect | Clustering was allowed during partitioning of a romaine batch to packages and package to servings. Smaller $Clstr_{parb}$ or $Clstr_{parp}$ values indicated higher clustering of cells [46,85]. | $Clstr_{parb}$ (unitless), $Clstr_{parp}$ (unitless) | $Clstr_{parb} = 0.1$<br>$Clstr_{parp} = 0.1$ | 38; 9,862; $2.8 \times 10^5$<br>Median number of cases reduced by 48.2%. |
| | | | $Clstr_{parb} = 1$<br>$Clstr_{parp} = 1$ | 44; 20,103; $2.9 \times 10^6$<br>Median number of cases increased by 5.6%. |

## Results

### Preharvest model

Under the baseline model conditions, our model predicted a 1.00% prevalence of ECO157 in romaine batches across all three regions. The median ECO157 count in contaminated romaine batches (637.5kg romaine/batch or 2,125 plants/batch) after harvest 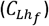 was 3.30 Log_10_CFU/romaine batch (5^th^ – 95^th^ percentile: 0.59 – 5.41).

The sensitivity analysis of the variable parameters in the preharvest model (Figure 3), determined by SRCC, indicated that the ECO157 count in irrigation water 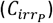 has a fair-strength (0.29) positive correlation with the ECO157 count in romaine batches (outcome of preharvest). The rest of the parameters were either poorly correlated or showed no correlation.

**Figure 3.**
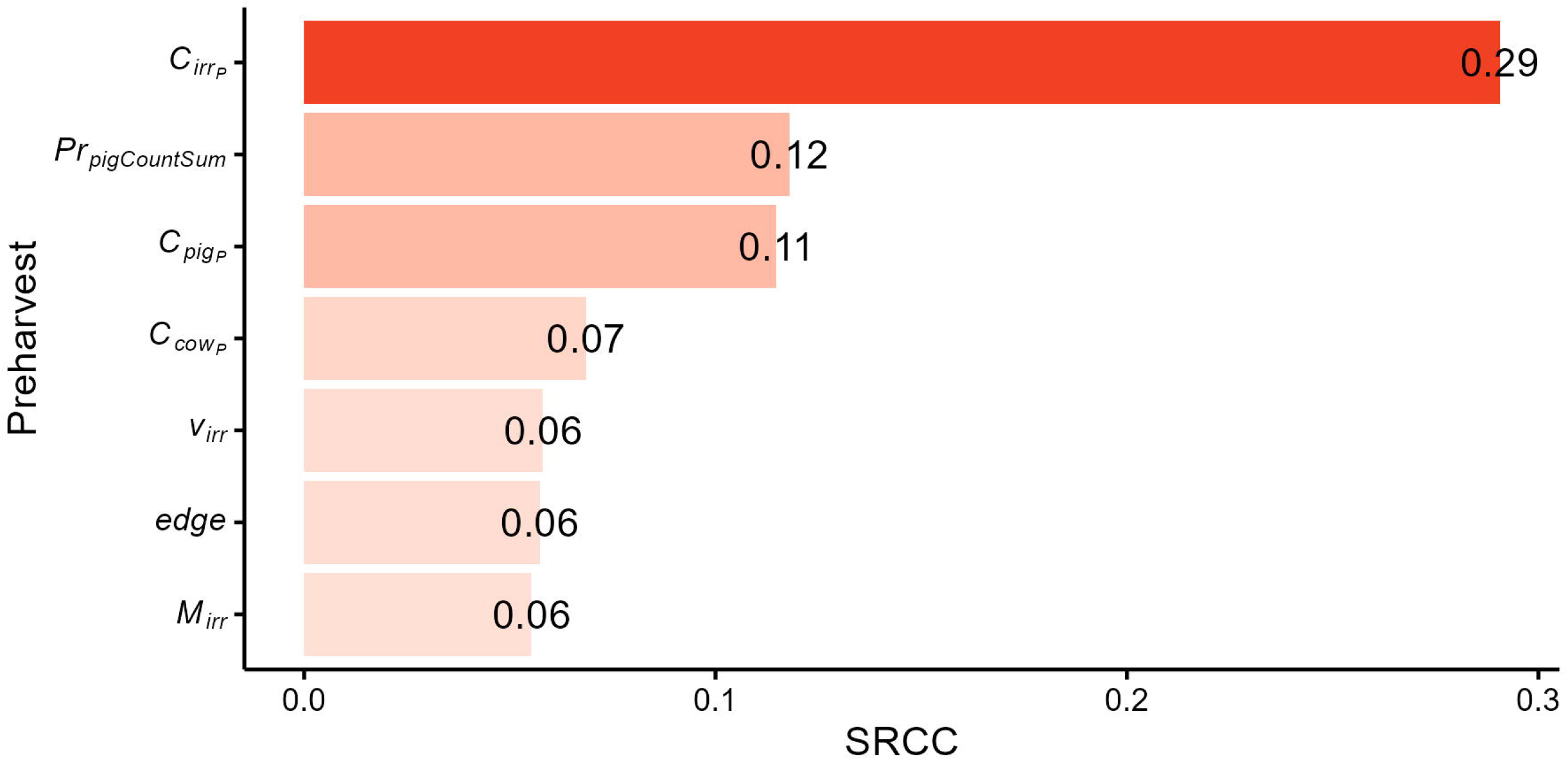
Tornado plot showing the impact of variable parameters on the preharvest output: the predicted *E. coli* O157:H7 (ECO157) count on romaine batches after harvest, ready for processing. The Spearman rank-order correlation coefficients (SRCC) are shown next to each parameter bar. Only variables statistically significant after the Bonferroni correction and with a correlation coefficient > 0.05 are shown. All values are positive, indicating positive correlation (i.e., an increase in each parameter value increases ECO157 count in romaine batches).

To assess the impact of preharvest contamination sources (irrigation, wildlife, runoff, and BSAAO) on the ECO157 counts in romaine batches, we ran the baseline preharvest model separately for each source and visualized their effects using violin plots (i.e., we evaluated the effect of each source individually, excluding the effect of the other three sources). Figure 4 (a) shows the distribution of ECO157 counts in contaminated romaine batches (i.e., with ECO157 counts ≥1 CFU/romaine batch), for each preharvest source, including the median, interquartile range, and 5^th^ to 95^th^ percentile across 100,000 iterations. Irrigation was identified as the primary contributor to ECO157 counts in romaine batches (Figure 4 (b)) with a median ECO157 count of 3.91 Log_10_CFU/romaine batch (5^th^ – 95^th^ percentile: 0.00 – 5.29) and an ECO157 prevalence of 0.53%. This was followed by wildlife, runoff and BSAAO contamination sources (Figure 4 (b)). While wildlife (median ECO157 count: 2.27 Log_10_CFU/romaine batch, 5^th^ – 95^th^ percentile: 0.47 – 4.47) and runoff (median ECO157 count: 2.28 Log_10_CFU/romaine batch, 5^th^ – 95^th^ percentile: 0.48 – 5.28) both led to similar median ECO157 counts when contamination occurred, contamination by wildlife intrusion events was more frequent (0.26%) than runoff contamination (0.21%). The predicted ECO157 count in romaine batches contributed by BSAAO contamination was low and occurred in 2 out of 100,000 batches (0.002% occurrence), with ECO157 counts of 8 and 16 CFU/romaine batch. This result on BSAAO contamination was not negligible given the volume of romaine production in the USA: Between 2018 and 2022, an average of 3,578 million kilograms of lettuce were produced annually, with romaine and leaf lettuce together accounting for 53% of the total production *[101]*.

**Figure 4.**
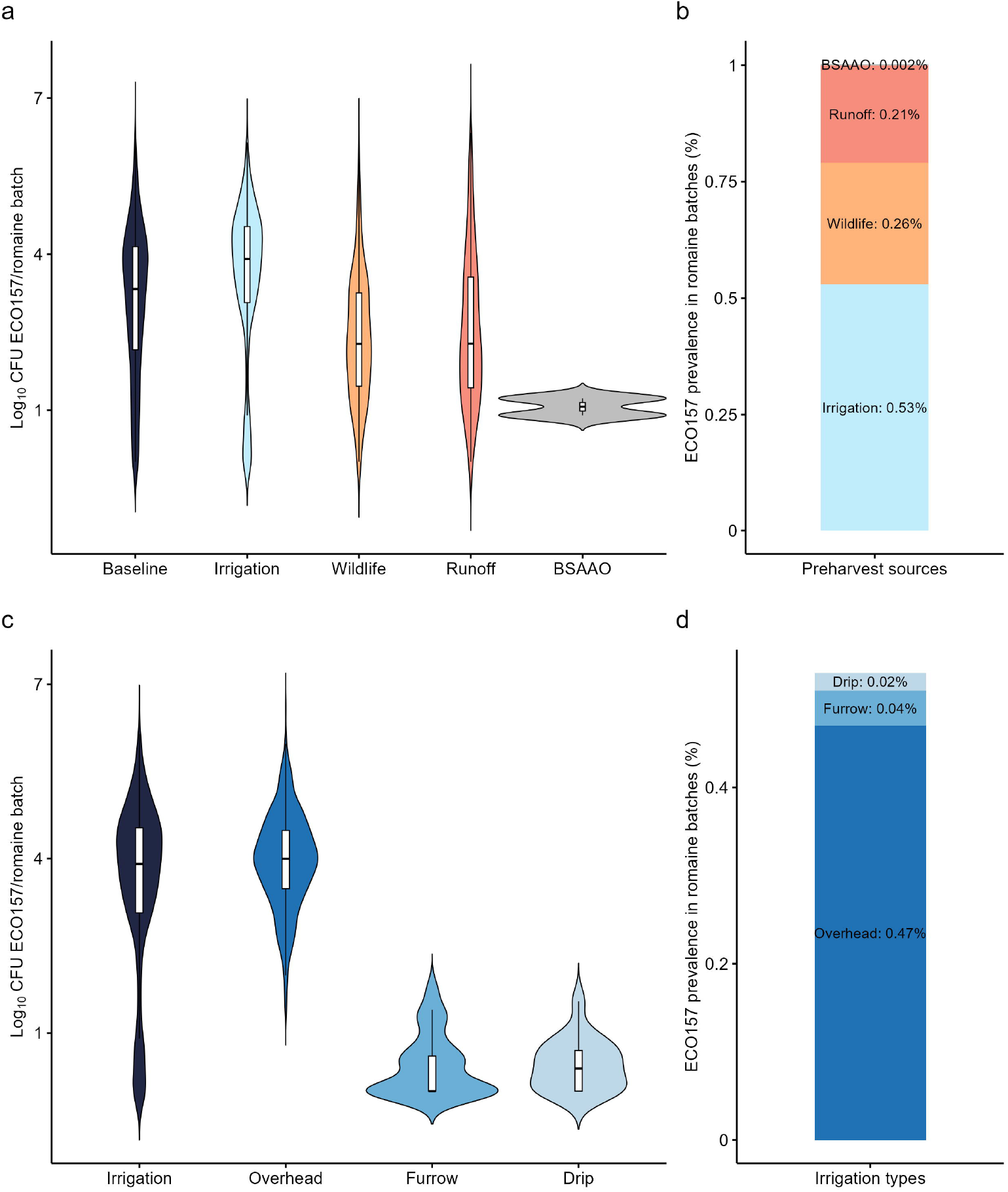
*Escherichia coli* O157:H7 (ECO157) counts in contaminated romaine batches in the baseline model (with all four sources of preharvest contamination accounted for), as well as in models considering each individual source of preharvest contamination separately **(a)**. The ECO157 prevalence in romaine batches contributed by each individual preharvest contamination source included in the model, one at a time **(b)**. ECO157 counts in contaminated romaine batches in the baseline irrigation model (considering only the three irrigation systems with no other preharvest sources), as well as in irrigation models considering each individual irrigation source contamination separately **(c)**. The ECO157 prevalence in romaine batches contributed by each individual irrigation contamination source included in the irrigation model, one at a time **(d)**.

We also evaluated the impact of three irrigation systems—overhead spray, furrow, and drip— on ECO157 counts in romaine batches (Figure 4 (c) and (d)). We found that overhead spray irrigation was the primary contributor to ECO157 counts in romaine batches (median ECO157 count: 4.00 Log_10_CFU/romaine batch, 5^th^ – 95^th^ percentile: 2.53 – 5.30), leading to ECO157 counts in romaine batches most frequently (0.47%). In contrast, we predicted that furrow and drip irrigation rarely caused ECO157 counts in romaine batches (0.04% and 0.02%, respectively) and resulted in substantially lower ECO157 counts when contamination occurred (Furrow median ECO157 count: 0.00 Log_10_CFU/romaine batch, 5^th^ – 95^th^ percentile: 0.00 – 1.40; drip median ECO157 count: 0.30 Log_10_CFU/romaine batch, 5^th^ – 95^th^ percentile: 0.00 – 1.54).

We ran a sensitivity analysis (using SRCC) for the variable parameters in the baseline irrigation model (considering only the three irrigation systems with no other preharvest contamination sources), as well as for each individual irrigation source contamination separately — i.e., (i) baseline irrigation model with contamination from only overhead spray irrigation, (ii) baseline irrigation model with contamination from only furrow irrigation, and (iii) baseline irrigation model with contamination from only drip irrigation. Similar to what was observed in the baseline model, we found a poor to fair correlation between the ECO157 counts in a given romaine batch and the ECO157 count in irrigation water 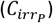. Other variables were also either poorly correlated or had no correlation with ECO157 counts in romaine batches, regardless of the irrigation system (Supplementary Table S10).

The preharvest regression tree captured the importance of irrigation and wildlife-related parameters in predicting ECO157 count in a romaine batch (Figure 5). The highest ECO157 counts in romaine batches were predicted when the ECO157 count in irrigation water was ≥ 0.3 CFU/100ml and applied via an overhead spray system. On the other hand, irrigation from furrow or drip systems led to substantially lower ECO157 counts in romaine batches. Also demonstrated was the importance of wildlife intrusions. When wildlife intrusion occurred for one or more days during the countdown period, it resulted in a mean of 1,000 CFU ECO157 per romaine batch, given that the fecal material from the feral swine contained 3 Log_10_CFU ECO157 or more per gram of feces.

**Figure 5.**
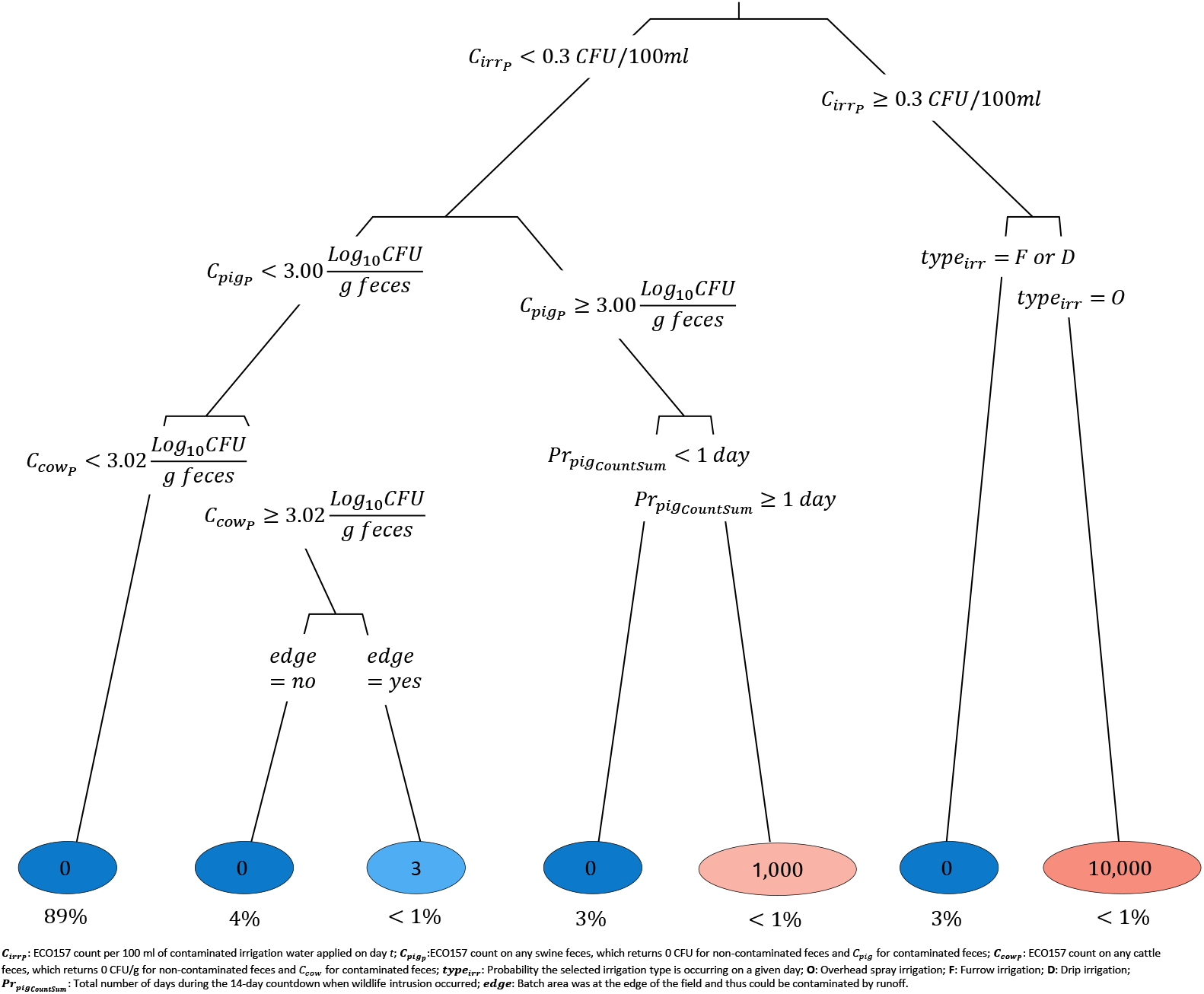
Preharvest model regression tree predicting the mean *Escherichia coli* O157:H7 (ECO157) counts in romaine batches under various preharvest conditions. Branch labels indicate the rules used for splitting each node, and the circled numbers at the end of each branch indicate the predicted mean ECO157 counts, expressed in CFU/romaine batch. The intensity of the circle colors was proportional to the value of predicted ECO157 counts for a given branch (blue: low; red: high). Underneath each circle (e.g., 0), the percentage of model iterations falling in that branch (e.g., 89%) are displayed (e.g., 89% is shown under 0, indicating that a mean ECO157 count of 0 CFU/romaine batch was observed for this branch, which appeared in 89% of 100,000 iterations).

### Postharvest model

The postharvest model predicted that the median number of cases resulting from the consumption of romaine was 19,040 annually (5^th^ – 95^th^ percentile: 42 – 3.6 ×10^6^). Although our predicted median number of illness cases aligned with the reported range for the annual median illness cases (12,189 – 19,893), the predicted distribution of illness cases was wider than the reported validation data (reported 90% CI: 3,394 – 47,134 cases annually). The validation data fell between 32^nd^ and 64^th^ percentiles of the predicted illness cases. Further predictions on hospitalizations, cases of haemolytic uraemic syndrome (HUS) and mortality were provided in Supplementary Table S11, based on CDC surveys on ECO157 outbreaks *[102]*. Considering the number of romaine servings consumed per year in the USA, the annual median risk of ECO157 due to consumption of ECO157-contaminated fresh-cut romaine was predicted as 0.000132% (or 1 illness in approximately 759,641 consumed servings). In addition, the predicted median prevalence of ECO157 contaminated romaine packages was 0.14% following the retail storage, but in 51% of the iterations the contaminated package (225g) had only 1 CFU per package.

The sensitivity analysis of variable parameters in postharvest model (Figure 6), determined by SRCC, showed that the median number of illness cases was very strongly correlated with the ECO157 count in romaine batches entering processing 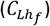. The remaining parameters were either poorly correlated or showed no correlation with the number of illness cases.

**Figure 6.**
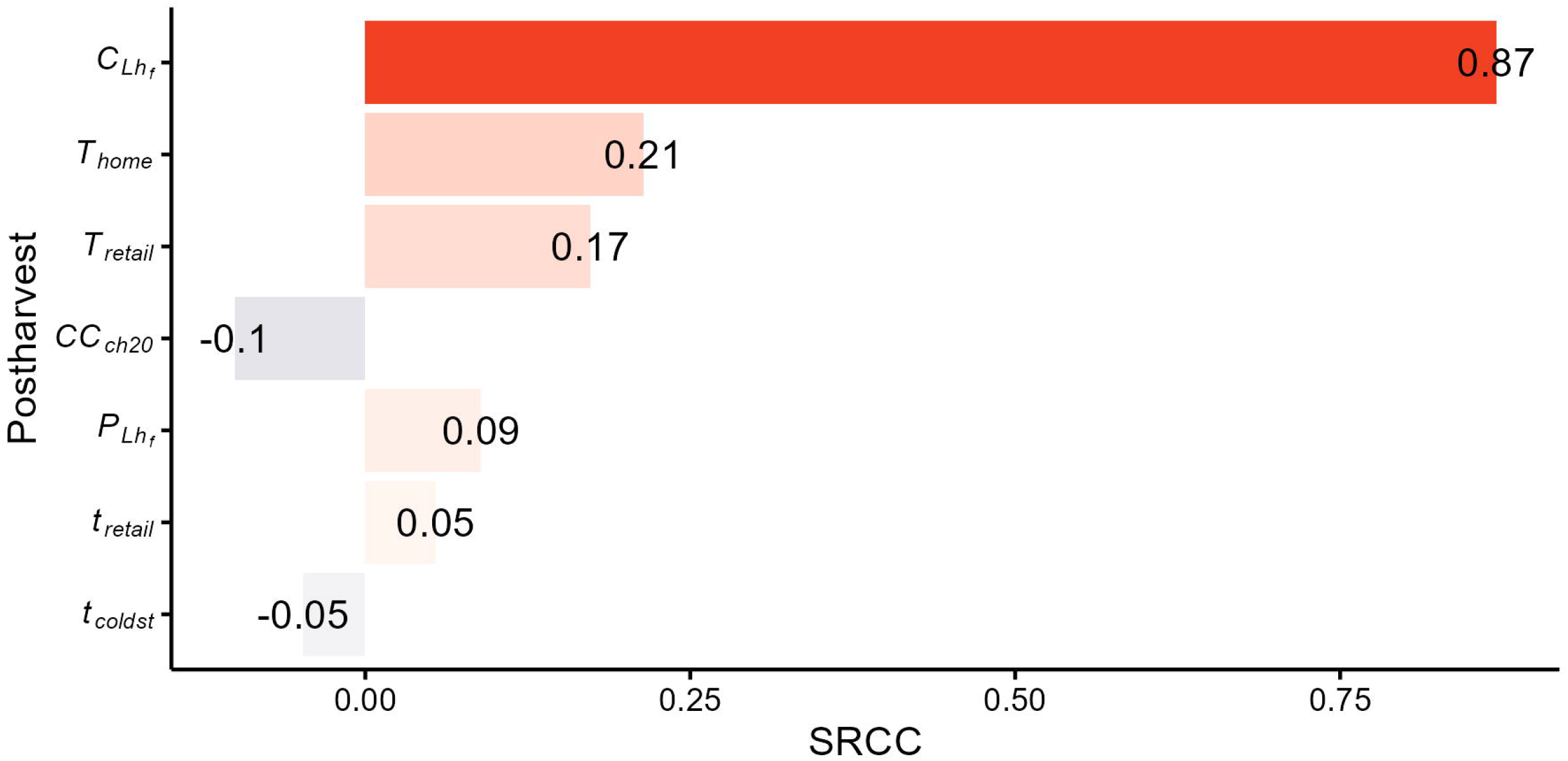
Tornado plot showing the impact of variable parameters on the postharvest output, i.e., the median predicted annual number of *E. coli* O157:H7 (ECO157) illness cases. The spearman rank-order correlation coefficients (SRCC) are shown next to each parameter bar. Only variables statistically significant after the Bonferroni correction and with a correlation coefficient > 0.05 are shown. Positive values indicate positive correlation (i.e., an increase in a given parameter increases the number of illness cases), while negative values indicate negative correlation (i.e., an increase in a given parameter decreases the number of illness cases).

The postharvest regression trees demonstrated how ECO157 counts, and consequently the number of illness cases, substantially increases due to both preharvest and postharvest conditions (Figure 7). Specifically, the ECO157 prevalence and counts in preharvest romaine batches, as well as the temperatures and durations of postharvest retail and home storage, play a critical role in this increase. In Figure 7 (a), the regression tree shows a high number of illness cases, although occurring with a small probability, when 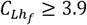 Log_10_CFU/romaine batch, *T*_*retail*_≥5^°^C and *T*_*home*_ ≥5^°^C. Illness cases were lowest when 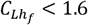 Log_10_ CFU/romaine batch and *T*_*home*_ < 5^°^C. Figure 7 (b) indicated that the ECO157 counts in contaminated servings were increased by a combination of time-temperature abuses during retail and home storage: ≥ 4 day storage at ≥ 8.9^°^C in retail, combined with home storage at ≥5.1^°^C resulted in the highest ECO157 counts in romaine servings. On the other hand, the ECO157 counts were lowest when 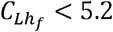 Log_10_ CFU/romaine batch, *T*_*retail*_< 9° C and *T*_*home*_< 9^°^C. Figure 7 (c) indicated that the highest prevalence of ECO157 contaminated romaine servings occurred when the preharvest ECO157 counts and prevalence in romaine batches was higher, along with retail storage at ≥ 5^°^C

**Figure 7.**
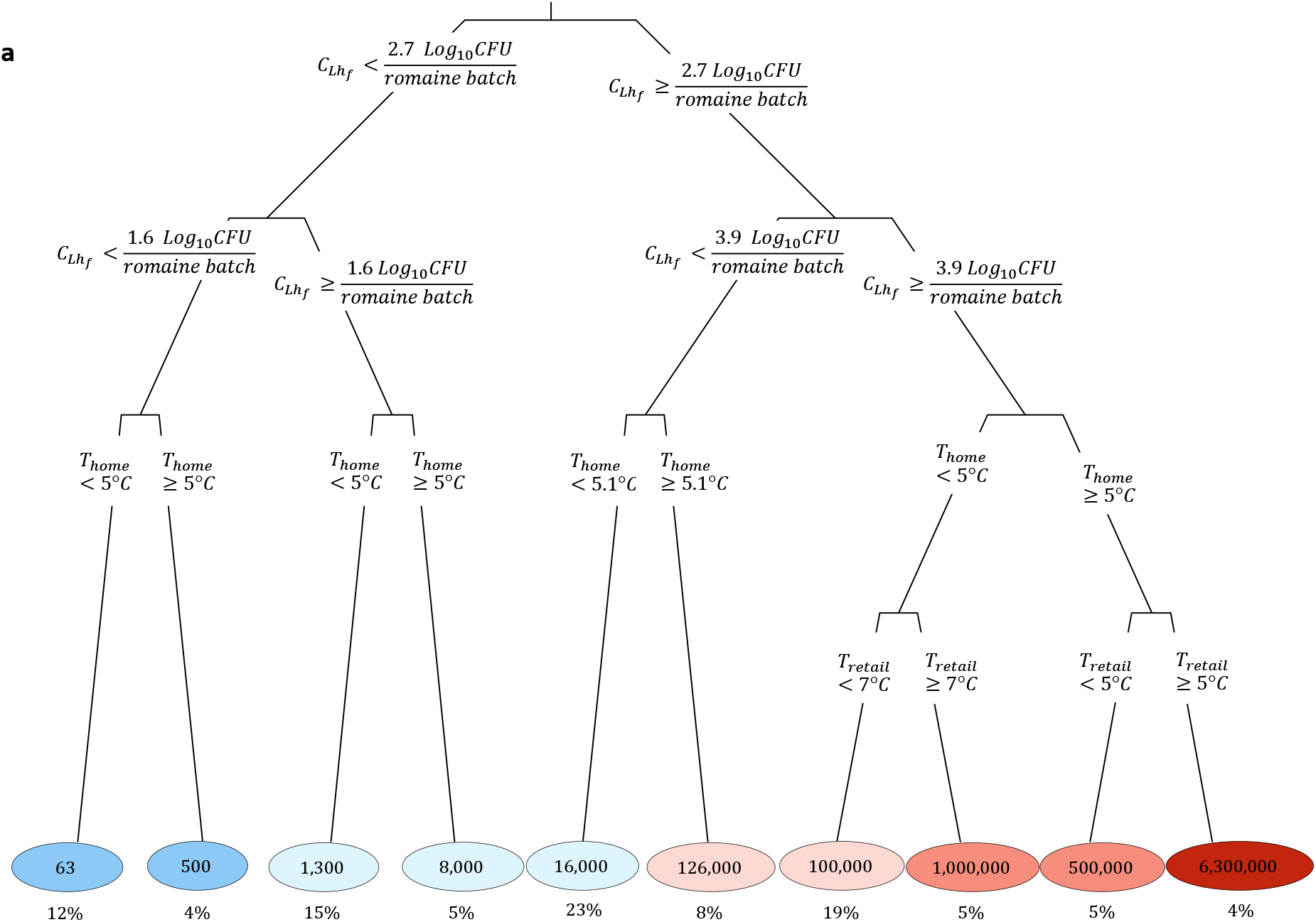
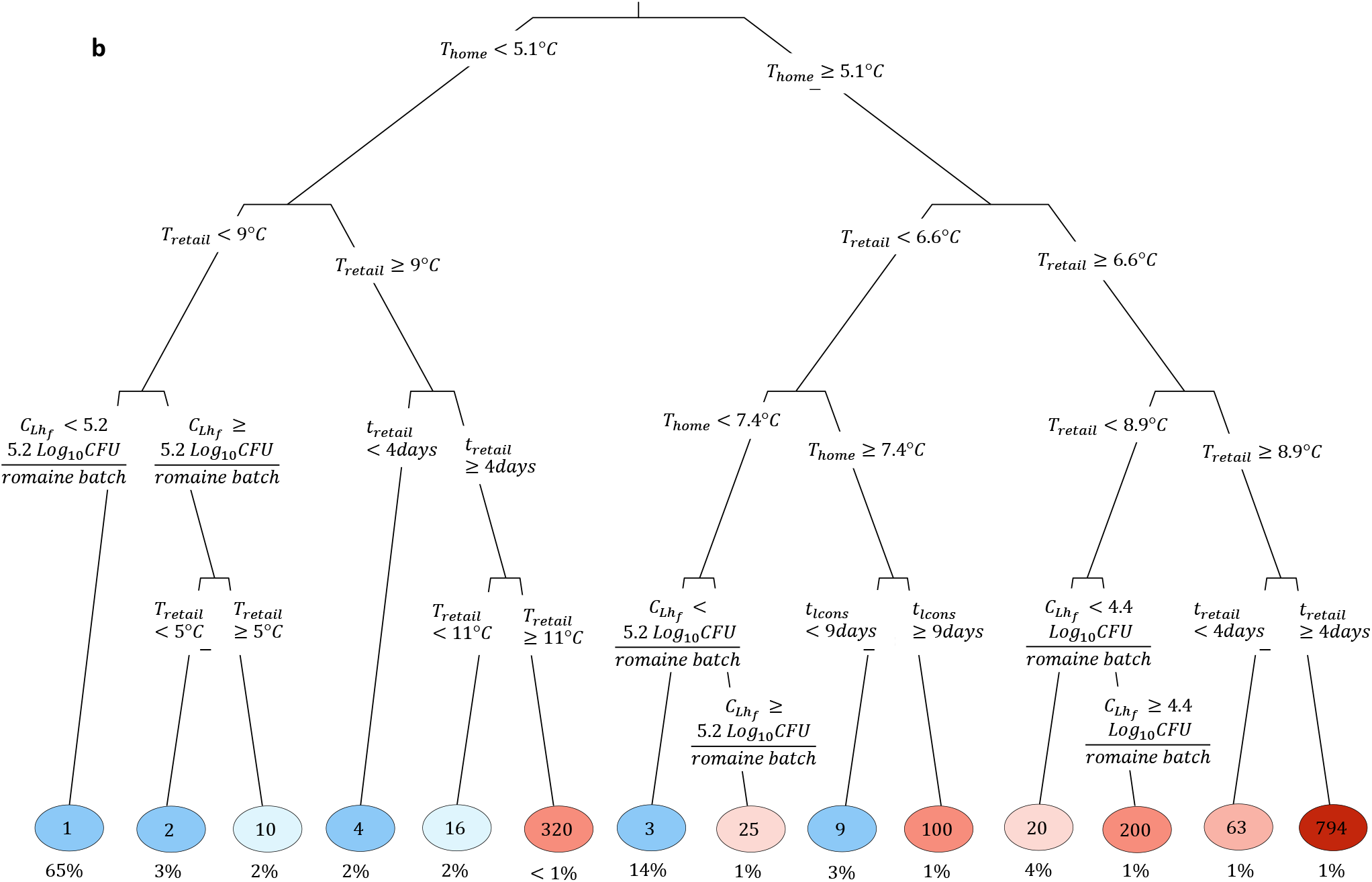
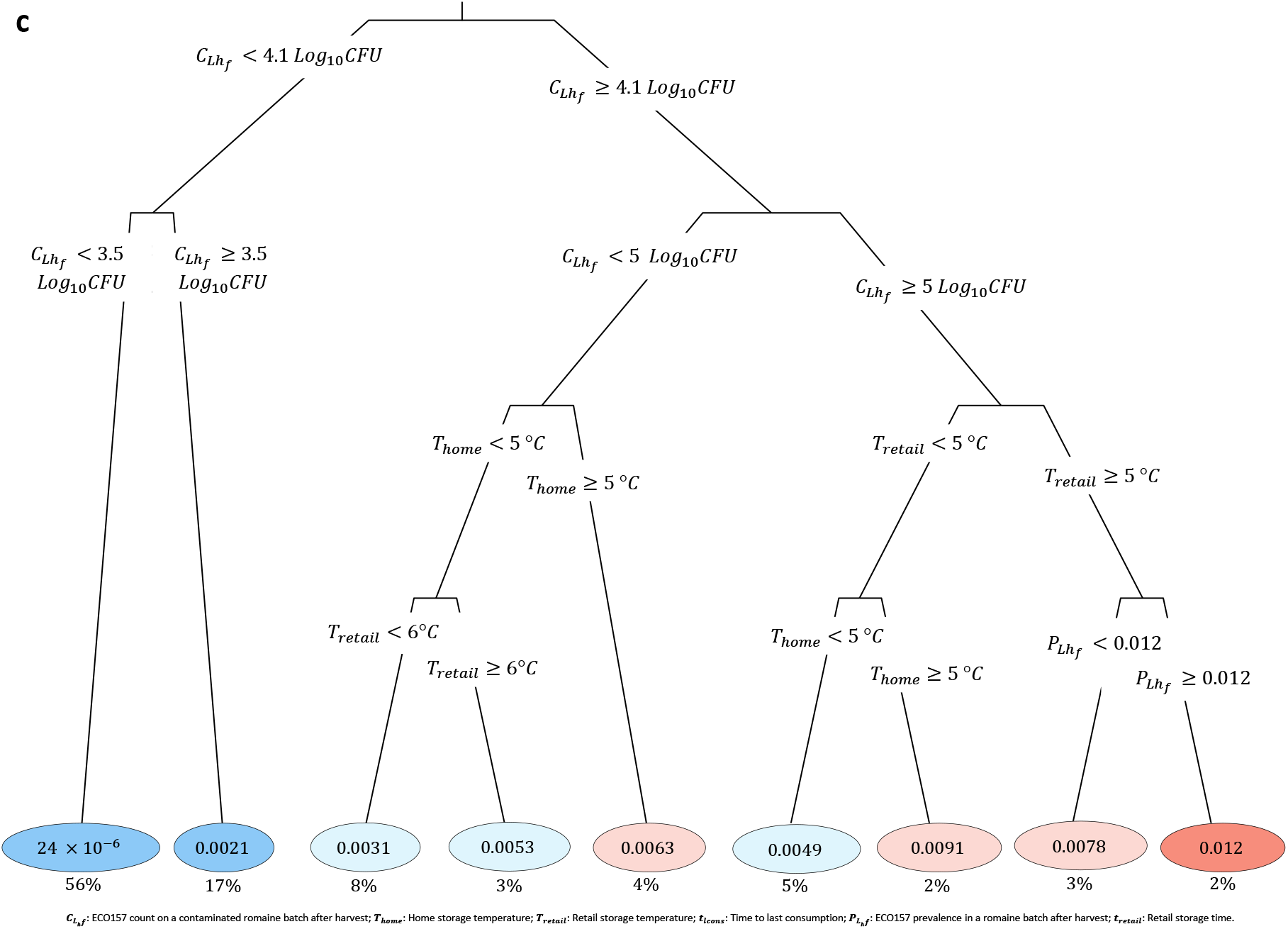
Regression trees for postharvest model, predicting illness outcome: **(a)** the mean number of illness cases from Escherichia coli O157:H7 (ECO157), **(b)** ECO157 counts in contaminated servings in CFU, and **(c)** ECO157 prevalence in romaine servings (unitless). Branch labels indicated the rules used for splitting each node, and the circled numbers at the end of each branch indicated the predicted mean value for the outcomes. The intensity of the circle colors was proportional to the value of predictions at the branch (blue: low; red: high). Underneath each circle (e.g., 63 cases), the percentage of model iterations falling in that branch (e.g., 12%) were displayed (e.g., 12% was shown under 63, indicating that a mean illness case number of 63 was observed for this branch, which appeared in 12% of 100,000 iterations).

In the preharvest model, none of the uncertainty variables were correlated with the predicted ECO157 count in romaine batches. Additionally, only approximately 0.3% of illness cases were attributed to the presence of persister cells in the baseline model, as detailed in Supplementary Table S12.

### Scenario analysis

In the 9 scenarios we tested (excluding clustering effect), the median number of illness cases decreased by 17.1% to 99.7% (Table 1). Out of the nine scenarios tested independently, six targeted reducing ECO157 counts at the preharvest stage. Among scenarios tested, the most effective was a postharvest “best case” wash alternative with 3.0 to 5.0 Log_10_CFU reduction *[97]*, which achieved a 99.7% reduction in median illness cases. The reduction rate in this scenario was a general estimate based on experimental results from various investigated wash methods—such as electrolyzed water combined with ultrasound or UV exposure, and alternative sanitizers like bacteriophages, phage lysins, or nanoparticles *[97]*. Because these alternative washes are still experimental and (i) likely cannot be implemented at scale or outside of laboratory settings and (ii) would not have been applied in the production of all lettuce produced nationwide, we also evaluated more moderate and realistic reductions in ECO157 levels. Specifically, we modeled additional scenarios with median reductions of 1.0, 1.5, 2.0 and 2.5 Log_10_CFU. These corresponded to reductions in the median number of illness cases by 61.9%, 87.3%, 95.7% and 98.4%, respectively. Surface water treatments and transitioning to furrow or drip irrigation resulted in slightly lower but highly effective reductions in ECO157 counts at the preharvest stage, achieving between 90.5% and 96.8% reductions in median illness cases. In contrast, consumer washing (28.0%), cattle vaccination (25.9%) and temperature reduction at retail (17.1%) were considerably less effective at reducing the number of illness cases.

The scenario analyses also showed that the degree of clustering assumed during the partitioning process affects the risk of illness. Increased clustering (in a batch and package) increased the dose of exposure and thus the probability of illness given exposure, but it also reduced the prevalence of ECO157 positive servings, which led to reduced number of cases (Table 1).

## Discussion

This risk assessment model simulated ECO157 counts during the production and consumption of fresh-cut romaine, providing a comprehensive view of how various preharvest and postharvest factors influence food safety risk. Our findings indicated that the model’s predictions of ECO157 counts, and associated illness cases aligned with surveillance studies in the USA. Irrigation emerged as the most significant source of ECO157 counts in fresh-cut romaine, particularly when considering the impact of other contamination sources: inadequately treated BSAAO, runoff, and wildlife intrusion. However, the public health impact could be mitigated through effective interventions. Our predictions also indicated a high number of illness cases due to the combined effects of preharvest and postharvest factors. Additionally, we identified key knowledge gaps in assessing human ECO157 exposure through contaminated leafy greens.

### The developed risk assessment tracks the dynamics of ECO157 cross-contamination between romaine and soil prior to harvest

We developed a novel difference equation model, which was incorporated into our preharvest model, quantifying ECO157 counts in both soil and romaine in the field. Similar approaches have been used in other risk assessment models, such as those describing the transmission of Listeria monocytogenes contamination in a smoked salmon facility *[103]*, Salmonella in pig slaughterhouses *[104]*, and Salmonella Typhimurium among farm swine *[105]*. This approach enabled us to track daily dynamics of ECO157 counts on romaine during the days prior to harvest. It allowed us to track the specific impacts of irrigation, wildlife, runoff and BSAAO; as well as to perform calculations related to ECO157 growth, biphasic decay and cross-contamination. Each of these factors has distinct characteristics that would be impossible to accurately capture with a static equation.

### Model predictions for preharvest and retail level ECO157 counts are consistent with previous reports

Our findings generally aligned with existing literature and foodborne illness surveillance data for the USA. The predicted annual median illness cases were in agreement with the average observed foodborne illnesses linked to ECO157-contaminated romaine in the USA. However, the distribution of the predicted illness cases was wider than the reported illness range. One potential reason for this discrepancy was that that we did not account for romaine loss/waste at the consumer stage, assuming instead that all retail romaine would be consumed. This assumption is likely inaccurate, given that it has been estimated that 39.7% lettuce loss occurred at the consumer level in the USA in 2011 *[18]*. Another reason for the right skewness may be explained by the interactions between various parameters in the model, which will be discussed in the following sections. The predicted ECO157 prevalence at preharvest (1.00%) falls within the range of previously reported values for leafy greens from CA’s central coast region [106], which ranged from <0.01% in 2007 to 2.5% in 2013. While a previous study by Zhang et al. (2018) reported a 0.02% average prevalence of ECO157 on romaine at the retail level in the USA (95% Confidence Interval by Clopper-Pearson method: <0.01 - 0.10, based on 5,548 samples of 25g each) *[11]*, our predicted median ECO157 prevalence of contaminated romaine packages was over 7 times higher (0.14%, predicted for 225g packages). One potential explanation for the discrepancy between the reported and predicted prevalence could be the limitation of microbial tests in detecting contamination when smaller sample sizes are used. For instance, in Zhang et al.’s (2018) study, the detection limit was 1 CFU in a 25g sample (BAM method for diarrheagenic *E. coli*), equivalent to 0.04 CFU/g. For a 225g sample—such as the packages in our model—the detection limit would be 1 CFU per 225g, or 0.0044 CFU/g. Under this scenario, testing the entire 225g package could detect ECO157 levels 9 times lower than a 25g sample. Assuming a 225g sample size, estimating ECO157 counts could yield a prevalence 9 times higher than that observed in retail samples, potentially leading to 0.18% (0.02% × 9) ECO157 prevalence. In our risk assessment model, we considered any package (and serving) contaminated if it contained even a single ECO157 cell. Nevertheless, the previously observed ECO157 prevalence (0.02%) *[11]* falls within the predicted distribution of ECO157 prevalence simulated in our study (*P*_*Pack*_,Supplementary Table S2).

### Irrigation is predicted to be the most important source of ECO157 contamination of fresh-cut romaine with appropriate interventions reducing public health impact

Our findings suggested that mitigation strategies at preharvest should prioritize reducing contamination of irrigation water, particularly in cases of overhead spray irrigation supplied via surface water sources. Aligned with our findings, contaminated irrigation water has been defined as a major risk factor in the contamination of leafy greens *[11]*. Notably, contaminated surface water irrigation has either been directly linked to or strongly associated with several multistate ECO157 outbreaks of leafy greens in the USA *[107,108,109]*, including spinach *[107]* and iceberg lettuce outbreaks in 2006 *[109]*, and a romaine outbreak in 2018 *[108]*. For the romaine outbreak in 2018, ECO157 introduction into the irrigation water through aerial and land-based spray applications of crop protection chemicals was proposed as a plausible route of introduction *[110]*. On the other hand, the spinach and iceberg lettuce outbreaks appeared to have been facilitated by cross-contamination, where contaminated surface water and wastewater flowed into groundwater or surface water used for irrigating the lettuce plants *[107,109]*.

Our model also suggests that treatment of surface irrigation water—an important intervention strategy already required by the California and Arizona LGMAs since the 2023 update—may achieve a substantial reduction of human illness cases. This outcome was predicted for various surface water treatments, including chlorination, PAA or UV. We consistently observed a strong effectiveness across all treatment methods in reducing illness cases, based on the assumption that the different methods achieved ECO157 reduction in surface irrigation water, ranging from a minimum of 1.00 Log_10_ CFU/ml (chlorination) to a maximum of 3.09 Log_10_ CFU/ml (PAA). Previous research has also reported that these methods reduce *E. coli* counts by up to 6 Log_10_CFU/ml in agricultural water sources, with PAA typically having the strongest impact *[96,111-116]*. However, the effectiveness of these methods can be highly variable, even when implemented correctly *[115]*. Their site-specific applicability is influenced by a range of factors including water quality (such as total suspended solids, turbidity, pH, oxygen demand and temperature), applied dose, contact time, and contaminant characteristics *[117-119]*. In an open leafy green production field, these factors can be affected by rainfall, runoff or sediment activity upstream of the water sources *[116]*. On the other hand, both California and Arizona LGMAs prohibit the use of surface irrigation waters that fail to meet the performance requirements. To use such waters, treatment and testing are required to demonstrate treatment efficacy and compliance with microbial standards. Therefore, future models should (i) better account for the variability in treatment effectiveness and (ii) assess the impact of reducing treatment variability: A less variable treatment with lower Log_10_CFU/ml reduction may be more effective than one with a higher reduction but greater variability. Addressing these challenges can help leafy green growers, who must navigate multiple treatment options influenced by various factors while meeting LGMAs’ requirements, make more informed decisions.

Our data also suggested that transitioning from overhead spray irrigation to furrow or, ideally, drip irrigation systems could provide a viable solution for reducing illness cases (Figure 4, Table 1). Despite introducing numerous parameters specific to each irrigation system that reasonably differentiate between the three irrigation systems, our model suggested that the primary driver behind the higher number of illness cases associated with overhead spray irrigation was its capacity for increased direct contact between irrigation water and romaine (Supplementary Table S10). Conversely, drip irrigation, delivering irrigation water to the plant roots, minimized water contact, and consequently resulted in the lowest number of illness cases among the three irrigation systems. Numerous field studies *[120-122]* and risk assessments *[40,41]* have consistently demonstrated that overhead spray irrigation system leads to higher counts of generic *E. coli* and ECO157 on romaine compared to furrow or drip irrigation systems, and argued that the type of irrigation system plays a critical role in mitigating the risk of ECO157 contamination on leafy greens *[111,123,124]*. Aligned with our model’s results, research also supports that furrow irrigation leads to greater produce contamination than drip irrigation, highlighting drip irrigation as the most effective irrigation method for reducing health risks from contaminated irrigation water *[40,72,125]*. In our study, transitioning fully from overhead spray and furrow irrigation to drip irrigation was predicted to be more effective than chlorination and UV treatments of surface irrigation water, though less effective than PAA treatment.

### The combined effect of preharvest and postharvest factors defines the public health risk

The developed model illustrated that interactions of preharvest and postharvest factors affect the number of predicted illness cases (Figure 7). This aligns with findings from observational studies, which support the multifactorial nature of food safety *[126]*. Our model highlighted how certain co-occurring factors could, though rarely, lead to a very large number of illnesses. Specific factors, such as higher ECO157 counts in irrigation water, the use of overhead spray irrigation systems instead of furrow or drip irrigation, or increased number of wildlife intrusion, led to elevated ECO157 counts on preharvest romaine (Figure 5), which were further increased by improper handling and storage conditions during postharvest, including time-temperature abuse from harvest until consumption (Figure 7). This highlights the importance of simulation tools like the one developed here that can be used to study impacts and prevention of rare events (occurring individually or in combination) which would be challenging to study empirically.

Several aspects of farm-to-fork continuum of fresh-cut romaine production and consumption were included in the model but did not emerge as significant risk factors. These included harvesting months, weather conditions (i.e., precipitation intensity, number of sunny days, decay related to sun exposure) and several others. Weather conditions, including temperature, seasonality and precipitation events, have previously shown to influence pathogen counts on leafy greens *[10,35,127]*. In addition, historically, more leafy greens outbreaks have occurred in summer and fall seasons than spring and winter seasons in the USA *[1]*. The likely reason why our model did not capture the significance of these factors, including weather conditions, was that weather conditions alone was insufficient to exacerbate the risk of contamination unless cooccurring with other factors that introduce ECO157 contamination into the field. For example, heavy rain will introduce contamination via runoff from a neighboring dairy farm only if there was a nearby dairy farm and cattle on the farm that shed ECO157. Similarly, heavy rain would contaminate romaine via splash but only if soil was already contaminated. In addition, it was possible that certain factors that may increase ECO157 contamination during certain months or under certain weather conditions were not or not fully included in our model (e.g., increased leaf damage or blistering induced by certain weather conditions) *[128]*.

Our study shows that a “best case” highly effective produce washing (3.0 to 5.0 Log_10_CFU/g reduction) is predicted to reduce the illness cases by over 99%, even with high preharvest contamination levels (Table 1). This best case scenario was based on research studies on alternative sanitizers that reported reductions around 5.0 Log_10_CFU; however, most of these studies have been limited to small-scale experiments and have not been adopted by industry *[97]*. This high level of reduction however appears to typically be unachievable, at least with current approaches and at realistic production scales, as indicated by studies that suggest substantially lower reduction rates *[97]*. In the baseline model, the impact of postharvest washing on illness risk was limited due to the use of a conservative and likely more realistic ECO157 reduction — a maximum of 1.03 Log_10_CFU/g reduction (Figure 6). There are however some reports that suggest that chlorine-based produce washes can achieve higher ECO157 reductions by preventing cross-contamination of ECO157 in the romaine batch during washing *[17,129]*. Importantly, our scenario analysis (Table 1) showed that even a modest improvement—such as a median reduction of 1.5 Log_10_CFU/g rather than the baseline 1.03 Log_10_CFU/g reduction, or just ∼0.5 Log_10_CFU/g improvement—could result in 61.9% decrease in predicted median illness cases. This supports the importance of continuous improvement of produce wash treatments as even relatively small improvements in effectiveness may lead to tangible public health improvements. Our findings also suggest that it is important for industry to verify produce wash effectiveness under realistic commercial scale conditions to correctly assess risk reductions that can be expected by a given produce wash system. The reduction is influenced by various factors, including the concentration and state (free or bound) of chlorine, pH of the water, the duration of contact time, and the characteristics of the organism *[84,130,131]*. Addressing these parameters has been the focus of multiple studies, which aim to close knowledge gaps on produce wash efficacy *[130,132-134]*.

### Model limitations and key knowledge gaps

Our model here reflected a synthesis of the current knowledge on ECO157 contamination throughout the farm-to-fork supply chain of fresh-cut romaine lettuce in the USA. However, we acknowledge limitations related to the model structure and parameterization. The most consequential assumption in our model was about dilution factors in preharvest, which facilitated modeling the spread of ECO157 from BSAAO, runoff, irrigation and fecal material on the field. For ECO157 spread via BSAAO, we assumed a typical tiling depth of 15cm to 20cm. In contrast, for irrigation, swine feces introduction caused by feral swine, and runoff, we assumed a uniform spread of ECO157 to a depth of 1cm. As shown in Supplementary Table S13, the depth significantly impacts the predicted ECO157 counts. Therefore, our analysis indicates that using a 1cm depth was a conservative approach because the risk of romaine contamination from contaminated soil decreases as the depth of bacterial spread increases. Despite this, our assumptions about spread were considered reasonable based on the complexity of acquiring an accurate depth of ECO157 spread in these various media (runoff, swine feces and BSAAO) within 24 hours of their introduction to the field. Another limitation was that, while BSAAOs of poultry origin are the most common in fields across the USA, our study focused on those of dairy origin. Another assumption in the preharvest model was a constant and low-level ECO157 transfer from water and soil to romaine through furrow and drip irrigation. However, since these irrigation methods involve less direct contact with the romaine compared to overhead spray irrigation, we do not expect this assumption to have a major impact on the study conclusions —a finding also supported by the preharvest uncertainty analysis. Additionally, we did not account for the exterior leaves removed in the field, as we do not know the fraction of cells on these outer vs inner leaves. However, based on our assumptions, the impact of this practice was minimal as a 33.3% reduction in the ECO157 count on romaine resulted in 13.5% reduction in the median predicted illness cases (as detailed in Supplementary Table S14). Preharvest contamination was modeled at batch level, ignoring the spatial distribution of contamination among plants and soil in different locations of the field. However, we consider this to be a reasonable simplification since romaine was washed in batches, and the process of washing may further distribute ECO157 within a romaine batch. The preharvest model focused on open-field romaine production, excluding other potential production methods, such as greenhouses or vertical farming. This decision is expected to have minimal impact, as the majority of lettuce in the USA is grown in open fields *[101]*.

During postharvest, presence of cell clustering during partitioning — a poorly understood phenomenon in romaine packaging and servings — was predicted to moderately reduce illness cases (up to 48% reduction when high clustering). This reduction was due to the prediction of a lower ECO157 prevalence in batches, as clustering decreased the probability of consumers being exposed to servings with high ECO157 counts, as also predicted by Zoellner et al. (2019) *[85]*. Clustering acted as a form of containment, minimizing the spread of contamination. For the industry, this insight may encourage mechanical processing that minimizes disturbance of clustered cells, preventing cells from spreading across the produce. For example, preventing cross-contamination during postharvest wash process (e.g., by adding chlorine), helps contain contamination within a single romaine batch, thereby reducing the risk of human illness (Table 1). We assumed that 100% of annual romaine consumption in the US is produced domestically, even though USDA estimates suggest that only about 85% is US-grown *[138]*, based on reported import and export data *[139]*. We acknowledge this as a limitation of our study. The dose-response model used here describes the distribution of susceptibility to ECO157 and accounts for population variability. Based on the assumptions of the Beta-Poisson model, (i) each cell is capable of causing illness, and (ii) each cell is equally infective *[87]*. However, ECO157 illness is a complex phenomenon influenced by factors such as the genetic diversity of ECO157 strains, variations in virulence factors, and host susceptibility. As a result, some strains may have a higher or lower infectious dose (ID_50_), which is estimated to be between 10 and 100 bacteria, according to the National Advisory Committee on Microbiological Criteria for Foods (NACMFS) *[140]*. Therefore, our results should be interpreted with caution, as these factors are not fully captured by the model.

## Conclusion

The risk assessment model presented here provides a comprehensive analysis of the impact of various preharvest and postharvest factors and their interactions. Our results suggest that illnesses occur mainly (52%) due to ECO157 contamination originating from untreated irrigation water applied through overhead spray irrigation. Our data further suggests that preharvest ECO157 risk, including that from irrigation, can be reduced either through water treatments or by switching to furrow or drip irrigation. In postharvest, maintaining the cold chain is crucial to prevent high numbers of illness cases. While our data indicates that an effective postharvest wash can help mitigate the public heath impact of both preharvest and postharvest contamination, future work on the impact of postharvest wash variability on public health risk is important. We identify knowledge gaps regarding the microbial quality of irrigation waters, as well as the effectiveness of preharvest surface water treatments and postharvest wash processes. In conclusion, the developed QMRA is expected to inform development of sustainable food safety strategies along the supply chain and guide further research to address key knowledge gaps.

## Acknowledgements

This work was supported through a grant (award 2019-51181-30016) from the U.S. Department of Agriculture (USDA), National Institute of Food and Agriculture (NIFA). Partial support received from a grant (award 2020-67021-32855) from the USDA NIFA.

## Author contributions

E.B.: conceptualization, data curation, formal analysis, investigation, methodology, software, validation, visualization, writing-original draft, writing – review and editing; S.I.M.: conceptualization, data curation, methodology, software, writing – review and editing; L.K.S.: conceptualization, funding acquisition, writing – review and editing; M.D.D.: conceptualization, funding acquisition, writing – review and editing; M.W.: conceptualization, funding acquisition, writing – review and editing; R.I.: conceptualization, funding acquisition, investigation, methodology, project administration, software, supervision, validation, visualization, writing – review and editing. All authors have seen and approved the manuscript.

## Data availability statement

The code associated with this project is stored in the GitHub repository linked here: https://github.com/IvanekLab/Romaine_ECO157_QMRA/ and is publicly accessible.

## References

1. Turner, K., Moua, C. N., Hajmeer, M., Barnes, A. & Needham, M. Overview of leafy greens–related food safety incidents with a California link: 1996 to 2016. J. Food Prot. 82, 405–414 (2019).

2. CDC. NORS Dashboard. https://wwwn.cdc.gov/norsdashboard/ (2022).

3. CDC. Active investigations in foodborne outbreaks. https://www.cdc.gov/foodborne-outbreaks/active-investigations/all-foodborne-outbreak-notices.html (2024).

4. Coulombe, G., Catford, A., Martinez-Perez, A. & Buenaventura, E. Outbreaks of Escherichia coli O157:H7 infections linked to romaine lettuce in Canada from 2008 to 2018: An analysis of food safety context. J. Food Prot. 83, 1444–1462 (2020). 10.4315/JFP-20-029

5. Benjamin, L. A. et al. Risk factors for Escherichia coli O157 on beef cattle ranches located near a major produce production region. Epidemiol. Infect. 143, 81–93 (2015).

6. Jay, M. T. et al. Escherichia coli O157:H7 in feral swine near spinach fields and cattle, central California coast. Emerg. Infect. Dis. 13, 1908 (2007).

7. California Food Emergency Response Team (CalFERT). Investigation of an Escherichia coli O157:H7 outbreak associated with Dole pre-packaged spinach. California Department of Health Services, Sacramento, CA (2007).

8. Cooley, M. B. et al. Development of a robust method for isolation of Shiga toxin-positive Escherichia coli (STEC) from fecal, plant, soil, and water samples from a leafy greens production region in California. PLOS ONE 8, e65716 (2013). 10.1371/journal.pone.0065716

9. Atwill, E. R. et al. Transfer of Escherichia coli O157:H7 from simulated wildlife scat onto romaine lettuce during foliar irrigation. J. Food Prot. 78, 240–247 (2015). 10.4315/0362-028X.JFP-14-277

10. Oliveira, M., Viñas, I., Usall, J., Anguera, M. & Abadias, M. Presence and survival of Escherichia coli O157:H7 on lettuce leaves and in soil treated with contaminated compost and irrigation water. Int. J. Food Microbiol. 156, 133–140 (2012). 10.1016/j.ijfoodmicro.2012.03.014

11. Zhang, G. et al. Survey of foodborne pathogens, aerobic plate counts, total coliform counts, and Escherichia coli counts in leafy greens, sprouts, and melons marketed in the United States. J. Food Prot. 81, 400–411 (2018).

12. Ackers, M. L. et al. An outbreak of Escherichia coli O157:H7 infections associated with leaf lettuce consumption. J. Infect. Dis. 177, 1588–1593 (1998).

13. Hillborn, E. D. et al. A multistate outbreak of Escherichia coli O157:H7 infections associated with consumption of mesclun lettuce. Arch. Intern. Med. 159, 1758–1764 (1999).

14. FAO. Prevention and control of microbiological hazards in fresh fruits and vegetables, Part 4: Specific commodities. https://openknowledge.fao.org/server/api/core/bitstreams/151c9dda-508b-4b70-972a-85725eb12145/content (2023).

15. Yang, Y., Luo, Y., Millner, P., Turner, E. & Feng, H. Assessment of Escherichia coli O157:H7 transference from soil to iceberg lettuce via a contaminated field coring harvesting knife. Int. J. Food Microbiol. 153, 345–350 (2012).

16. Buchholz, A. L., Davidson, G. R., Marks, B. P., Todd, E. C., Ryser, E. T. Transfer of Escherichia coli O157:H7 from equipment surfaces to fresh-cut leafy greens during processing in a model pilot-plant production line with sanitizer-free water. J. Food Prot. 75, 1920–1929 (2012). 10.4315/0362-028X.JFP-11-558

17. López-Gálvez, F., Allende, A., Selma, M. V. & Gil, M. I. Prevention of Escherichia coli cross-contamination by different commercial sanitizers during washing of fresh-cut lettuce. Int. J. Food Microbiol. 133, 167–171 (2009). 10.1016/j.ijfoodmicro.2009.05.017

18. USDA ERS. Food availability per capita data system. https://www.ers.usda.gov/data-products/food-availability-per-capita-data-system/ (2020).

19. California LGMA. About us. https://lgma.ca.gov/about-us (2024).

20. Arizona LGMA. About us. https://www.arizonaleafygreens.org/about-us (2023).

21. FDA. FSMA Final Rule on Produce Safety: Standards for the Growing, Harvesting, Packing, and Holding of Produce for Human Consumption. https://www.fda.gov/food/food-safety-modernization-act-fsma/fsma-final-rule-produce-safety (2024).

22. FSMA PSR § 112.41, § 112.42. https://www.ecfr.gov/current/title-21/chapter-I/subchapter-B/part-112 (2024).

23. CFR § 205.203. https://www.ecfr.gov/current/title-7/subtitle-B/chapter-I/subchapter-M/part-205/subpart-C/section-205.203 (2024).

24. CFR § 112.55. https://www.ecfr.gov/current/title-21/chapter-I/subchapter-B/part-112/subpart-F/section-112.55 (2024).

25. CFR §lil112.83 and §112.112. https://www.regulations.gov/document/FDA-2011-N-0921-18558 (2024).

26. FSMA PSR. https://www.regulations.gov/document/FDA-2011-N-0921-18558 (2015).

27. Allende, A., Truchado, P., Lindqvist, R. & Jacxsens, L. Quantitative microbial exposure modelling as a tool to evaluate the impact of contamination level of surface irrigation water and seasonality on fecal hygiene indicator E. coli in leafy green production. Food Microbiol. 75, 82–89 (2018).

28. Koseki, S. & Isobe, S. Prediction of pathogen growth on iceberg lettuce under real temperature history during distribution from farm to table. Int. J. Food Microbiol. 104, 239–248 (2005).

29. Franz, E., Semenov, A. V. & van Bruggen, A. H. Modelling the contamination of lettuce with Escherichia coli O157:H7 from manure-amended soil and the effect of intervention strategies. J. Appl. Microbiol. 105, 1569–1584 (2008).

30. Franz, E., Tromp, S. O., Rijgersberg, H. & Van Der Fels-Klerx, H. J. Quantitative microbial risk assessment for Escherichia coli O157:H7, Salmonella, and Listeria monocytogenes in leafy green vegetables consumed at salad bars. J. Food Prot. 73, 274–285 (2010).

31. Tromp, S. O., Rijgersberg, H. & Franz, E. Quantitative microbial risk assessment for Escherichia coli O157:H7, Salmonella enterica, and Listeria monocytogenes in leafy green vegetables consumed at salad bars, based on modeling supply chain logistics. J. Food Prot. 73, 1830–1840 (2010).

32. Danyluk, M. D. & Schaffner, D. W. Quantitative assessment of the microbial risk of leafy greens from farm to consumption: preliminary framework, data, and risk estimates. J. Food Prot. 74, 700–708 (2011).

33. Rodríguez, F. P. et al. A mathematical risk model for Escherichia coli O157:H7 cross-contamination of lettuce during processing. Food Microbiol. 28, 694–701 (2011).

34. Ottoson, J. R., Nyberg, K., Lindqvist, R. & Albihn, A. Quantitative microbial risk assessment for Escherichia coli O157 on lettuce, based on survival data from controlled studies in a climate chamber. J. Food Prot. 74, 2000–2007 (2011).

35. Castro-Ibáñez, I., Gil, M. I., Tudela, J. A., Ivanek, R. & Allende, A. Assessment of microbial risk factors and impact of meteorological conditions during production of baby spinach in the Southeast of Spain. Food Microbiol. 49, 173–181 (2015).

36. Mishra, A., Pang, H., Buchanan, R. L., Schaffner, D. W. & Pradhan, A. K. A system model for understanding the role of animal feces as a route of contamination of leafy greens before harvest. Appl. Environ. Microbiol. 83, e02775–16 (2017).

37. Kundu, A., Wuertz, S. & Smith, W. A. Quantitative microbial risk assessment to estimate the risk of diarrheal diseases from fresh produce consumption in India. Food Microbiol. 75, 95–102 (2018).

38. Pang, H, Lambertini, E., Buchanan, R. L., Schaffner, D. W. & Pradhan, A. K. Quantitative microbial risk assessment for Escherichia coli O157:H7 in fresh-cut lettuce. J. Food Prot. 80, 302–311 (2017).

39. O’Flaherty, E., Solimini, A. G., Pantanella, F., De Giusti, M. & Cummins, E. Human exposure to antibiotic-resistant Escherichia coli through irrigated lettuce. Environ. Int. 122, 270–280 (2019).

40. Rock, C. M. et al. Review of water quality criteria for water reuse and risk-based implications for irrigated produce under the FDA Food Safety Modernization Act, produce safety rule. Environ. Res. 172, 616–629 (2019).

41. Bozkurt, H., Bell, T., van Ogtrop, F., Phan-Thien, K. Y. & McConchie, R. Assessment of microbial risk during Australian industrial practices for Escherichia coli O157:H7 in fresh-cut cos lettuce: A stochastic quantitative approach. Food Microbiol. 95, 103691 (2021).

42. FDA. FDA-iRISK. https://irisk.foodrisk.org/ (2025).

43. USDA. Risk assessment of E. coli O157:H7 in ground beef. https://www.fsis.usda.gov/node/2003 (2001).

44. Ebel, E. et al. Draft risk assessment of the public health impact of Escherichia coli O157:H7 in ground beef. J. Food Prot. 67, 1991–1999 (2004).

45. USDA. Microbial risk assessment guideline. Pathogenic microorganisms with focus on food and water. https://www.fsis.usda.gov/sites/default/files/media_file/2020-07/Microbial_Risk_Assessment_Guideline_2012-001.pdf (2012).

46. Nauta, M. J. Microbiological risk assessment models for partitioning and mixing during food handling. Int. J. Food Microbiol. 100, 311–322 (2005).

47. Munther, D. S., Carter, M. Q., Aldric, C. V., Ivanek, R. & Brandl, M. T. Formation of Escherichia coli O157:H7 persister cells in the lettuce phyllosphere and application of differential equation models to predict their prevalence on lettuce plants in the field. Appl. Environ. Microbiol. 86, e01602–19 (2020).

48. Turini, T. et al. Iceberg lettuce production in California. https://escholarship.org/uc/item/7w47j6zv (2011).

49. Scallan, E. et al. Foodborne illness acquired in the United States—major pathogens. Emerg. Infect. Dis. 17, 7–15 (2011).

50. Painter, J. A. et al. Attribution of foodborne illnesses, hospitalizations, and deaths to food commodities by using outbreak data, United States, 1998–2008. Emerg. Infect. Dis. 19, 407–415 (2013).

51. Longfellow’s Greenhouses. http://plants.longfellowsgreenhouses.com/12100007/Plant/Print?id=29076.

52. Rural Migration News (RMN). https://migration.ucdavis.edu/rmn/blog/post/?id=2699 (2022).

53. Agriculture Marketing Resource Center. Lettuce. https://www.agmrc.org/commodities-products/vegetables/lettuce (2021).

54. Geisseler, D. & Horwath, W. R. Lettuce production in California. https://www.cdfa.ca.gov/is/ffldrs/frep/FertilizationGuidelines/pdf/Lettuce_Production_CA.pdf (2016).

55. Brouwer, A. F. et al. Modeling biphasic environmental decay of pathogens and implications for risk analysis. Environ. Sci. Technol. 51, 2186–2196 (2017).

56. Bezanson, G. et al. Comparative examination of Escherichia coli O157:H7 survival on romaine lettuce and in soil at two independent experimental sites. J. Food Prot. 75, 480– 487 (2012).

57. Erickson, M. C. et al. Surface and internalized Escherichia coli O157:H7 on field-grown spinach and lettuce treated with spray-contaminated irrigation water. J. Food Prot. 73, 1023–1029 (2010).

58. Moyne, A. L. et al. Fate of Escherichia coli O157:H7 in field-inoculated lettuce. Food Microbiol. 28, 1417–1425 (2011).

59. Williams, T. R., Moyne, A. L., Harris, L. J. & Marco, M. L. Season, irrigation, leaf age, and Escherichia coli inoculation influence the bacterial diversity in the lettuce phyllosphere. PLoS One 8, e68642 (2013).

60. Chen, Z. et al. Prevalence of Escherichia coli O157 and Salmonella spp. in solid bovine manure in California using real-time quantitative PCR. Lett. Appl. Microbiol. 69, 23–29 (2019).

61. Rogers, S. W. et al. Decay of bacterial pathogens, fecal indicators, and real-time quantitative PCR genetic markers in manure-amended soils. Appl. Environ. Microbiol. 77, 4839–4848 (2011).

62. Chen, H. Contribution of the persister state to the tolerance of Escherichia coli O157:H7 to stresses encountered within the fresh produce chain. Master’s Thesis, University of Guelph (2020).

63. Gravuer, K. Compost application rates for California croplands and rangelands for a CDFA Healthy Soils Incentives Program. https://www.cdfa.ca.gov/oefi/healthysoils/docs/CompostApplicationRate_WhitePaper.pdf (2016).

64. Aqua-Calc. https://www.aqua-calc.com/ (2023).

65. Valenzuela, H. R., Kratky, B. & Cho, J. Lettuce production guidelines for Hawaii. https://www.ctahr.hawaii.edu/oc/freepubs/pdf/res-164.pdf (1995).

66. California Department of Fish and Wildlife. Wild pig. https://wildlife.ca.gov/Hunting//Wild-Pig (2001).

67. Hamilton, D. W., Luce, W. G. & Heald, A. D. Production and characteristics of swine manure. Oklahoma State University Extension Facts Publication F-1735 (1997).

68. Ferretti, F., Storer, K., Coats, J. & Massei, G. Temporal and spatial patterns of defecation in wild boar. Wildl. Soc. Bull. 39, 65–69 (2015).

69. USGS. Freshwater withdrawals in the United States. https://www.usgs.gov/special-topics/water-science-school/science/freshwater-withdrawals-united-states (2018).

70. Smith, B. A., Fazil, A. & Lammerding, A. M. A risk assessment model for Escherichia coli O157:H7 in ground beef and beef cuts in Canada: Evaluating the effects of interventions. Food Control 29, 2 (2013).

71. Benjamin, L. et al. Occurrence of generic Escherichia coli, E. coli O157, and Salmonella spp. in water and sediment from leafy green produce farms and streams on the Central California coast. Int. J. Food Microbiol. 165, 65–76 (2013).

72. Stine, S. W. et al. Application of microbial risk assessment to the development of standards for enteric pathogens in water used to irrigate fresh produce. J. Food Prot. 68, 913–918 (2005).

73. Allende, A. et al. Quantitative contamination assessment of Escherichia coli in baby spinach primary production in Spain: Effects of weather conditions and agricultural practices. Int. J. Food Microbiol. 257, 238–246 (2017).

74. FDA. Standards for the Growing, Harvesting, Packing, and Holding of Produce for Human Consumption. Federal Register. https://www.federalregister.gov/documents/2015/11/27/2015-28159/standards-for-the-growing-harvesting-packing-and-holding-of-produce-for-human-consumption (2015).

75. Girardin, L. et al. Behaviour of the pathogen surrogates Listeria innocua and Clostridium sporogenes during production of parsley in fields fertilized with contaminated amendments. FEMS Microbiol. Ecol. 54, 287–295 (2005).

76. Alzahrani, A. Persister formation and revival in Shiga Toxin-Producing Escherichia coli subjected to environmental stress conditions. PhD Thesis, University of Guelph. https://atrium.lib.uoguelph.ca/server/api/core/bitstreams/e5a584c9-3e95-41a1-aa4a-7e0abf5e0c42/content (2020).

77. Hofsteenge, N., van Nimwegen, E. & Silander, O.K. Quantitative analysis of persister fractions suggests different mechanisms of formation among environmental isolates of E. coli. BMC Microbiol 13, 25 (2013).

78. Niu, H., Gu, J. & Zhang, Y. Bacterial persisters: molecular mechanisms and therapeutic development. Sig. Transduct. Target. Ther. 9, 174 (2024).

79. Wang, D. et al. Comparative persistence of Salmonella and Escherichia coli O157:H7 in loam or sandy loam soil amended with bovine or swine manure. Can. J. Microbiol. 64, 979–991 (2018).

80. Tran, D. T. Q. et al. Environmental drivers for persistence of Escherichia coli and Salmonella in manure-amended soils: A meta-analysis. J. Food Prot. 83, 1268–1277 (2020).

81. McKellar, R. C., LeBlanc, D. I., Rodríguez, F. P. & Delaquis, P. Comparative simulation of Escherichia coli O157:H7 behaviour in packaged fresh-cut lettuce distributed in a typical Canadian supply chain in the summer and winter. Food Control 35, 192–199 (2014).

82. McKellar, R. C. LeBlanc, D. I., Lu, J. & Delaquis, P. Simulation of Escherichia coli O157:H7 behavior in fresh-cut lettuce under dynamic temperature conditions during distribution from processing to retail. Foodborne Pathog. Dis. 9, 239–244 (2012).

83. When to harvest vegetables. University of Georgia Extension. https://extension.uga.edu/publications/detail.html?number=C935&title=when-to-harvest-vegetables (2022).

84. Keskinen, L. A., Burke, A. & Annous, B. A. Efficacy of chlorine, acidic electrolyzed water and aqueous chlorine dioxide solutions to decontaminate Escherichia coli O157:H7 from lettuce leaves. Int. J. Food Microbiol. 132, 134–140 (2009).

85. Zoellner, C., Wiedmann, M. & Ivanek, R. An assessment of Listeriosis risk associated with a contaminated production lot of frozen vegetables consumed under alternative consumer handling scenarios. J. Food Prot. 82, 2174–2193 (2019).

86. USDA. Lettuce, Cos or Romaine, Raw; Portion. https://fdc.nal.usda.gov/food-details/746769/nutrients/ (2015).

87. Cassin, M. H., Lammerding, A. M., Todd, E. C., Ross, W., & McColl, R. S. Quantitative risk assessment for Escherichia coli O157:H7 in ground beef hamburgers. Int. J. Food Microbiol. 41, 21–44 (1998).

88. U.S. Census Bureau. Census Bureau projects U.S. population of 312.8 million on New Year’s Day. https://www.census.gov/newsroom/releases/archives/population/cb11-219.html (2011).

89. Wickham, H., François, R., Henry, L., Müller, K. & Vaughan, D. dplyr: A Grammar of Data Manipulation. R package version 1.1.4. https://dplyr.tidyverse.org. (2023).

90. Akoglu, H. User’s guide to correlation coefficients. Turk. J. Emerg. Med. 18, 91–93 (2018).

91. Mukaka, M.M. Statistics corner: A guide to appropriate use of correlation coefficient in medical research. Malawi Med. J. 24, 69–71 (2012).

92. Therneau, T., Atkinson, B. & Ripley, B. rpart: Recursive Partitioning and Regression Trees. https://cran.r-project.org/package=rpart (2018).

93. Mir, R. et al. Cattle intestinal microbiota shifts following Escherichia coli O157:H7 vaccination and colonization. PLOS ONE 14, e0226099 (2019).

94. Chang, T. Evaluation of multiple disinfection methods to mitigate contaminated irrigation water. Master’s Thesis, University of Tennessee https://trace.tennessee.edu/utk_gradthes/3352 (2015).

95. Dery, J. L., Gerrity, D. & Rock, C. Minimizing Risks: Use of surface water in pre-harvest agricultural irrigation; part ii: sodium and calcium hypochlorite (chlorine) treatment methods. https://repository.arizona.edu/bitstream/handle/10150/669941/AZ1831-2020.pdf?sequence=1&isAllowed=y (2011).

96. Beauvais, W. et al. The effectiveness of treating irrigation water using ultraviolet radiation or sulphuric acid fertilizer for reducing generic Escherichia coli on fresh produce – a controlled intervention trial. J. Appl. Microbiol. 131, 1360–1377 (2021).

97. FAO/WHO. Joint FAO/WHO expert meeting on microbiological risk assessment on the prevention and control of microbiological hazards in fresh fruits and vegetables -part 4: commodity-specific interventions. https://www.who.int/publications/m/item/joint-fao-who-expert-meeting-on-microbiological-risk-assessment-on-the-prevention-and-control-of-microbiological-hazards-in-fresh-fruits-and-vegetables-part4 (2022).

98. FDA, University of California Davis. Head Lettuce. https://www.wifss.ucdavis.edu/wp-content/uploads/2016/10/HeadLettuce_PDF.pdf

99. EcoSure 2007 Cold Temperature Database. https://www.foodrisk.org/resources/display/21

100. Verrill, L. Consumer vegetable and fruit washing practices in the United States, 2006 and 2010. Food Prot. Trends 32, 164–172 (2012).

101. USDA ERS. Vegetables and Pulses Outlook (2023). Available at: https://downloads.usda.library.cornell.edu/usda-esmis/files/cj82k729x/pn89fk86d/3f463k190/VGS-370.pdf

102. Tack, D. M. et al. Shiga toxin-producing Escherichia coli outbreaks in the United States, 2010-2017. Microorganisms, 9, 1529 (2021).

103. Ivanek, R., Gröhn, Y. T., Wiedmann, M. & Wells, M. T. Mathematical model of Listeria monocytogenes cross-contamination in a fish processing plant. J. Food Prot. 67, 2688– 2697 (2004).

104. de Freitas Costa, E., Corbellini, L. G., da Silva, A.P.S.P. & Nauta, M. A stochastic model to assess the effect of meat inspection practices on the contamination of pig carcasses. Risk Anal. 37, 1849–1864 (2017).

105. Ivanek, R., Snary, E. L., Cook, A. J., & Gröhn, Y. T. A mathematical model for the transmission of Salmonella Typhimurium within a grower-finisher pig herd in Great Britain. J. Food Prot. 67, 2403–2409 (2004).

106. Karp, D. S. et al. Co-managing fresh produce for nature conservation and food safety. Proc. Natl. Acad. Sci. U. S. A. 112, 11126–11131 (2015).

107. Gelting, R. J. et al. Irrigation water issues potentially related to the 2006 multistate E. coli O157:H7 outbreak associated with spinach. Agric. Water Manag. 98, 1395–1402 (2011).

108. Bottichio, L. et al. Shiga toxin-producing Escherichia coli infections associated with romaine lettuce—United States, 2018. Clin. Infect. Dis. 71, e323–e330 (2020).

109. Gelting, R. J. et al. A systems analysis of irrigation water quality in an environmental assessment of an E. coli O157:H7 outbreak in the United States linked to iceberg lettuce. Agric. Water Manag. 150, 11–118 (2015).

110. FDA. Environmental Assessment Factors Potentially Contributing to Contamination of Romaine Lettuce Implicated in Outbreaks. (2019). Available at: https://www.fda.gov/food/outbreaks-foodborne-illness/environmental-assessment-factors-potentially-contributing-contamination-romaine-lettuce-implicated#summary

111. Allende, A. & Monaghan, J. Irrigation water quality for leafy crops: a perspective of risks and potential solutions. Int. J. Environ. Res. Public Health 12, 7457–7477 (2015).

112. Dery, J. L., Choppakatla, V., Sughroue, J. & Rock, C. Minimizing Risks: Use of Surface Water in Pre-Harvest Agricultural Irrigation; Part III: Peroxyacetic Acid (PAA) Treatment Methods. Available at: https://extension.arizona.edu/sites/extension.arizona.edu/files/pubs/az1884-2021.pdf

113. Banach, J. L. et al. Application of water disinfection technologies for agricultural waters. Agric. Water Manag. 244, 106527 (2021).

114. Collvignarelli, M. et al. Overview of the main disinfection processes for wastewater and drinking water treatment plants. Sustainability 10, 86 (2018).

115. Rock, C. CPS Rapid Response – Yuma Valley, Final Report. Center for Produce Safety (2019).

116. Rock, C., Cooper, K. & Yemmireddy, V. K. Agriculture Water Treatment – Southwest Region Final Project Report. Center for Produce Safety (2021).

117. USEPA. Peracetic Acid Fact Sheet https://www.epa.gov/sites/default/files/2019-08/documents/disinfection_-_paa_fact_sheet_-_2012.pdf (2012).

118. USEPA. Chlorination Fact Sheet. https://www3.epa.gov/npdes/pubs/chlo.pdf (1999).

119. USEPA. UV Disinfection Fact Sheet. https://www3.epa.gov/npdes/pubs/uv.pdf (1999).

120. Fonseca, J. M. et al. Escherichia coli survival in lettuce fields following its introduction through different irrigation systems. J. Appl. Microbiol. 110, 893–902 (2011).

121. Moyne, A. L. et al. Fate of Escherichia coli O157:H7 in field-inoculated lettuce. Food Microbiol. 28, 1417–1425 (2011).

122. Solomon, E. B., Potenski, C. J. & Matthews, K. R. Effect of irrigation method on transmission to and persistence of Escherichia coli O157:H7 on lettuce. J. Food Prot. 65, 673–676 (2002).

123. Gil, M. I., Tudela, J. A., Luna, M. C. & Allende, A. Water management and its effect on the postharvest quality of fresh-cut vegetables. Stewart Postharvest Rev. 9, 1–8 (2013).

124. Gurtler, J. B. & Gibson, K. E. Irrigation water and contamination of fresh produce with bacterial foodborne pathogens. Curr. Opin. Food Sci. 47, 100889 (2022).

125. Song, I., Stine, S., Choi, C. & Gerba, C. Comparison of crop contamination by microorganisms during subsurface drip and furrow irrigation. J. Environ. Eng. 132, 1243–1248 (2006).

126. Park, S. et al. Multifactorial effects of ambient temperature, precipitation, farm management, and environmental factors determine the level of generic Escherichia coli contamination on preharvested spinach. Appl. Environ. Microbiol. 81, 2635–2650 (2015).

127. Holvoet, K., Sampers, I., Seynnaeve, M., & Uyttendaele, M. Relationships among hygiene indicators and enteric pathogens in irrigation water, soil and lettuce and the impact of climatic conditions on contamination in the lettuce primary production. Int. J. Food Microbiol. 171, 21–31 (2014).

128. Erickson, M. C. et al. Preharvest internalization of Escherichia coli O157:H7 into lettuce leaves, as affected by insect and physical damage. J. Food Prot. 73, 1809–1816 (2010).

129. Petri, E., Rodríguez, M. & García, S. Evaluation of combined disinfection methods for reducing Escherichia coli O157:H7 population on fresh-cut vegetables. Int. J. Environ. Res. Public Health 12, 8678–8690 (2015).

130. Mokhtari, A. et al. Evaluation of chlorine dioxide and peracetic acid on Escherichia coli O157:H7 reduction on lettuce. Food Control 62, 36–40 (2016).

131. Fu, T.-J., Li, Y., Awad, D.Zhou, T.-Y. & Liu, L. Factors affecting the performance and monitoring of a chlorine wash in preventing Escherichia coli O157:H7 cross-contamination during postharvest washing of cut lettuce. Food Control 94, 212–221 (2018).

132. Gombas, D. et al. Guidelines to validate control of cross-contamination during washing of fresh-cut leafy vegetables. J. Food Prot. 80, 312–330 (2017).

133. Luo, Y. et al. Determination of free chlorine concentrations needed to prevent Escherichia coli O157:H7 cross-contamination during fresh-cut produce wash. J. Food Prot. 74, 352–358 (2011).

134. Gómez-López, V. M., Lannoo, A. S., Gil, M. I. & Allende, A. Minimum free chlorine residual level required for the inactivation of Escherichia coli O157:H7 and trihalomethane generation during dynamic washing of fresh-cut spinach. Food Control 42, 132–138 (2014).

135. Latorre, A. et al. Quantitative risk assessment of listeriosis due to consumption of raw milk. J. Food Prot. 74, 1268–1281 (2011).

136. Pouillot, R., Lubran, M. B., Cates, S. C. & Dennis, S. Estimating parametric distributions of storage time and temperature of ready-to-eat foods for U.S. households. J. Food Prot. 73, 312–321 (2010).

137. Climate Data Online. https://www.ncei.noaa.gov/cdo-web/

138. USDA. U.S. lettuce production shifts regionally by season. https://www.ers.usda.gov/data-products/charts-of-note/chart-detail?chartId=106516 (2023).

139. USDA ERS. Vegetables and pulses data - data by commodity - imports and exports. https://data.ers.usda.gov/reports.aspx?ID=4027&programArea=veg&top=5&HardCopy=True&RowsPerPage=25&groupName=Vegetables&commodityName=Lettuce#P910689dff5c747409da423e3f8f44e38_4_292 (2025).

140. National Advisory Committee on Microbiological Criteria for Foods (NACMSF). Response to questions posed by the food and drug administration regarding virulence factors and attributes that define foodborne shiga toxin-producing Escherichia coli (STEC) as severe human pathogens. J. Food Prot, 82, 724–767 (2019).

