## Supplemental Material for "Risk assessment of *Escherichia coli* O157:H7 along the farm-to-fork fresh-cut romaine lettuce supply chain"

**Author affiliations**

### Supplementary Table S1. Parameters in the preharvest *Escherichia coli* O157:H7 (ECO157) model

| Parameter | Description (units) | Distribution/Value | Mean (5th; 95th percentile) | Source |
|---|---|---|---|---|
| $t$ | Day of the countdown period in the difference equation model (1)-(6) (day) | $0 \leq t < Modeldays$ | - | Calculated [c] |
| $A_{field}$ | Area of the field (approximately 1-acre field) ($cm^2$) | $6245 \times 6245 = 39000025$ | - | *[36]* |
| $A_{batch}$ | Area of a field that grows a romaine batch ("batch area", 1/12 of the field area) ($cm^2$) | $6245 \times 6245/12 = 3247400$ | - | *[36]* |
| $N_{plant}$ | Number of plants harvested from the field (romaine plants) | 26,000 | - | *[36]* |
| $Plant_{width}$ | The area that a single mature romaine plant occupied on the field, length x width of one plant ($cm^2$) | $25 \times 25 = 625$ | - | *[51]* |
| $Plant_{space}$ | The area allocated for one romaine plant in the field ($cm^2$) | $30 \times 50 = 1500$ | - | *[36]* |
| $Plant_{barea}$ | Number of plants grown in a batch area (romaine plants) | $17 \times 125 = 2167$ | - | *[36]* |
| $Plant_{wght}$ | Weight of one mature romaine plant (g) | 300 | - | Assumption [c] |
| $f_{loc}$ | Select geographic location where the romaine batches were grown (unitless). Notations:<br>CC: Coastal Region and Central Valley, CA<br>DR: Desert (Imperial) Region, CA<br>Y: Yuma Region, AZ | $Discrete(\{CC, DR, Y\}, \{0.6, 0.1, 0.3\})$ | - | *[52,53,54]* |
| $f_{month}$ | Select the month of harvest for the given geographic location (unitless). Notations:<br>January: 1, February: 2, March: 3, April: 4, May: 5, June: 6, July: 7, August: 8, September: 9, October: 10, November: 11, December: 12 | $f_{month}(f_{loc} = CC)$ $= DUniform(\{4,5,6,7,8,9,10,11\})$<br>$f_{month}(f_{loc} = DR) = DUniform(\{1,2,3,12\})$<br>$f_{month}(f_{loc} = Y) = DUniform(\{1,2,3,12\})$ | - | *[52,53]* |
| $f_w$ | Select the source of irrigation water (unitless). Notations:<br>S: Surface water<br>W: Well water | $f_w(f_{loc} = CC)$ $= Discrete(\{s, w\}, \{0.11, 0.89\})$<br>$f_w(f_{loc} = DR)$ $= Discrete(\{s, w\}, \{0.30, 0.70\})$<br>$f_w(f_{loc} = Y)$ $= Discrete(\{s, w\}, \{0.90, 0.10\})$ | - | *[70]* |
| $f_{irr}$ | Select the irrigation type for the given geographic location (unitless). Notations:<br>O: Overhead spray irrigation<br>F: Furrow irrigation<br>D: Drip irrigation | $f_{irr}(f_{loc} = CC)$ $= Discrete(\{O, F, D\}, \{0.4, 0, 0.6\})$<br>$f_{irr}(f_{loc} = DR)$ $= Discrete(\{O, F, D\}, \{0.2, 0.5, 0.3\})$<br>$f_{irr}(f_{loc} = Y)$ $= Discrete(\{O, F, D\}, \{0.08, 0.92, 0\})$ | - | Author opinion [c] |
| $Modeldays$ | Countdown period before harvest, modeled via a set of difference equations (day) | 14 | - | *[39]* |
| $edge$ | Batch area was at the edge of the field and thus could be contaminated by runoff (unitless): no=0 (baseline), yes=1 | $Binomial\left(1, \frac{1}{12}\right)$ | $0.08\ (0; 1)$ | Calculated [c] |
| $C_{man}$ | ECO157 count per g of BSAAO (CFU/g) | $10^{Uniform(3.1, 8.4)}$ | $2.0 \times 10^7 (2.0 \times 10^3; 1.3 \times 10^8)$ | *[36,60]* |
| $Sc_{vac}$ | Cattle vaccination intervention against ECO157 (unitless): no=0 (baseline), yes=1 | 0 | - | *[93]* |
| $Rh_{con}$ | Volume of 1g BSAAO/swine feces/cattle feces ($cm^3$/g) | 2.5 | - | *[64]* |
| $Dh_{con}$ | Depth of tiling on the field (cm) | $Uniform(15,20)$ | $17.50\ (15.25; 19.75)$ | *[65]* |

| | | | | |
|---|---|---|---|---|
| $M_{man}$ | Mass of BSAAO applied on the batch area (g) | $\frac{Uniform(5000000,15000000)}{12}$ | $8.3 \times 10^5$ $(4.6 \times 10^5; 1.2 \times 10^6)$ | *[63]* |
| $DiF_{man}$ | Dilution of ECO157 cells in BSAAO in the volume of the tilled soil batch layer (unitless) | $\frac{M_{man} \times Rh_{con}}{A_{batch} \times Dh_{con}}$ | - | Calculated [c] |
| $Pr_{man}$ | BSAAO was used (unitless): no=0, yes=1 | 0.1 | - | Author opinion [c] |
| $P_{man}$ | BSAAO applied on a field was contaminated with ECO157 (unitless): no=0, yes=1 | $Binomial(1, Pert(0,0.17,0.21))$ | $0.15\ (0; 1)$ | *[60]* |
| $C_{man_P}$ | ECO157 count in any BSAAO applied soil, which returns 0 CFU for a non-contaminated soil and $C_{man}$ for a contaminated soil (CFU/g) | $P_{man} \times Pr_{man} \times C_{man}$ | - | Calculated [c] |
| $P_{irr_S}$ | ECO157 prevalence in surface water (unitless) | $Pert(0,0.08,0.21)$ | $0.09\ (0.03; 0.16)$ | *[8]* |
| $P_{irr_W}$ | ECO157 prevalence in well water (unitless) | 0.001 | - | Assumption [c] |
| $P_{irr}$ | Irrigation water contaminated with ECO157 applied during the countdown period (unitless): no=0, yes=1 | $Binomial(1, P_{irr})$, where<br>$P_{irr}(f_w = W) = P_{irr_W}$<br>$P_{irr}(f_w = S) = P_{irr_S}$ | $0.04\ (0; 0)$ | Calculated [c] |
| $Sc_{wt}$ | Water treatment intervention applied prior to the overhead spray irrigation (O) using surface water (S) (unitless): None=0 (baseline), Ultra-Violet (UV)=1, Peracetic Acid (PAA)=2, Chlorine=3 | 0 | - | Selected [c] |
| $C_{irr_S}$ | ECO157 count per ml of contaminated surface water post treatment (CFU/ml) | $\frac{10^{Pert(0.1,0.76,1.77)+Normal(-1.9,0.6,Truncate(,0))}}{10^{CC}}$ where<br>$CC(Sc_{wt} = 0) = 0$<br>$CC(Sc_{wt} = 1) = CC_{UV}$<br>$CC(Sc_{wt} = 2) = CC_{PPA}$<br>$CC(Sc_{wt} = 3) = CC_{Ch}$ | $0.08\ (0.007; 1.09)$ | *[19,20,71]* |
| $C_{irr_W}$ | ECO157 count per ml of contaminated well water (CFU/ml) | 0.1 | - | *[19,20,34]* |
| $C_{irr}$ | ECO157 count per ml of contaminated irrigation water applied on day t (CFU/ml) | $C_{irr}(f_w = W) = C_{irr_W}$<br>$C_{irr}(f_w = S) = C_{irr_S}$ | - | Calculated [c] |
| $C_{irr_P}$ | ECO157 count on any irrigation water, which returns 0 CFU for a non-contaminated water and $C_{irr}$ for a contaminated water (CFU/ml) | $P_{irr} \times C_{irr}$ | - | Calculated [c] |
| $type_{irr}$ | Probability the selected irrigation type was occurring on a given day (unitless) | $type_{irr}(f_{irr} = O) = fO_t = Binomial(1, Pr_O)$<br>$type_{irr}(f_{irr} = F) = fF_t = Binomial(1, Pr_F)$<br>$type_{irr}(f_{irr} = D) = fD_t = Binomial(1, Pr_{D_t})$ | - | Calculated [c] |
| $Pr_O$ | Probability of overhead spray irrigation on day t (unitless) | 0.4 | - | *[48]* |
| $Pr_F$ | Probability of furrow irrigation on day t (unitless) | 0.1 | - | Author opinion [c] |
| $Pr_{D_t}$ | Probability of drip irrigation on day t (unitless) | 1 | - | Author opinion [c] |
| $W_O$ | Volume of water applied through overhead spray irrigation to a romaine batch per an irrigation event (ml/day) | $\frac{Uniform(1850222756,2466963675) \times 0.51}{30 \times Pr_O \times 12}$ | $7.7 \times 10^6$ $(6.7 \times 10^6; 8.6 \times 10^6)$ | *[48]* |
| $W_F$ | Volume of water applied through furrow irrigation to a romaine batch per an irrigation event (ml/day) | $\frac{Uniform(2466963675,3083704593) \times 0.51}{30 \times Pr_F \times 12}$ | $3.9 \times 10^7$ $(3.5 \times 10^7; 4.3 \times 10^7)$ | *[48]* |
| $W_D$ | Volume of water applied through drip irrigation to a romaine batch per an irrigation event (ml/day) | $\frac{Uniform(1233481837,1850222756) \times 0.51}{30 \times Pr_{D_t} \times 12}$ | $2.2 \times 10^6$ $(1.8 \times 10^6; 2.6 \times 10^6)$ | *[48]* |

| | | | | |
|---|---|---|---|---|
| $Daytogrow$ | Days from seeding to harvest (day) | $Daytogrow(f_{loc} = CC) = Uniform(65{,}80)$<br>$Daytogrow(f_{loc} = DR) = 130$<br>$Daytogrow(f_{loc} = Y) = 130$ | 72 (65; 79)<br>-<br>- | *[48]* |
| $D_{man}$ | Number of days before BSAAO application (day) | $Daytogrow$ | - | *[19,20]* |
| $g_{irr}$ | Volume of water applied per an irrigation event in a batch area (ml/day) | $g_{irr}(f_{irr} = O) = W_O$<br>$g_{irr}(f_{irr} = F) = W_F$<br>$g_{irr}(f_{irr} = D) = W_D$ | - | Calculated [c] |
| $V_O$ | Volume of irrigation water captured per g of romaine per an overhead spray irrigation event (ml/g/day) | $Uniform(1.8{,}21.6)$ | 11.7 (2.8; 20.6) | *[27]* |
| $V_F$ | Volume of irrigation water captured per romaine batch per a furrow irrigation event (ml/day) | 100 | - | Assumption [c] |
| $V_D$ | Volume of irrigation water captured per romaine batch per a drip irrigation event (ml/day) | 10 | - | Assumption [c] |
| $v_{irr}$ | Volume of irrigation water captured by romaine batch per an irrigation event (ml/day) | $v_{irr}(f_{irr} = O) = V_O \times Plant_{barea} \times Plant_{wght}$<br>$v_{irr}(f_{irr} = F) = V_F$<br>$v_{irr}(f_{irr} = D) = V_D$ | - | Calculated [c] |
| $Tr_{man_O}$ | Fraction of the top 1cm layer of soil batch transferred to a romaine batch via splash caused by overhead spray irrigation per irrigation event (unitless) | $BetaGeneral(0.4{,}0.8{,}0.05{,}16.4) \times Plant_{barea} \times \frac{Plant_{wght}}{5} \times \frac{Rh_{con}}{A_{batch} \times 1}$ | 0.05 (0.0005; 0.13) | *[73]* |
| $Tr_{man_F}$ | Fraction of the top 1cm layer of soil batch transferred to a romaine batch via splash caused by furrow irrigation per irrigation event (unitless) | 0.0005 | - | Assumption [c] |
| $Tr_{man_D}$ | Fraction of the top 1cm layer of soil batch transferred to a romaine batch via splash caused by drip irrigation per irrigation event (unitless) | 0.0001 | - | Assumption [c] |
| $M_{irr}$ | Mass of soil transferred to a romaine batch via splash per irrigation event (g/day) | $M_{irr}(f_{irr} = O) = Tr_{man_O}$<br>$M_{irr}(f_{irr} = F) = Tr_{man_F}$<br>$M_{irr}(f_{irr} = D) = Tr_{man_D}$ | - | Calculated [c] |
| $Tr_{ecirr}$ | Fraction of ECO157 cells transferred from irrigation water to romaine (unitless) | 0.1 | - | Assumption [c] |
| $DiF_{irr}$ | Dilution factor for ECO157 cells in irrigation water in the top 1cm layer of soil batch (unitless) | $\frac{g_{irr} \times \left(1 - \frac{v_{irr}}{g_{irr}}\right)}{(A_{batch} \times 1)}$ | - | Calculated [c] |
| $LN_0$ | ECO157 count in a romaine batch at the start of the 14-day countdown period (t=0) (CFU) | $LN_0(f_{irr} = O) = 0$<br>$LN_0(f_{irr} = F) = 0$<br>$LN_0(f_{irr} = D) = 0$ | - | Assumption [c] |
| $CC_{UV}$ | $Log_{10}$ ECO157 count reduction of surface water contamination by UV treatment before overhead spray irrigation ($Log_{10}$CFU) | $Uniform(1.30{,}2.91)$ | 2.10 (1.38; 2.83) | *[94,96]* |
| $CC_{PAA}$ | $Log_{10}$ ECO157 count reduction of surface water contamination by PAA treatment before overhead spray irrigation ($Log_{10}$CFU) | $Uniform(2.48{,}3.09)$ | 2.78 (2.51; 3.06) | *[94,96]* |
| $CC_{Ch}$ | $Log_{10}$ ECO157 count reduction of surface water contamination by Chlorine treatment before overhead spray irrigation ($Log_{10}$CFU) | $Uniform(1.00{,}2.78)$ | 1.89 (1.09; 2.69) | *[94]* |
| $C_{cow_{prep}}$ | $Log_{10}$ ECO157 count on fresh cattle feces ($Log_{10}$CFU/g) | $Pert(0.89{,}3.08{,}8.4)$ | 3.60 (1.58; 6.04) | *[36]* |

| | | | | |
|---|---|---|---|---|
| $C_{cow_{tot}}$ | Log10 ECO157 count on fresh cattle feces, accounting for the effect of ECO157 vaccine intervention, if implemented (Log10CFU/g) | $C_{cow_{tot}} = Sc_{vac} \times Max(C_{cow} - 1,0) + (1 - Sc_{vac}) \times C_{cow}$ | - | Calculated [c] |
| $C_{cow}$ | ECO157 count on fresh cattle feces (CFU/g) | $10^{C_{cow_{tot}}}$ | - | Calculated [c] |
| $C_{cow_P}$ | ECO157 count on any cattle feces, which returns 0 CFU for non-contaminated feces and $C_{cow}$ for contaminated feces (CFU/g) | $C_{cow} \times P_{cow_m}$ | - | Calculated [c] |
| $C_{pig}$ | Log10 ECO157 count on fresh swine feces (Log10CFU/g) | $Pert(0.89,3.59,7.00)$ | 3.71 (1.86; 5.65) | *[36]* |
| $C_{pig_P}$ | ECO157 count on any swine feces, which returns 0 CFU for non-contaminated feces and $C_{pig}$ for contaminated feces (CFU/g) | $10^{C_{pig}} \times P_{pig_m}$ | - | Calculated [c] |
| $Pr_{pig}$ | Probability of feral swine entering the field on day t (unitless) | 0.00405 | - | *[6]* |
| $Pr_{pig_{Countt}}$ | Wildlife intrusion occurred on day t (unitless): no=0, yes=1 | $Binomial\ (1, Pr_{pig})$ | 0 (0; 1) | Calculated [c] |
| $Pr_{pig_{CountSum}}$ | Total number of days during the 14-day countdown when wildlife intrusion occurred (day) | $\sum_t Pr_{pig_{countt}}$ | - | Calculated [c] |
| $Mdef_{pig}$ | Mass of feces excreted by a feral swine per day (g swine feces/day) | 4,265 | - | *[36]* |
| $Tdef_{pig}$ | Number of times a feral swine defecated per day (times/day) | 4 | - | *[68]* |
| $Ndef_{pig}$ | Number of times feral swine (between 1-5 swine) defecated on the field (times/day) | $Poisson\big(1, Truncate(1,20)\big)$ | 1.58 (1; 3) | Assumption [c] |
| $M_{pig}$ | Mass of feces excreted by feral swine intruding the batch area (g) | $\frac{Mdef_{pig}}{Tdef_{pig}} \times Ndef_{pig}$ | - | Calculated [c] |
| $DiF_{pig}$ | Dilution factor for ECO157 cells in feral swine feces in soil batch associated with plants near which the swine defecated (unitless) | $\frac{M_{pig} \times Rh_{con}}{A_{batch} \times 1}$ | - | Calculated [c] |
| $M_{cow}$ | Mass of cattle runoff (carrying fresh cattle feces only) introduced into the edge batch area soil per runoff event (g/day) | $Triang(5000,10000,15000)$ | 10000 (6581; 13418) | *[36]* |
| $DiF_{cow}$ | Dilution factor for ECO157 cells in cattle runoff introduced into in the top 1cm layer of soil batch (unitless) | $\frac{M_{cow} \times Rh_{con}}{A_{batch} \times 1}$ | - | Calculated [c] |
| $Pr_{LtoS}$ | Proportion of fecal matter (swine and runoff) landing on romaine rather than in soil (unitless) | $\frac{Plant_{width}}{Plant_{space} - Plant_{width}}$ | - | *[36]* |
| $Tr_{ecsoil}$ | Proportion of ECO157 cells in soil transferring to romaine during a rain or irrigation event (unitless) | $Uniform(0.00004,0.63)$ | 0.315 (0.032; 0.600) | *[75]* |
| $R_{dman}$ | ECO157 decay rate for non-persisters in BSAAO amended soil (Log10CFU/day)[a] | 0.038 | - | *[61]* |
| $R_{dL}$ | ECO157 decay rate for non-persisters on romaine in field (1/day) [a,b] | $1 - 10^{h_{sun_m} \times \frac{R_{sun_m}}{24}}$ | - | *[27,34]* |
| $R_{ds}$ | ECO157 decay rate for non-persisters in soil (1/day) [a] | $1 - 10^{-0.1744}$ | - | *[36]* |
| $R_{dman_P}$ | ECO157 decay rate for persisters in BSAAO amended soil (Log10CFU/day) [a] | $\frac{R_{dman}}{10}$ | - | *[61,62]* |
| $R_{dL_P}$ | ECO157 decay rate for persisters on romaine in field (1/day) [a] | $\frac{1 - 10^{h_{sun_m} \times \frac{R_{sun_m}}{24}}}{10}$ | - | *[47]* |
| $R_{ds_P}$ | ECO157 decay rate in soil for persisters (1/day) [a] | $\frac{R_{ds}}{10}$ | - | *[62]* |

| | | | | |
|---|---|---|---|---|
| $NtoP_{man}$ | Switch rate from normal to persister in BSAAO amended soil (1/day) [a] | 0.001 | - | *[76,77]* |
| $NtoP_L$ | Switch rate from normal to persister on romaine (1/day) [a] | 0.0004 | - | *[47]* |
| $NtoP_S$ | Switch rate from normal to persister in soil (1/day) [a] | 0.0001 | - | *[76,77]* |
| $Pmax_{man}$ | Maximum fraction of persisters in BSAAO amended soil (unitless) | $\frac{NtoP_{man}}{10^{R_{dman}}}$ | - | *[47]* |
| $Pmax_S$ | Maximum fraction of persisters in soil (unitless) | $\frac{NtoP_S}{R_{ds}}$ | - | Calculated |
| $Pmax_L$ | Maximum fraction of persisters on romaine (unitless) | $\frac{NtoP_L}{R_{dL}}$ | - | Calculated |
| $BwaitP$ | Waiting time: The period before harvest during which overhead spray and furrow irrigation was stopped (day) | $Triang(2,4,8)$ | $4\ (2;6)$ | *[74]* |
| $Tr_{blade}$ | Average amount of soil attached to a harvesting blade after contact with soil in one cross contamination event (g/blade) | 10.22 | - | *[15]* |
| $Tr_{ecblade}$ | Proportion of ECO157 cells transferred from a harvesting blade to a romaine plant in one cross contamination event (unitless) | 0.0013 | - | *[15]* |
| $N_{blade}$ | ECO157 count in soil attached on blades (CFU/soil batch x soil batch/blade=CFU/blade) | $\frac{LN_{14} \times Tr_{blade} \times Rh_{con}}{1 \times A_{batch}}$ | - | Calculated [c] |
| $CC_{blade}$ | ECO157 cells transferred from harvesting blades to a romaine batch (CFU/romaine batch) | $Tr_{ecblade} \times N_{blade} \times Plant_{barea} \times Plant_{wght} \times \frac{1}{3}$ | - | Calculated [c] |
| $C_{Lh}$ | Preharvest outcome: ECO157 count on harvested romaine batch (CFU) | $LN_{14} + CC_{blade}$ | - | Calculated [c] |

[a] In the difference equation model the rate parameter indicates the proportionate change in ECO157 population per time step.
[b] See Supplementary Table S3 for more details.
[c] Assumption: Parameter value accepted as true with little to no information; Author opinion: Parameter value accepted as true based on authors' experience; Calculated: Parameter value calculated based on other parameters; Selected: Used in scenario analysis (See Supplementary Table S9 for details for their use.)

### Supplementary Table S2. Parameters in the postharvest *Escherichia coli* O157:H7 (ECO157) model

| Parameter | Description (units) | Distribution/Value | Mean (5th-95th percentile) | Source |
|---|---|---|---|---|
| $C_{Lh_f}$ | *E. coli* O157:H7 (ECO157) count in a contaminated harvested romaine batch ($Log_{10}$CFU/romaine batch) | $Triang(-0.66,3.81,6.36)$ | $3.17\ (0.59;5.41)$ | Output from preharvest model fitted in @RISK |
| $P_{Lh_f}$ | ECO157 prevalence in a harvested romaine batch (unitless) | $Triang(0.005,0.01,0.02)$ | $0.01\ (0.007;0.02)$ | Output from preharvest model fitted in @RISK |
| $t_{coldst}$ | Cold storage time (h) | $Triang(1,24,72)$ | $32\ (10;59)$ | Author opinion [a] |
| $T_{endcoldst}$ | Temperature after cold storage (°C) | $Triang(0,4.44,T_{harvest_m})$ | $5\ (4.3;5.7)$ | *[33]* |
| $T_{coldst}$ | Temperature during cold storage (°C) | $\frac{1}{2} \times (T_{endcoldst} + T_{harvest_m})$ | - | *[38]* |
| $G_{coldst}$ | Cold storage growth rate ($Log_{10}$CFU/h) | $\frac{(0.023 \times (T_{coldst} - 1.2))^2}{2.303}$ | - | *[38]* |

| | | | | |
|---|---|---|---|---|
| $D_{coldst}$ | Cold storage die-off rate ($Log_{10}$CFU/h) | $\frac{Lognorm(0.013,0.001,Shift(0.001))}{2.303}$ | 0.006 (0.005; 0.007) | *[38]* |
| $GorD_{coldst}$ | Growth (G) or die off (D) based on cold storage temperature ($Log_{10}$CFU/h) | $G, if\ T_{coldst} > 5,$ $D, if\ T_{coldst} \leq 5$ | - | *[38]* |
| $CC_{coldst}$ | Change in ECO157 count during cold storage ($Log_{10}$CFU) | $CC_{coldst}(GorD_{coldst} = G) = G_{coldst} \times t_{coldst}$ $CC_{coldst}(GorD_{coldst} = D) = -D_{coldst} \times t_{coldst})$ | - | Calculated [a] |
| $C_{endcoldst}$ | ECO157 count on a contaminated romaine batch after cold storage, going to processing (CFU/romaine batch) | $\mathrm{Log}_{10}(ROUNDDOWN\left(10^{Max\{C_{Lh_f} + CC_{coldst},0\}}, 0\right)$ | - | Calculated [a] |
| $B$ | Romaine batch size (g) | $Plant_{barea} \times Plant_{wght}$ | - | Calculated [a] |
| $Sc_{wash}$ | 1 or 2 based on the scenarios below, (baseline=1) | 1 | - | Selected [a] |
| $CC_{ch20}$ | $Log_{10}$CFU ECO157 reduction with chlorine washing ($Log_{10}$CFU/g romaine) | $Pert(0.01,0.52,1.03)$ | 0.52 (0.20; 0.84) | *[84]* |
| $CC_{ch1}$ | Chlorine wash alternative 1 | $Pert(0.5,1.0,1.5)$ | 1.00 (0.69; 1.31) | Assumption [a] |
| $CC_{ch2}$ | Chlorine wash alternative 2 | $Pert(1.0,1.5,2.0)$ | 1.50 (1.19; 1.81) | Assumption [a] |
| $CC_{ch3}$ | Chlorine wash alternative 3 | $Pert(1.5,2.0,2.5)$ | 2.00 (1.69; 2.31) | Assumption [a] |
| $CC_{ch4}$ | Chlorine wash alternative 4 | $Pert(2.0,2.5,3.0)$ | 2.50 (0.69; 1.31) | Assumption [a] |
| $CC_{ch5}$ | Chlorine wash alternative 5 | $Uniform(3.0,5.0)$ | 4.00 (3.10; 4.90) | *[97]* |
| $CC_{w_{agent}}$ | ECO157 reduction associated with the scenario selected for the washing step ($Log_{10}$CFU) | $CC_{w_{agent}}(Sc_{wash} = 1) = CC_{ch20}$ $CC_{w_{agent}}(Sc_{wash} = 2) = CC_{ch1}$ $CC_{w_{agent}}(Sc_{wash} = 3) = CC_{ch2}$ $CC_{w_{agent}}(Sc_{wash} = 4) = CC_{ch3}$ $CC_{w_{agent}}(Sc_{wash} = 5) = CC_{ch4}$ $CC_{w_{agent}}(Sc_{wash} = 6) = CC_{ch5}$ | - | Selected [a] |
| $C_{wash}$ | ECO157 count on a contaminated romaine batch after washing (CFU/romaine batch) | $\mathrm{Log}_{10}(ROUNDDOWN(Max\{10^{C_{endcoldst} - CC_{w_{agent}}},0\}, 0))$ | - | Calculated [a] |
| $P0_{pack}$ | Probability a non-contaminated package is partitioned from a contaminated romaine batch (unitless) | $EXP\left(\frac{\Gamma(Clstr_{par} \times A_{pack}) \times \Gamma(Clstr_{par} \times (A_{pack} - 1) + C_{wash})}{\Gamma\left(Clstr_{par} \times (A_{pack} - 1)\right) \times \Gamma(Clstr_{par} \times A_{pack} + C_{wash})}\right)$ | - | *[46]* |
| $P_{pack}$ | ECO157 prevalence on a romaine package (unitless) | $(1 - P0_{pack}) \times P_{Lh_f}$ | 0.003 ($2.5 \times 10^{-5}$; 0.016) | Calculated [a] |
| $Sc_{clstrb}$ | Clustering in romaine batch: no=0 (baseline), yes=1 | 0 | - | Selected [a] |
| $Clstr_{parb}$ | Clustering parameter | 1 | - | *[46]* |
| $W_{romainepack}$ | Weight of shredded romaine per package (8 oz) (g) | 225 | - | |
| $A_{pack}$ | Number of packages in a romaine batch (packages) | $\frac{B}{W_{lettucepack}}$ | - | Calculated [a] |
| $P_{contpack_{dist}}$ | Distribute ECO157 cells to a package when clustering | $Beta\left(Clstr_{parb}, Clstr_{parb} \times (A_{pack} - 1)\right)$ | $3.5 \times 10^{-4}$ ($1.8 \times 10^{-5}$; 0.001) | Calculated [a] |
| $P_{contpack}$ | Proportion of ECO157 cells from a contaminated romaine batch in a package (unitless) | $(1 - Sc_{clstrb}) \times \left(\frac{1}{A_{pack}}\right) + Sc_{clstrb} \times P_{contpack_{dist}}$ | - | *[46]* |
| $C_{pack}$ | ECO157 count in a contaminated package ($Log_{10}$CFU/pack romaine) | $Log_{10}\left(Binomial(C_{wash}, P_{contpack}) + 1\right)$ | - | Calculated [a] |
| $t_{retail}$ | Retail storage time (h) | $Triang(0.5,4,7) \times 24$ | 92.0 (37.6; 144.3) | *[42]* |
| $T_{retail}$ | Retail storage temperature (°C) | $Normal\left(4.4441,2.9642,Truncate(0,20.56)\right)$ | 4.9 (0.9; 9.4) | *[82,99]* |
| $G_{retail}$ | Retail growth rate ($Log_{10}$CFU/g romaine/h) | $\frac{\left(0.023 \times (T_{trnspt} - 1.2)\right)^2}{2.303}$ | - | *[38]* |

| | | | | |
|---|---|---|---|---|
| $D_{retail}$ | Retail die-off rate ($Log_{10}$CFU/g romaine/h) | $\frac{Lognorm(0.013,0.001,Shift(0.001))}{2.303}$ | 0.006 (0.005; 0.007) | *[82]* |
| $GorD_{retail}$ | Growth (G) or die off (D) based on retail temperature ($Log_{10}$CFU/h) | $G, if\ T_{retail} > 5,$ $D, if\ T_{retail} \leq 5$ | - | *[38]* |
| $CC_{retail}$ | Change in ECO157 count during retail storage ($Log_{10}$CFU) | $CC_{retail}(GorD_{retail} = G) = G_{retail} \times t_{retail}$ $CC_{retail}(GorD_{retail} = D) = -D_{retail} \times t_{retail}$ | - | *[38]* |
| $C_{retail}$ | ECO157 count in a contaminated package after retail storage ($Log_{10}$CFU/package romaine) | $\mathrm{Log}_{10}(ROUNDDOWN\left(10^{Max\{C_{pack}+CC_{retail},0\}}, 0\right))$ | - | Calculated [a] |
| $t_{transport}$ | Transportation time (h) | $Normal(1.421,0.46478, Truncate(0.1833,3.8667), Shift(-0.24609))$ | 1.18 (0.43; 1.94) | *[99]* |
| $T_{trhom}$ | Package temperature before putting into home refrigerator (°C) | $Normal(8.386,3.831, Truncate(0,20))$ | 8.51 (2.54; 14.67) | *[99]* |
| $T_{trnspt}$ | Temperature during transportation (°C) | $\frac{1}{2} \times (T_{retail} + T_{home})$ | - | *[135]* |
| $G_{trnspt}$ | Transportation growth rate ($Log_{10}$CFU/) | $\frac{\left(0.023 \times (T_{trnspt} - 1.2)\right)^2}{2.303}$ | - | *[38]* |
| $D_{trnspt}$ | Transportation die-off rate ($Log_{10}$CFU/h) | $\frac{Lognorm(0.013,0.001,Shift(0.001))}{2.303}$ | 0.006 (0.005; 0.007) | *[82]* |
| $GorD_{trnspt}$ | Growth (G) or die off (D) based on transport temperature | $G, if\ T_{trnspt} > 5,$ $D, if\ T_{trnspt} \leq 5$ | - | *[38]* |
| $CC_{trspt}$ | Change in ECO157 count during transportation ($Log_{10}$CFU) | $CC_{trnspt}(GorD_{trnspt} = G) = G_{trnspt} \times t_{trnspt}$ $CC_{trnspt}(GorD_{trnspt} = D) = -D_{trnspt} \times t_{trnspt}$ | - | *[38]* |
| $C_{trnspt}$ | ECO157 count in a contaminated package after transportation ($Log_{10}$CFU/romaine package) | $\mathrm{Log}_{10}(ROUNDDOWN(10^{Max\{C_{retail}+CC_{trnspt},0\}}, 0))$ | - | Calculated [a] |
| $t_{fcons}$ | Time to first consumption (h) | $Weibull(1.13,2.84) \times 24$ | 65.21 (4.92; 179.97) | *[136]* |
| $t_{lcons}$ | Time to last consumption (h) | $Weibull(1.7,7.96) \times 24$ | 170.5 (33.3; 364.3) | *[136]* |
| $t_{home}$ | Home storage time (h) | $\frac{1}{2} \times (t_{fcons} + t_{lcons})$ | - | Calculated [a] |
| $T_{home}$ | Home storage temperature (°C) | $Normal\left(3.4517,2.4442, RiskTruncate(-5,17.22)\right)$ | 3.45 (−0.56; 7.47) | *[38]* |
| $G_{home}$ | Home growth rate ($Log_{10}$CFU/h) | $\frac{\left(0.023 \times (T_{home} - 1.2)\right)^2}{2.303}$ | - | *[38]* |
| $D_{home}$ | Home die-off rate ($Log_{10}$CFU/h) | $\frac{Lognorm(0.013,0.001,Shift(0.001))}{2.303}$ | 0.006 (0.005; 0.007) | *[82]* |
| $GorD_{home}$ | Growth (G) or die off (D) based on home storage temperature | $G, if\ T_{home} > 5,$ $D, if\ T_{home} \leq 5$ | - | *[38]* |
| $CC_{home}$ | Change in ECO157 count during home storage ($Log_{10}$CFU) | $CC_{home}(GorD_{home} = G) = G_{home} \times t_{home}$ $CC_{home}(GorD_{home} = D) = -D_{home} \times t_{home}$ | - | *[38]* |
| $C_{home}$ | ECO157 count in a contaminated package after home storage ($Log_{10}$CFU/romaine package) | $\mathrm{Log}_{10}(ROUNDDOWN(10^{Max\{C_{trnspt}+CC_{home},0\}}, ($ | - | Calculated [a] |
| $P0_{ser}$ | Probability a non-contaminated serving was partitioned from a contaminated romaine package | $EXP\left(\frac{\Gamma(Clstr_{par} \times Ser_{pack}) \times \Gamma(Clstr_{par} \times (Ser_{pack} - 1) + C_{home})}{\Gamma(Clstr_{par} \times (Ser_{pack} - 1)) \times \Gamma(Clstr_{par} \times Ser_{pack} + C_{home})}\right)$ | - | Calculated [a] |
| $P_{ser}$ | ECO157 prevalence on a romaine serving | $(1 - P0_{ser}) \times P_{pack} \times P_{Lh_f}$ | 0.002 ($1.8 \times 10^{-6}$; 0.009) | Calculated [a] |
| $Sc_{clstrp}$ | Clustering in package: no=0 (baseline), yes=1 | 0 | - | Selected [a] |
| $Clstr_{parp}$ | Clustering parameter | 1 | - | *[46]* |

| | | | | |
|---|---|---|---|---|
| $Ser$ | Serving size (g romaine/ romaine serving) | 85 | - | *[86]* |
| $Ser_{pack}$ | Number of servings in a package ( romaine servings) | $\frac{W_{lettucepack}}{Ser}$ | - | Calculated [a] |
| $P_{contser_{prep}}$ | Distribute ECO157 cells to a serving when clustering (unitless) | $Beta\left(Clstr_{parp}, Clstr_{parp} \times \left(Ser_{pack} - 1\right)\right)$ | 0.38 (0.031; 0.838) | Calculated [a] |
| $P_{contser}$ | Proportion of ECO157 cells from contaminated package in a serving (unitless) | $(1 - Sc_{clstrp}) \times \left(\frac{1}{Ser_{pack}}\right) + Sc_{clstrp} \times P_{contser_{prep}}$ | - | *[46]* |
| $C_{serH}$ | ECO157 count in a contaminated serving after home storage ($Log_{10}$CFU/serving romaine) | $Log_{10}(Binomial(10^{C_{home}}, P_{contser}) + 1)$ | - | Calculated [a] |
| $Sc_{conw}$ | Consumer wash romaine before consumption: no=0 (baseline), yes=1 | 0 | - | Selected [a] |
| $P_{conw}$ | Probability a USA consumer washes the prepacked romaine (unitless) | 0.62 | - | *[100]* |
| $CC_{conw_{prep}}$ | ECO157 reduction by consumer wash with water prep cell ($Log_{10}$CFU) | $Triang(0.65, 0.99, 0.99) \times P_{conw}$ | 0.54 (0.45; 0.61) | *[39]* |
| $CC_{conw}$ | $Log_{10}$CFU ECO157 reduction by consumer wash with water ($Log_{10}$CFU) | $Sc_{conw} \times CC_{conw_{prep}}$ | - | Calculated [a] |
| $C_{serW}$ | ECO157 count in a contaminated serving after consumer wash (CFU/romaine serving) | $10^{Max\{C_{serH} - CC_{conw}, 0\}}$ | - | Calculated [a] |
| $alpha$ | Dose-response parameter alpha (unitless) | 0.267 | - | *[87]* |
| $beta$ | Dose-response parameter beta (unitless) | 229.2928 | - | *[87]* |
| $Pill$ | Probability of illness as a result of consumption of a contaminated romaine serving (unitless) | $1 - \left(1 + \frac{C_{serW}}{beta}\right)^{-alpha}$ | - | *[36]* |
| $USpop$ | USA population (person) | 311,556,874 | - | *[88]* |
| $A$ | Annual romaine consumption in the USA (g/consumer/year) | 3,946 | - | *[18]* |
| $Np$ | Number of romaine servings consumed per person per year (romaine serving/consumer/year) | $\frac{A}{Ser}$ | - | Calculated [a] |
| $Nsc_{US}$ | Total number of romaine servings consumed per year in the USA (romaine serving/year) | $USpop \times Np$ | - | Calculated [a] |
| $Ncsc_{US}$ | Number of contaminated romaine servings consumed per year in the USA (romaine serving/year) | $Nsc_{US} \times P_{ser}$ | - | Calculated [a] |
| $Ncase_{US}$ | Number of illness cases per year in the USA (person) | $Ncsc_{US} \times Pill$ | - | Calculated [a] |

[a] Assumption: Parameter value accepted as true with little to no information; Author opinion: Parameter value accepted as true based on authors' experience; Calculated: Parameter value calculated based on other parameters; Selected: Used in scenario analysis (See Supplementary Table S9 for details for their use.)

### Supplementary Table S3. Parameters varying by the harvest month of romaine production

| Parameter | $P_{sun_m}$ | $P_{cow_m}$ | $P_{pig_m}$ | $h_{sun_m}$ | $R_{sun_m}$ | $Prec_{norm_m}$ | $Prec_{actual_m}$ | $T_{harvest_m}$ |
|---|---|---|---|---|---|---|---|---|
| **Description (unit)** | Probability of a sunny day (unitless) | ECO157 prevalence in fresh cattle feces (unitless) | ECO157 prevalence in fresh feral swine feces (unitless) | Average sunny hours per day (h)[e] | Solar decay rate (1/day) | Precipitation level normalized (unitless) | Actual (average) level of precipitation during the year (mm) | Temperature at harvest (°C) [d] |
| **Source** | *[137]* | *[8]* | *[8]* | *[27,34]* | *[27,34]* | *[137]* | *[137]* | *[137]* |
| **Harvest month** [a] | **Value/Distribution** | | | | | | | |
| January | 0.97 | $Binomial(1,0.01)$ | 0 [c] | $DR = 10.30$ $Y = 9.00$ | $-0.48$ | 0.21 | 11 | $DR = 7.20$ $Y = 8.90$ |
| February | 0.93 | $Binomial(1,0.02)$ | 0 [c] | $DR = 11.00$ $Y = 9.70$ | $-0.48$ | 0.31 | 15 | $DR = 7.20$ $Y = 9.90$ |
| March | 0.97 | $Binomial(1,0.06)$ [c] | 0 [c] | $DR = 12.00$ $Y = 10.70$ | $-0.48$ | 0.12 | 7 | $DR = 12.70$ $Y = 12.90$ |
| April | 0.87 | $Binomial(1,0.09)$ | 0 [c] | $CC = 8.90$ | $-0.48$ | 0.81 | 36 | $CC = 8.20$ |
| May | 0.90 | $Binomial(1,0.04)$ | 0 [c] | $CC = 9.10$ | $-0.48$ | 0.41 | 19 | $CC = 9.70$ |
| June | 0.97 | $Binomial(1,0.13)$[b] | 0 [c] | $CC = 9.80$ | $-0.52$ | 0.07 | 5 | $CC = 11.40$ |
| July | 1.00 | $Binomial(1,0.21)$ | $Binomial(1,0.16)$ | $CC = 9.60$ | $-0.52$ | 0.00 | 2 | $CC = 13.00$ |
| August | 1.00 | $Binomial(1,0.10)$ | $Binomial(1,0.13)$ | $CC = 9.10$ | $-0.52$ | 0.02 | 3 | $CC = 13.30$ |
| September | 0.97 | $Binomial(1,0.08)$ | $Binomial(1,0.11)$ | $CC = 9.10$ | $-0.52$ | 0.02 | 3 | $CC = 13.10$ |
| October | 0.93 | $Binomial(1,0.17)$ | $Binomial(1,0.08)$ | $CC = 8.50$ | $-0.52$ | 0.41 | 19 | $CC = 11.20$ |
| November | 0.87 | $Binomial(1,0.03)$ | $Binomial(1,0.10)$ | $CC = 7.60$ | $-0.52$ | 1.00 | 44 | $CC = 8.00$ |
| December | 0.97 | $Binomial(1,0.02)$ | $Binomial(1,0.12)$ | $DR = 10.00$ $Y = 8.80$ | $-0.48$ | 0.21 | 11 | $DR = 7.20$ $Y = 8.30$ |

[a] Selected month is based on the location of the romaine production.
[b] Empirical data was 0. However, the value considered here was the average of estimates from the prior and the following months.
[c] Empirical data was 0.
[d] CC: Coastal Region and Central Valley, CA; DR: Desert Region (Imperial), CA; Y:Yuma Region, AZ.

**Supplementary Table S4. Terms in in the difference equation model in the preharvest model.**

| Parameter | Description (units) | Distribution/Value |
|---|---|---|
| $P_{Sinitial}$ | Soil batch contamination on day t=0 (day 0, a day before the start of the 14-day countdown): no=0, yes=1 | $P_{man} \times Pr_{man}$ |
| $S_0$ | ECO157 count in soil batch on day t=0 (CFU) | $\frac{\left(Sc_{vac} \times \frac{C_{man}}{10} + (1 - Sc_{vac}) \times C_{man}\right) \times DiF_{man}}{10^{D_{man} \times R_{dman}}}$ |
| $SN_0$ | Non-persister ECO157 count in soil batch on day t=0 (CFU) | $S_0 \times (1 - Pmax_{man})$ |
| $SP_0$ | Persister ECO157 count in soil batch on day t=0 (CFU) | $S_0 \times Pmax_{man}$ |
| $C_{S_0}$ | Soil batch contamination with non-persister and persister cells on day t=0 (CFU) | $(SN_0 + SP_0) \times P_{Sinitial}$ |
| $LN_0$ | Non-persister ECO157 count in romaine batch on day t=0 in an overhead spray/furrow or drip irrigated field (CFU) | 0 |
| $LP_0$ | Persister ECO157 count in romaine batch on day t=0 in an overhead spray/furrow or drip irrigated field (CFU) | 0 |
| $C_{L_0}$ | Romaine batch contamination with non-persister and persister cells on day t=0 (CFU) | $LN_0 + LP_0$ |
| $SW$ | ECO157 count introduced via feral swine to soil batch per intrusion event (CFU) | $(1 - Pr_{LtoS}) \times 10^{C_{pig}} \times DiF_{pig}$ |
| $LW$ | ECO157 count introduced via feral swine to romaine batch per intrusion event (CFU) | $Pr_{LtoS} \times 10^{C_{pig}}$ |
| $LsR$ | Fraction of ECO157 transferred from soil batch to romaine batch via splash during rain per rain event (unitless) | $Tr_{man_O} \times Tr_{ecsoil} \times Prec_{norm_m}$ |
| $LsI$ | Fraction of ECO157 transferred from soil batch to romaine batch via splash during irrigation per irrigation event (unitless) | $M_{irr} \times Tr_{ecsoil} \times Median(Prec_{norm_m})$ |
| $SR$ | ECO157 introduced to batch area (soil + romaine) with runoff per runoff event (CFU) | $C_{cow} \times Prec_{norm_m}$ |
| $SSR$ | ECO157 count in soil batch introduced with runoff per runoff event (CFU) | $SR \times (1 - Pr_{LtoS}) \times DiF_{cow}$ |
| $LSR$ | ECO157 count in romaine batch introduced with runoff per runoff event (CFU) | $SR \times Pr_{LtoS}$ |
| $Bs_t$ | Select weather on day t: sunny=1, rainy=0 (Overhead spray or furrow irrigation may occur on a sunny day only) [a] | $Binomial\left(1, P_{sun_m}\right)_t$ |
| $fO_t$ | Overhead spray irrigation scheduled to occur on day t: no=0, yes=1 | $Binomial(1, Pr_O)_t$ |
| $fF_t$ | Furrow irrigation scheduled to occur on day t: no=0, yes=1 | $Binomial(1, Pr_F)_t$ |
| $SI_t$ | ECO157 count in soil batch introduced via irrigation per irrigation event (CFU) | $C_{irr} \times DiF_{irr}$ |
| $pVL$ | Fraction of irrigation water caught by romaine (unitless) | $\frac{v_{irr}}{g_{irr}}$ |
| $LI_t$ | ECO157 count in romaine batch via irrigation per irrigation event (CFU) | $C_{irr} \times g_{irr} \times Tr_{ecirr} \times pVL$ |
| $pRunoff_t$ | Runoff event occurs on day t: no=0, yes=1 | $(1 - Bs_t) \times P_{cow_m} \times edge$ |
| $Pr_{pig_{countt}}$ | Wildlife intrusion occurs on day t: no=0, yes=1 | $Binomial\ (1, Pr_{pig})$ |
| $Pr_{pigCountSum}$ | Total number of days during the 14-day countdown when wildlife intrusion occurs | $\sum_t Pr_{pig_{countt}}$ |
| $pPEC_t$ | Wildlife intrusion occurs by feral swine with contaminated feces on day t: no=0, yes=1 | $Binomial\ (1, P_{pig_m}) \times Pr_{pig_{countt}}$ |
| $BwaitP_t$ | Overhead spray or furrow irrigation occurs on day t: no=0, yes=1 | $IF(t > BwaitP, 1{,}0)$, where $t = \{0{,}1, \dots, 14\}$ |
| $pSpl_t$ | Irrigation splash occurs on day t: no=0, yes=1 | $IF(OR(Bs_t \times fOt \times BwaitP_t = 1, Bs_t \times fFt \times BwaitP_t = 1, fDt = 1), 1{,}0)$ |
| $pSpR_t$ | Rain splash occurs on day t: no=0, yes=1 | $1 - Bs_t$ |

[a] See Supplementary Table S3 for more details.

**Supplementary Table S5. Difference equation model in the preharvest model.**

| Compartment | Description (units) | Equation |
|---|---|---|
| $Irr_{S_t}$ | ECO157 count in soil batch introduced via irrigation on day t (CFU) | One of (depending on the irrigation method for the iteration: O=overhead spray, F=furrow, D=drip):<br>$Irr_S(f_{irr} = O)_t = SI_t \times Bs_t \times P_{irr} \times fO_t \times BwaitP_t$<br>$Irr_S(f_{irr} = F)_t = SI_t \times Bs_t \times P_{irr} \times fF_t \times BwaitP_t$<br>$Irr_S(f_{irr} = D)_t = SI_t \times P_{irr} \times Pr_D$ |
| $Pig_{S_t}$ | ECO157 count in soil batch introduced via feral swine on day t (CFU) | $SW \times pPEC_t$ |
| $Runoff_{S_t}$ | ECO157 count in soil batch introduced via runoff on day t (CFU) | $SSR \times pRunoff_t$ |
| $R_{splash_t}$ | Fraction of ECO157 transferred from soil batch to romaine batch via rain splash on day t (unitless) | $LsR \times pSpR_t$ |
| $I_{splash_t}$ | Fraction of ECO157 transferred from soil batch to romaine batch via irrigation splash on day t (unitless) | $LsI \times pSpl_t$ |
| $SNtoSP_t$ | Switch of non-persister ECO157 to persister cells on day t (CFU) | $(1 - Pr_{man}) \times SN_t \times NtoP_S + Pr_{man} \times SN_t \times NtoP_{man}$ |
| $SNdecay_t$ | Decay of non-persister ECO157 in soil batch on day t (CFU) | $(1 - Pr_{man}) \times SN_t \times R_{ds} + Pr_{man} \times \frac{SN_t}{10^{R_{dman}}}$ |
| $SPdecay_t$ | Decay of persister ECO157 in soil batch on day t (CFU) | $(1 - Pr_{man}) \times SP_t \times R_{dsP} + Pr_{man} \times \frac{SP_t}{10^{R_{dmanP}}}$ |
| $SN_{t+1}$ | Non-persister ECO157 count in soil batch on day t+1 (CFU) | $SN_t + Irr_{S_t} + Pig_{S_t} + Runoff_{S_t} - (R_{splash_t} + I_{splash_t}) \times SN_t - SNtoSP_t - SNdecay_t$ |
| $SP_{t+1}$ | Persister ECO157 count in soil batch on day t+1 (CFU) | $SP_t + SNtoSP_t - (R_{splash_t} + I_{splash_t}) \times SP_t - SPdecay_t$ |
| $CS_{t+1}$ | Total ECO157 count (non-persister and persister) in soil batch on day t+1 (CFU) | $SN_{t+1} + SP_{t+1}$ |
| $Irr_{L_t}$ | ECO157 count introduced to romaine batch via irrigation on day t (CFU) | One of (depending on the irrigation method for the iteration: O=overhead spray, F=furrow, D=drip):<br>$Irr_L(f_{irr} = O)_t = LI_t \times Bs_t \times P_{irr} \times fO_t \times BwaitP_t$<br>$Irr_L(f_{irr} = F)_t = LI_t \times Bs_t \times P_{irr} \times fF_t \times BwaitP_t$<br>$Irr_L(f_{irr} = D)_t = LI_t \times P_{irr} \times Pr_D$ |
| $Pig_{L_t}$ | ECO157 count introduced to romaine batch via feral swine on day t (CFU) | $LW \times pPEC_t$ |
| $Runoff_{L_t}$ | ECO157 count introduced to romaine batch via runoff on day t (CFU) | $LSR \times pRunoff_t$ |
| $LNtoLP_t$ | Switch of non-persister ECO157 to persister cells on day t (CFU) | $LN_t \times NtoP_L$ |
| $LNdecay_t$ | Decay of non-persister ECO157 in romaine batch on day t (CFU) | $LN_t \times R_{dL}$ |
| $LPdecay_t$ | Decay of persister ECO157 in romaine batch on day t (CFU) | $LP_t \times R_{dL_P}$ |
| $LN_{t+1}$ | Non-persister ECO157 count in romaine batch on day t+1 (CFU) | $LN_t + Irr_{L_t} + Pig_{L_t} + Runoff_{L_t} - (R_{splash_t} + I_{splash_t}) \times (SN_t + SP_t) - LNtoLP_t - LNdecay_t$ |
| $LP_{t+1}$ | Persister ECO157 count in romaine batch on day t+1 (CFU) | $LP_t + LNtoLP_t - LPdecay_t$ |
| $C_{L_{t+1}}$ | Total ECO157 count (non-persister and persister) in romaine batch on day t+1 (CFU) | $LN_{t+1} + LP_{t+1}$ |

### Supplementary Table S6. Scenario analysis for the effect of irrigation water.

| Scenario | Definition (Reference) | ID of parameters affected (unit) | Parameter value in the scenario | Predicted illness cases resulted from the tested scenario: 5th; 50th; 95th percentile |
|---|---|---|---|---|
| Baseline | No changes made in the model. | None | All parameters were at their baseline conditions. | 42; 19,040; 3.6×10$^6$ |
| Decrease the ECO157 prevalence in well waters by 10-fold. | The well water prevalence decreased from 0.1% to 0.01%. | $P_{irr_W}$ (unitless) | $P_{irr_W}$=0.0001 instead of $P_{irr_W}$=0.001 | 42; 15,091; 1.6×10$^6$<br>Median number of cases decreased by 20.7%. |
| Microbiological testing limit set by the more restrictive 2023 LGMA was used to parameterize surface irrigation water intended for overhead spray irrigation | For Type B agricultural water intended for overhead spray irrigation within 21 days of harvest, the maximum generic *Escherichia coli* count in any single water sample should not exceed 235 MPN/100ml, or $Log_{10}(2.35)$. | $C_{irr_S}$ (CFU/ml) | $C_{irr_S}$ from … $Pert(0.1, \mathbf{0.76}, 1.77)$… to …$Pert(0.1, \mathbf{0.37}, 1.77)$… | 50; 13,122; 1.7×10$^6$<br>Median number of cases reduced by 31.1%. |

### Supplementary Table S7. Scenario analysis for the effect of assuming no cell presence on romaine before 14-day countdown period.

| Scenario | Definition (Reference) | ID of parameters affected (unit) | Parameter value in the scenario | Predicted illness cases resulted from the tested scenario: 5th; 50th; 95th percentile |
|---|---|---|---|---|
| Baseline | No changes made in the model. | None | All parameters were at their baseline conditions. | 42; 19,040; 3.6×10$^6$ |
| 100 CFU ECO157 per romaine batch at the start of the 14-day countdown | Each romaine batch started with 100 CFU of ECO157 at the start of the 14-day countdown, rather than 0 CFU. | $LN_0$ and $LP_0$ (CFU) | $LN_0 = 99$ CFU and $LP_0 = 1$ CFU instead of both being 0. | 39; 18,955; 4.7×10$^6$<br>Median number of cases reduced by 0.5%.<br><br>(The ECO157 prevalence increased from 1% to 1.1%. However, 45.1% of the contamination consisted of < 3 $Log_{10}$ CFU per romaine batch, compared to the 41.6% predicted at baseline. This shift resulted in a decrease in the median number of illness cases, as the lower number of cells in the percentiles reduced the overall risk.) |
| 1,000 CFU ECO157 per romaine batch at the start of the 14-day countdown | Each romaine batch started with 1000 CFU of ECO157 at the start of the 14-day countdown, rather than 0 CFU. | $LN_0$ and $LP_0$ (CFU) | $LN_0 = 998$ CFU and $LP_0 = 2$ CFU instead of both being 0. | 692; 1,644; 6.9×10$^4$<br>Median number of cases reduced by 91.4%.<br><br>(The ECO157 prevalence increased from 1% to 38%. However, 77.5% of the contamination consisted of just 1 CFU per romaine batch, which drastically reduced the median number of illness cases due to the impact of single cells on the percentiles.) |

**Supplementary Table S8. Correlation of parameters with the *Escherichia coli* O157:H7 (ECO157) counts in romaine batch at harvest in the preharvest ECO157 model. Parameters are categorized based on their inclusion in the Spearman rank-order correlation coefficient (SRCC) analysis.**

| Parameter | Description (units) | Included in Preharvest SRCC (Yes, No or NA [a]. If Yes, SRCC value) [b] | Comment |
|---|---|---|---|
| $A_{field}$ | Area of the field (approximately 1-acre field) ($cm^2$) | NA | Constant [c] |
| $A_{batch}$ | Area of a field that grows a romaine batch ("batch area", 1/12 of the field area) ($cm^2$) | NA | Constant [c] |
| $N_{plant}$ | Number of plants harvested from the field (romaine plants) | NA | Constant [c] |
| $Plant_{width}$ | The area that a single mature romaine plant occupied on the field, length x width of one plant ($cm^2$) | NA | Constant [c] |
| $Plant_{space}$ | The area allocated for one romaine plant in the field ($cm^2$) | NA | Constant [c] |
| $Plant_{barea}$ | Number of plants grown in a batch area (romaine plants) | NA | Constant [c] |
| $Plant_{wght}$ | Weight of one mature romaine plant (g) | NA | Constant [c] |
| $f_{loc}$ | Select geographic location where the romaine batches were grown (unitless). Notations:<br>CC: Coastal Region and Central Valley, CA<br>DR: Desert (Imperial) Region, CA<br>Y: Yuma Region, AZ | NA | Categorical [c] |
| $f_{month}$ | Select the month of harvest for the given geographic location (unitless). Notations:<br>January: 1, February: 2, March: 3, April: 4, May: 5, June: 6, July: 7, August: 8, September: 9, October: 10, November: 11, December: 12 | NA | Categorical [c] |
| $f_w$ | Select the source of irrigation water (unitless). Notations:<br>S: Surface water<br>W: Well water | NA | Categorical [c] |
| $f_{irr}$ | Select the irrigation type for the given geographic location (unitless). Notations:<br>O: Overhead spray irrigation<br>F: Furrow irrigation<br>D: Drip irrigation | NA | Categorical [c] |
| $Modeldays$ | Countdown period before harvest, modeled via a set of difference equations (day) | NA | Constant [c] |
| $edge$ | Batch area was at the edge of the field and thus could be contaminated by runoff (unitless): no=0 (baseline), yes=1 | 0.0573 | - |
| $C_{man}$ | ECO157 count per g of BSAAO (CFU/g) | No | Included in $C_{man_P}$ |
| $Sc_{vac}$ | Cattle vaccination intervention against ECO157 (unitless): no=0 (baseline), yes=1 | NA | Assessed only in scenario analysis. |
| $Rh_{con}$ | Volume of 1g BSAAO/swine feces/cattle feces ($cm^3$/g) | NA | Constant [c] |
| $Dh_{con}$ | Depth of tiling on the field (cm) | No | Included in $DiF_{man}$ |
| $M_{man}$ | Mass of BSAAO applied on the batch area (g) | No | Included in $DiF_{man}$ |
| $DiF_{man}$ | Dilution of ECO157 cells in BSAAO in the volume of the tilled soil batch layer (unitless) | -0.0002 | - |
| $Pr_{man}$ | BSAAO was used (unitless): no=0, yes=1 | No | Included in $C_{man_P}$ |
| $P_{man}$ | BSAAO applied on a field was contaminated with ECO157 (unitless): no=0, yes=1 | No | Included in $C_{man_P}$ |
| $C_{man_P}$ | ECO157 count in any BSAAO applied soil, which returns 0 CFU for a non-contaminated soil and $C_{man}$ for a contaminated soil (CFU/g) | 0.0024 | - |
| $P_{irr_S}$ | ECO157 prevalence in surface water (unitless) | No | Included in $C_{irr_P}$ |
| $P_{irr_W}$ | ECO157 prevalence in well water (unitless) | No | Included $C_{irr_P}$ |
| $P_{irr}$ | Irrigation water contaminated with ECO157 applied during the countdown period (unitless): no=0, yes=1 | No | Included $C_{irr_P}$ |
| $Sc_{wt}$ | Water treatment intervention applied prior to the overhead spray irrigation (O) using surface water (S) (unitless): None=0 (baseline), Ultra-Violet (UV)=1, Peracetic Acid (PAA)=2, Chlorine=3 | NA | Assessed only in scenario analysis. |
| $C_{irr_S}$ | ECO157 count per ml of contaminated surface water post treatment (CFU/ml) | No | Included in $C_{irr_P}$ |
| $C_{irr_W}$ | ECO157 count per ml of contaminated well water (CFU/ml) | No | Included in $C_{irr_P}$ |
| $C_{irr}$ | ECO157 count per ml of contaminated irrigation water applied on day t (CFU/ml) | No | Included in $C_{irr_P}$ |

| | | | |
|---|---|---|---|
| $C_{irr_P}$ | ECO157 count on any irrigation water, which returns 0 CFU for a non-contaminated water and $C_{irr}$ for a contaminated water (CFU/ml) | 0.2906 | - |
| $type_{irr}$ | Probability the selected irrigation type was occurring on a given day (unitless) | NA | Categorical [c] |
| $Pr_O$ | Probability of overhead spray irrigation on day t (unitless) | -0.0033 | Included as $fO_{t_{Sum}}$ [e] |
| $Pr_F$ | Probability of furrow irrigation on day t (unitless) | 0.0044 | Included as $fF_{t_{Sum}}$ [f] |
| $Pr_D$ | Probability of drip irrigation on day t (unitless) | NA | Constant [c] |
| $W_O$ | Volume of water applied through overhead spray irrigation to a romaine batch per an irrigation event (ml/day) | No | Included in $g_{irr}$ |
| $W_F$ | Volume of water applied through furrow irrigation to a romaine batch per an irrigation event (ml/day) | No | Included in $g_{irr}$ |
| $W_D$ | Volume of water applied through drip irrigation to a romaine batch per an irrigation event (ml/day) | No | Included in $g_{irr}$ |
| $Daytogrow$ | Days from seeding to harvest (day) | -0.0116 | - |
| $D_{man}$ | Number of days before BSAAO application (day) | NA | Constant [c] |
| $g_{irr}$ | Volume of water applied per an irrigation event in a batch area (ml/day) | -0.0064 | - |
| $V_O$ | Volume of irrigation water captured per g of romaine per an overhead spray irrigation event (ml/g/day) | No | Included in $v_{irr}$ |
| $V_F$ | Volume of irrigation water captured per romaine batch per a furrow irrigation event (ml/day) | No | Included in $v_{irr}$ |
| $V_D$ | Volume of irrigation water captured per romaine batch per a drip irrigation event (ml/day) | No | Included in $v_{irr}$ |
| $v_{irr}$ | Volume of irrigation water captured by romaine batch per an irrigation event (ml/day) | 0.0581 | - |
| $Tr_{man_O}$ | Fraction of the top 1cm layer of soil batch transferred to a romaine batch via splash caused by overhead spray irrigation per irrigation event (unitless) | No | Included in $M_{irr}$ |
| $Tr_{man_F}$ | Fraction of the top 1cm layer of soil batch transferred to a romaine batch via splash caused by furrow irrigation per irrigation event (unitless) | No | Included in $M_{irr}$ |
| $Tr_{man_D}$ | Fraction of the top 1cm layer of soil batch transferred to a romaine batch via splash caused by drip irrigation per irrigation event (unitless) | No | Included in $M_{irr}$ |
| $M_{irr}$ | Mass of soil transferred to a romaine batch via splash per irrigation event (g/day) | 0.0553 | - |
| $Tr_{ecirr}$ | Fraction of ECO157 cells transferred from irrigation water to romaine (unitless) | NA | Constant [c] |
| $DiF_{irr}$ | Dilution factor for ECO157 cells in irrigation water in the top 1cm layer of soil batch (unitless) | NA | Calculated by parameters included directly in the SRCC analysis. |
| $LN_0$ | ECO157 count in a romaine batch at the start of the 14-day countdown period (t=0) (CFU) | NA | Constant [c] |
| $CC_{UV}$ | $Log_{10}$ ECO157 count reduction of surface water contamination by UV treatment before overhead spray irrigation ($Log_{10}$CFU) | NA | Assessed only in scenario analysis. |
| $CC_{PAA}$ | $Log_{10}$ ECO157 count reduction of surface water contamination by PAA treatment before overhead spray irrigation ($Log_{10}$CFU) | NA | Assessed only in scenario analysis. |
| $CC_{Ch}$ | $Log_{10}$ ECO157 count reduction of surface water contamination by Chlorine treatment before overhead spray irrigation ($Log_{10}$CFU) | NA | Assessed only in scenario analysis. |
| $C_{cow_{prep}}$ | $Log_{10}$ ECO157 count on fresh cattle feces ($Log_{10}$CFU/g) | No | Included in $C_{cow_P}$ |
| $C_{cow_{tot}}$ | $Log_{10}$ ECO157 count on fresh cattle feces, accounting for the effect of ECO157 vaccine intervention, if implemented ($Log_{10}$CFU/g) | No | Included in $C_{cow_P}$ |
| $C_{cow}$ | ECO157 count on fresh cattle feces (CFU/g) | No | Included in $C_{cow_P}$ |
| $C_{cow_P}$ | ECO157 count on any cattle feces, which returns 0 CFU for non-contaminated feces and $C_{cow}$ for contaminated feces (CFU/g) | 0.0686 | - |
| $C_{pig}$ | $Log_{10}$ ECO157 count on fresh swine feces ($Log_{10}$CFU/g) | No | Included in $C_{pig_P}$ |
| $C_{pig_P}$ | ECO157 count on any swine feces, which returns 0 CFU for non-contaminated feces and $C_{pig}$ for contaminated feces (CFU/g) | 0.1149 | - |
| $Pr_{pig}$ | Probability of feral swine entering the field on day t (unitless) | No | Included as $Pr_{pigCountSum}$ [g] |
| $Pr_{pig_{countt}}$ | Wildlife intrusion occurred on day t (unitless): no=0, yes=1 | No | Included as $Pr_{pigCountSum}$ [g] |
| $Pr_{pigCountSum}$ | Total number of days during the 14-day countdown when wildlife intrusion occurred (day) | 0.1180 | - |
| $Mdef_{pig}$ | Mass of feces excreted by a feral swine per day (g swine feces/day) | NA | Constant [c] |

| | | | |
|---|---|---|---|
| $Tdef_{pig}$ | Number of times a feral swine defecated per day (times/day) | NA | Constant [c] |
| $Ndef_{pig}$ | Number of times feral swine (between 1-5 swine) defecated on the field (times/day) | -0.0008 | - |
| $M_{pig}$ | Mass of feces excreted by feral swine intruding the batch area (g) | NA | Calculated by parameters included directly in the SRCC analysis. |
| $DiF_{pig}$ | Dilution factor for ECO157 cells in feral swine feces in soil batch associated with plants near which the swine defecated (unitless) | NA | Calculated by parameters included directly in the SRCC analysis. |
| $M_{cow}$ | Mass of cattle runoff (carrying fresh cattle feces only) introduced into the edge batch area soil per runoff event (g/day) | 0.0037 | - |
| $DiF_{cow}$ | Dilution factor for ECO157 cells in cattle runoff introduced into in the top 1cm layer of soil batch (unitless) | NA | Calculated by parameters included directly in the SRCC analysis. |
| $Pr_{LtoS}$ | Proportion of fecal matter (swine and runoff) landing on romaine rather than in soil (unitless) | NA | Constant [c] |
| $Tr_{ecsoil}$ | Proportion of ECO157 cells in soil transferring to romaine during a rain or irrigation event (unitless) | 0.0045 | - |
| $R_{dman}$ | ECO157 decay rate for non-persisters in BSAAO amended soil ($Log_{10}$CFU/day)[a] | NA | Constant [c] |
| $R_{dL}$ | ECO157 decay rate for non-persisters on romaine in field (1/day) [a,b] | NA | Calculated by parameters included directly in the SRCC analysis. |
| $R_{ds}$ | ECO157 decay rate for non-persisters in soil (1/day) [a] | NA | Constant [c] |
| $R_{dman_P}$ | ECO157 decay rate for persisters in BSAAO amended soil ($Log_{10}$CFU/day) [a] | NA | Constant [c] |
| $R_{dL_P}$ | ECO157 decay rate for persisters on romaine in field (1/day) [a] | NA | Calculated by parameters included directly in the SRCC analysis. |
| $R_{ds_P}$ | ECO157 decay rate in soil for persisters (1/day) [a] | NA | Constant [c] |
| $NtoP_{man}$ | Switch rate from normal to persister in BSAAO amended soil (1/day) [a] | NA | Constant [c] |
| $NtoP_L$ | Switch rate from normal to persister on romaine (1/day) [a] | NA | Constant [c] |
| $NtoP_S$ | Switch rate from normal to persister in soil (1/day) [a] | NA | Constant [c] |
| $Pmax_{man}$ | Maximum fraction of persisters in BSAAO amended soil (unitless) | NA | Constant [c] |
| $Pmax_S$ | Maximum fraction of persisters in soil (unitless) | NA | Constant [c] |
| $Pmax_L$ | Maximum fraction of persisters on romaine (unitless) | NA | Constant [c] |
| $BwaitP$ | Waiting time: The period before harvest during which overhead spray and furrow irrigation was stopped (day) | 0.0020 | - |
| $t$ | Average amount of soil attached to a harvesting blade after contact with soil in one cross contamination event (g/blade) | NA | Constant [c] |
| $Tr_{blade}$ | Proportion of ECO157 cells transferred from a harvesting blade to a romaine plant in one cross contamination event (unitless) | NA | Constant [c] |
| $Tr_{ecblade}$ | ECO157 count in soil attached on blades (CFU/soil batch x soil batch/blade=CFU/blade) | NA | Constant [c] |
| $N_{blade}$ | ECO157 cells transferred from harvesting blades to a romaine batch (CFU/romaine batch) | NA | Calculated by parameters included directly in the SRCC analysis. |
| $CC_{blade}$ | Maximum fraction of persisters in soil (unitless) | NA | Calculated by parameters included directly in the SRCC analysis. |
| $C_{Lh}$ | Maximum fraction of persisters on romaine (unitless) | NA | Calculated by parameters included directly in the SRCC analysis. |
| $P_{sun_m}$ [h] | Probability of a sunny day (unitless) | -0.0086 | Included as $Bs_{tSum}$ [i] |
| $P_{cow_m}$ [h] | ECO157 prevalence in fresh cattle feces (unitless) | No | Included in $C_{cow_P}$ |
| $P_{pig_m}$ [h] | ECO157 prevalence in fresh feral swine feces (unitless) | No | Included in $C_{pig_P}$ |
| $h_{sun_m}$ [h] | Average sunny hours per day (h) | -0.0170 | - |
| $R_{sun_m}$ [h] | Solar decay rate (1/day) | -0.0105 | - |
| $Prec_{norm_m}$ [h] | Precipitation level normalized (unitless) | -0.0004 | - |
| $Prec_{actual_m}$ [h] | Actual (average) level of precipitation during the year (mm) | No | Included in $Prec_{norm_m}$ |

[a] Not applicable.

[b] Bonferroni-corrected alpha threshold is α=0.05/21=0.0024.

[c] Constant value.
[d] Categorical data.
[e] The parameter $fO_{t_{Sum}}$ represents the total number of days when overhead spray irrigation was applied within a 14-day countdown period. It is calculated by adding together the $fO_t$ values from Supplemental Table 1 over a 14-day period.
[f] The parameter $fF_{t_{Sum}}$ represents the total number of days when furrow irrigation was applied within a 14-day countdown period. It is calculated by adding together the $fF_t$ values from Supplemental Table 1 over a 14-day period.
[g] See Supplementary Table S3 for the relationship between $Pr_{pig}$ and $Pr_{pigCountSum}$.
[h] This parameter is used in the preharvest model and taken from Supplementary Table S3.
[i] The parameter $Bs_{tSum}$ represents the total number of sunny days within a 14-day countdown period. It is calculated by adding together the as $Bs_t$ values from Supplementary Table S3 over a 14-day period.

### Supplementary Table S9. Parameterization of the scenarios.

| Scenario | Parameterization based on Table 1 and Table 2 |
|---|---|
| Cattle vaccination | When the cell value of parameter $Sc_{vac}$ (Supplementary Table S1) was changed from 0 to 1, a 1-Log$_{10}$CFU/g reduction in the ECO157 counts in BSAAO and cattle feces was applied. |
| Surface water treatment for overhead spray irrigation by chlorine | When the cell value of parameter $Sc_{wt}$ (Supplementary Table S1) was changed from 0 to 3, ECO157 count in surface water was reduced using chlorine treatment ($CC_{Ch}$). |
| Surface water treatment for overhead spray irrigation by peracetic acid (PAA) | When the cell value of parameter $Sc_{wt}$ (Supplementary Table S1) was changed from 0 to 2, ECO157 count in surface water was reduced using PAA treatment ($CC_{PAA}$). |
| Surface water treatment for overhead spray irrigation by Ultraviolet radiation (UV) | When the cell value of parameter $Sc_{wt}$ (Supplementary Table S1) is changed from 0 to 1, ECO157 count in surface water was reduced using UV treatment ($CC_{UV}$). |
| Transition from overhead spray to furrow irrigation (i.e., all irrigation was through either furrow or drip) | To apply this scenario, we manually modified the values of $fO_t$ and $fF_t$ (Supplementary Table S1) and transitioned all overhead spray irrigated fields to furrow irrigated fields for all locations. |
| Transition from overhead spray and furrow to drip irrigation (all irrigation was through drip) | To apply this scenario, we manually modified the values of $fO_t$, $fF_t$ and $fD_t$ (Supplementary Table S1) and transitioned of all overhead spray and furrow irrigated fields to drip irrigated fields for all locations. |
| Chlorine wash alternatives with varying Log$_{10}$CFU reduction | When the cell value of parameter $Sc_{wash}$ (Supplementary Table S2) was changed from 1 to another number (2 to 6), the parameter for chlorine wash alternative ($CC_{ch1}, CC_{ch2}, CC_{ch3}, CC_{ch4} or\ CC_{ch5}$, respectively) was used instead of the baseline chlorine wash ($CC_{ch20}$) in postharvest. |
| Temperature reduction during retail | To apply this scenario, we manually modified the temperature distribution from Normal to Uniform with reduced temperature range. |
| Consumer wash | When the cell value of parameter $Sc_{conw}$ (Supplementary Table S2) was changed from 0 to 1, a total of 62% of the consumer washed the product before consumption, instead of 0%. |
| Clustering effect | When the cell value of parameter $Sc_{clstrb}$ (Supplementary Table S2) was changed from 0 to 1, clustering was allowed during partitioning of romaine batch to packages. Similarly, clustering from package to servings was allowed when the cell value of parameter $Sc_{clstrp}$ (Supplementary Table S2) was changed from 0 to 1. Parameters $Clstr_{parb}$ (Supplementary Table S2) and $Clstr_{parp}$ (Supplementary Table S2) determined the level of clustering in packages and servings, respectively. |

**Supplementary Table S10. Correlation of variable parameters with the *Escherichia coli* O157:H7 (ECO157) counts in romaine batches at harvest in the preharvest irrigation model (considering only the three irrigation systems with no other preharvest sources), as well as in irrigation models considering each individual irrigation source contamination separately. Parameters were categorized based on their inclusion in the Spearman rank-order correlation coefficient (SRCC) analysis.**

| Irrigation System Parameter (Definition) | Irrigation contamination only [a] | Contamination via overhead spray irrigation only [a] | Contamination via furrow irrigation only [a] | Contamination via drip irrigation only [a] |
|---|---|---|---|---|
| $C_{irr\,P}$ (ECO157 count on any irrigation water, which returns 0 CFU for a non-contaminated water and $C_{irr}$ for a contaminated water (CFU/ml)) | 0.41 | 0.38 | 0.13 | 0.08 |
| $v_{irr}$ (Volume of irrigation water caught by romaine batch per an irrigation event) | 0.09 | 0.09 | 0.03 | -0.02 |
| $M_{irr}$ (Mass of soil transferred to a romaine batch via splash per irrigation event) | 0.08 | 0.09 | 0.01 | -0.02 |

[a] The values indicate ρ, the Spearman rank-order correlation coefficient, calculated for a particular irrigation system. Only ρ values of 0.05 or higher, and -0.05 or lower, are shown (ρ values between -0.05 and 0.05 are indicated with "-"). Interpretation of coefficient absolute values: |ρ| 0 to <0.1, none; ρ=0.1 to <0.3, poor; ρ=0.3 to <0.6, fair; ρ=0.6 to <0.8, moderate; ρ=0.8 to 1.0 very strong correlation.

**Supplementary Table S11. Predicted hospitalizations, cases of hemolytic uremic syndrome (HUS), and mortality following *E. coli* O157:H7 (ECO157) illness from vegetable row crops *[102]*.**

| Probability of hospitalization among predicted ECO157 illness cases (%) | Predicted hospitalizations: 5th; 50th ; 95th percentile | Probability of HUS among predicted ECO157 illness cases (%) | Predicted HUS cases: 5th; 50th ; 95th percentile | Probability of death among predicted ECO157 illness cases (%) | Predicted death: 5th; 50th ; 95th percentile |
|---|---|---|---|---|---|
| 30 | 13; 5,712; $1.1\times10^{6}$ | 4 | 2; 762; $1.4\times10^{5}$ | 0.3 | 0; 57; $1.1\times10^{4}$ |

**Supplementary Table S12. Scenario analysis for the effect of persister cells.**

| Scenario No | Scenario | Definition (Reference) | ID of parameters affected (unit) | Parameter value in the scenario | Predicted illness cases resulted from the tested scenario: 5th; 50th; 95th percentile |
|---|---|---|---|---|---|
| 0 | Baseline | No changes made in the model. | None | All parameters were at their baseline conditions. | 42; 19,040; $3.6\times10^{6}$ |
| 1 | Switch rate from normal to persister in BSAAO amended soil was increased | Switch rate was increased by 100 times | $NtoP_{man}$ | $NtoP_{man} = 0.001\ \times 100 = 0.1$ | 48; 17,711; $3.2\times10^{6}$ Median number of cases reduced by 7.0%. |
| 2 | Switch rate from normal to persister on romaine was increased | Switch rate was increased by 1,000 times | $NtoP_L$ | $NtoP_L = 0.0004\ \times 1000 = 0.4$ | 23; 2,733; $7.1\times10^{5}$ Median number of cases reduced by 85.7%. |
| 3 | Switch rate from normal to persister in soil was increased | Switch rate was increased by 1,000 times | $NtoP_S$ | $NtoP_S = 0.0001\ \times 1000 = 0.1$ | 44; 19,848; $4.7\times10^{6}$ Median number of cases increased by 4.2%. |
| 4 | No non-persisters were included in both soil and romaine | Only persister cells were included at harvest; only persisters were transferred to postharvest model. | $SP_t$ and $LP_t$ | $C_{St_N} = 0$ and $C_{Lt_N} = 0$ | 9; 67; 6,578 Median number of cases reduced by 99.7%. |
| 1, 2, 3 & 4 | No non-persisters were included in both soil and romaine, and all switch rates were increased | Only persister cells were included at harvest, and all the switch rates were increased. | $SP_t$, $LP_t$, $NtoP_{man}$, $NtoP_L$ and $NtoP_S$ | $C_{St_N} = 0$ and $C_{Lt_N} = 0$ $NtoP_{man} = 0.001\ \times 100 = 0.1$ $NtoP_L = 0.0004\ \times 1000 = 0.4$ $NtoP_S = 0.0001\ \times 1000 = 0.1$ | 17; 992; $3.4\times10^{5}$ Median number of cases reduced by 94.8% |

### Supplementary Table S13. Scenario analysis for the effect of preharvest dilution depth.

| Scenario | Definition (Reference) | ID of parameters affected (unit) | Parameter value in the scenario | Predicted illness cases resulted from the tested scenario: 5th; 50th; 95th percentile |
|---|---|---|---|---|
| Baseline | No changes made in the model. | None | All parameters were at their baseline conditions. | 42; 19,040; $3.6\times10^{6}$ |
| Dilution depth was increased | Dilution depth for the irrigation, swine feces and runoff were increased from 1cm to 5cm. | $DiF_{irr}, DiF_{pig}, DiF_{cow}$ (unitless) | The denominator of $DiF_{irr}, DiF_{pig}, DiF_{cow}$ were changed from $A_{batch} \times 1$ to $A_{batch} \times 5$ | 48; 16,447; $2.7\times10^{6}$<br>Median number of cases reduced by 13.5%. |

### Supplementary Table S14. Scenario analysis for the removal of exterior leaves under the conservative assumption of uniform distribution of ECO157 across leaves.

| Scenario | Definition (Reference) | ID of parameters affected (unit) | Parameter value in the scenario | Predicted illness cases resulted from the tested scenario: 5th; 50th; 95th percentile |
|---|---|---|---|---|
| Baseline | No changes made in the model. | None | All parameters were at their baseline conditions. | 42; 19,040; $3.6\times10^{6}$ |
| Removal of exterior leaves; 20% [a] | A total of 20% reduction in the ECO157 count on romaine under the conservative assumption of uniform distribution of ECO157 across leaves. | $C_{Lh}$(CFU/romaine batch) and $Plant_{wght}$ (g) | $C_{Lh} \times 0.80$ and $Plant_{wght} \times 0.8$ | 48; 19,205; $3.6\times10^{6}$<br>Median number of cases increased by 0.09%. |
| Removal of exterior leaves; 23.8% [a,] | A total of 23.8% reduction in the ECO157 count on romaine, under the assumption that ECO157 contamination was 25% more on an exterior leaf compared to an interior leaf. | $C_{Lh}$(CFU/romaine batch) and $Plant_{wght}$ (g) | $C_{Lh} \times 0.762$ and $Plant_{wght} \times 0.8$ | 47; 18,437; $3.5\times10^{6}$<br>Median number of cases reduced by 3.2%. |
| Removal of exterior leaves; 28.2% [a] | A total of 28.2% reduction in the ECO157 count on romaine under the assumption that ECO157 contamination was 50% more on an exterior leaf compared to an interior leaf. | $C_{Lh}$(CFU/romaine batch) and $Plant_{wght}$ (g) | $C_{Lh} \times 0.727$ and $Plant_{wght} \times 0.8$ | 46; 17,724; $3.4\times10^{6}$<br>Median number of cases reduced by 6.9%. |
| Removal of exterior leaves; 33.3% [a] | A total of 33.3% reduction in the ECO157 count on romaine under the assumption that ECO157 contamination was 100% more on an exterior leaf compared to an interior leaf. | $C_{Lh}$(CFU/romaine batch) and $Plant_{wght}$ (g) | $C_{Lh} \times 0.666$ and $Plant_{wght} \times 0.8$ | 45; 16,468; $3.1\times10^{6}$<br>Median number of cases reduced by 13.5%. |

[a] A whole romaine weighed 300g before the exterior leaves were removed. If a plant was assumed to have 10 leaves and 2 exterior leaves were removed, the final weight of the romaine would be 240g and calculated as $Plant_{wght} \times 0.8$.
